# Exploratory Deep Phenotyping of Early Antidepressant Treatment Effects in a Longitudinal, Double-Blind, Randomized, Controlled, Parallel-Group Pre-Post Design – a Study Protocol from Research Consortium OptiMD

**DOI:** 10.64898/2025.12.01.25341376

**Authors:** André Manook, Andreas Hiergeist, Gerhard Liebisch, Ulrich Dischinger, Agnese Petrera, Jens Schwarzbach, André Gessner, Oliver Gruber, Thomas C. Baghai

## Abstract

**Background:** A better understanding of neurobiological mechanisms behind clinical depression and optimization of pharmacological treatment options remain enormous challenges. Early subtyping of depression might be achieved by discovering reliable trait markers or by early detection of changes in potential state markers, for example during the first days of antidepressant treatment.

**Objective:** The primary objective of this trial was to explore early neurobiological dynamics during the first week of antidepressant treatment within the same drug-naïve inpatients in a large deep-phenotyping exploration applying various measurement modalities as described below.

**Methods:** A longitudinal deep-phenotyping experimental design involved holistic measurements of various markers with state-of-the art technology over five axes of investigation: MR neuroimaging, dexamethasone/CRH-testing, gut microbiome composition, inflammatory proteome and lipidomics. Therapy during these measurements was controlled and standardized by conducting a double-blind, randomized parallel group design with four balanced arms during the first seven days of medication that encompassed three common first-line, yet pharmacologically distinguishable, antidepressants (escitalopram, mirtazapine, agomelatine) or placebo. The complete trial period has been 60 days including a third major longitudinal point of investigation at the end of trial. An additional control cohort with matched healthy volunteers was participating in the same set of investigations as patients at baseline, except dexamethasone/CRH-testing.

**Trial status:** This trial has extensively served two subprojects of the German National Research Consortium OptiMD. It has been registered at the European Clinical Trials Register with identifier 2013-003370-27 and was initiated in January 2016. Recruitment of 70 inpatients and 25 healthy participants has been completed in August 2021 without any known harms in regard to study participation. Collection of data is still ongoing and hence, databases are not yet closed. Analyses of first complete datasets have begun and we hope that our deep-phenotyping exploration might contribute to new perspectives on clinical depression and its treatment.

**Protocol outline:** 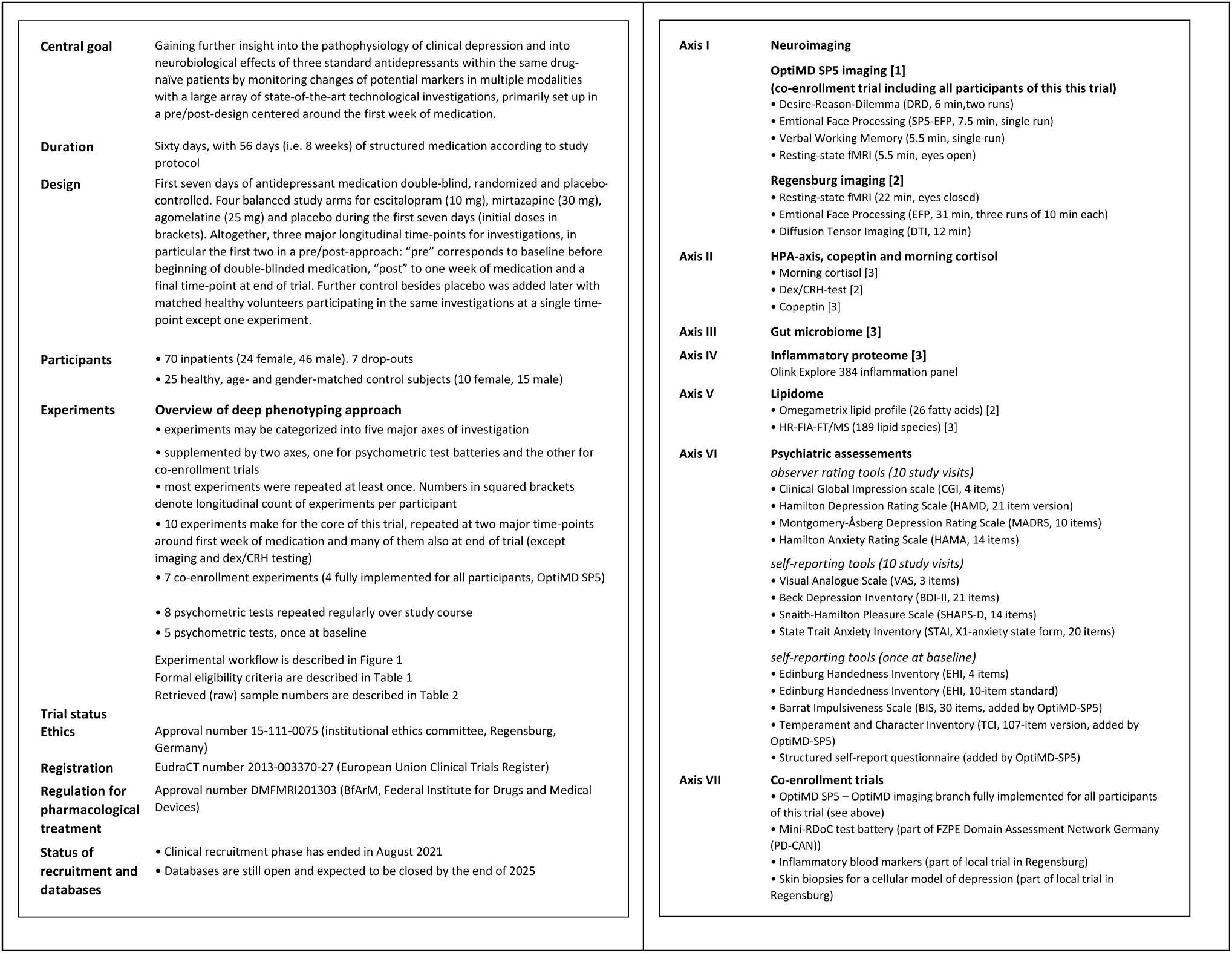

## Introduction

### Background and rationale

#### Clinical depression

Clinical depression expresses in heterogeneous psychiatric and medical phenotypes [1] and affected patients react differently to pharmaceutical interventions with antidepressants [2]. Onset of possible improvement may be observed as early as after one week of treatment in trial settings [3] while response and remission continue to show up to twelve weeks of unchanged treatment with antidepressants in naturalistic environments [4,5]. Therefore, practice guidelines still recommend patience, i.e. watchful waiting, of at least 2 to 4 weeks [6] in favor of switching medication too early. Moreover, more than half of patients with depression do not sufficiently improve during the first six weeks with antidepressant therapy [7–9]. The heterogeneity in symptom constellations and lacking predictability of efficacy on an individual patient level remain a challenge in everyday clinical life. This motivates to continue the exploration of potential diagnostic and predictive biomarkers [10–12]. After a decades-long quest it appears likely that none of them might be potent enough to serve as a single feasible marker. However, larger compound sets of biomarkers from different research modalities might become more helpful for clinical decision making [13]. Sincere hopes remain, that these someday might reliably aid in identifying clinical subtypes of depression and in predicting their possible treatment trajectories to various antidepressants and other available modes of therapy.

#### Deep phenotyping

Biomarkers [10] are mediators for phenotyping and response evaluation. Deep phenotyping [14] describes the process of gathering more comprehensive and detailed information from individuals such that more granular phenotypes may be constructed. While the idea may not be new, the term was coined when high-throughput methods gained momentum at the beginning of this century [15]. It reflects an experimental approach of collecting a large spectrum of data from individuals for consecutive pattern analysis and personalized predictions. It is noteworthy, that the increasingly employed adjective ‘deep’ as used here, originates from a humoristic novel written by Douglas Adams in 1979 [16] in which a utopian supercomputer was given the name ‘Deep Thought’ [17].

At one end of the spectrum, deep phenotyping aims at collecting the maximum amount of data from single individuals, for example in well-conceived N-of-1 trials [18], which is a study design increasingly gaining interest as an alternative to randomized controlled trials for its considerably lower biological variability [19]. At the other end of the deep phenotyping spectrum, extensive data collection encompasses whole populations [20].

#### Potential Biomarkers

##### Cortisol and HPA reactivity

The heterogeneity and complexity of clinical phenomena summarized in the diagnosis of clinical depression has understandably caused postulation and exploration of a myriad of potential diagnostic (i.e. trait) and predictive (i.e. state) biomarkers, so far. Neuroimaging, neuroendocrinologic, genetic and inflammation markers are probably the most prevalent ones besides metabolic and microbiome findings [21]. Up to now, none of these biomarkers appear to withstand meta-analytic assessments [22] which might be due to trial designs, sample sizes and their applied statistical methods.

The single biomarker that does show some predictive value in certain clinical situations appears to be reactivity along the hypothalamic-pituitary-adrenal (HPA) axis, as for example measured with the combined dexamethasone-suppression/corticotropin-releasing hormone stimulation (dex/CRH) test [22–26]. In particular, it appears to have relevant predictive validity in regard to relapse prediction [27,28]. However, a robust predictive validity of dex/CRH testing in regard to antidepressant response behavior remains controversial [29].

The responses after CRH-challenge tend to be elevated in depressed patients in the beginning and often attenuate in those who gain from antidepressant therapy [29]. Hence, it has been proposed a surrogate marker for clinical efficacy [30]. The dex/CRH test and its longitudinal response behavior during antidepressant treatment have been well established over the last 35 years and besides a few exceptions [31,32], which do not apply to this trial, cortisol and ACTH (adrenocorticotropic hormone, i.e. corticotropin) responses usually behave similar. Hence, ACTH analysis was no longer included in the version of the test applied here.

Kiem et al. have built a first bridge between dex/CRH testing and neuroimaging by combining the dex/CRH test with resting-state functional magnetic resonance imaging (rfMRI) in young healthy men [33]. This study revealed mostly inverse associations between connectivity of some brain networks and HPA reactivity. Particularly, lower connectivity between left and right hippocampus significantly predicted stimulated cortisol concentrations.

##### Copeptin

Overreactivity of arginine vasopressin (AVP) is postulated to be a central component in pathological dex/CRH test results [34]. Along the HPA-axis, AVP, formerly known as antidiuretic hormone, and CRH, synergistically signal the pituitary to secret corticotropin which activates release of cortisol from the adrenal cortex [35]. Further release of corticotropin is controlled by negative feedback through cortisol. In dex/CRH testing, dexamethasone also suppresses corticotropin release but it is not able to pass the blood-brain-barrier [36]. Thus, dexamethasone causes a central cortisol deficit which the brain tries to compensate by increasing AVP secretion in order to amplify CRH effects. In physiological HPA-axis regulation, centrally secreted AVP and CRH will only partially succeed in a so-called ‘escape of dexamethasone suppression’ [37]. Depressed patients, however, frequently show a clear ‘escape’ of dexamethasone suppression, presumably, as a result of hypothalamic AVP hypersecretion resulting in a hyperdrive of the HPA axis.

Copeptin is a C-terminal glycopeptide of 39 amino acid residues [38] cleaved from a large precursor peptide (pre-provasopressin) together with AVP and neurophysin II in equimolar ratios – hence, it is also known under the short name CT-proAVP. Its larger size and longer in vivo stability make it more feasible to quantify than vasopressin itself [39] which is why it has been considered as a surrogate marker for AVP. Furthermore, there has not yet been experimental evidence on a peripheral function of copeptin in contrast to the role of peripheral AVP in fluid homeostasis [40]. Hence, in endocrinology, copeptin has already arrived in clinical routine for differential diagnosis of polyuria–polydipsia syndromes like central versus nephrogenic diabetes insipidus.

In psychiatry, clinical studies with copeptin have been scarce. Here, copeptin has been investigated from two angles. On one hand, its concentrations in the afternoon after dexamethasone suppression but immediately before CRH challenge correlate with cortisol secretion after CRH challenge. As a surrogate marker for AVP, this not only further supports the concept of central AVP hypersecretion but also makes copeptin a predictor and, hence, a potential surrogate marker for HPA reactivity as expressed in dex/CRH testing [41,42]. This possible predictive validity for HPA reactivity may let copeptin evolve into a simpler and more feasible biomarker than the dex/CRH test in the future.

On the other hand, morning baseline copeptin without any previous HPA-axis manipulation, has shown strong potential for predictive validity of antidepressant response in a pilot study with 14 patients [43].

##### Inflammatory proteome

Properly reacting to environmental stressors is considered essential for survival and hence, an evolutionary advantage [44]. In this, a well-regulated HPA-axis plays a central role. Similarly, inflammation is an equally existential response to protect an organism after invasion or trauma [45]. However, stress, whether regulated physiologically or dysregulated as described for HPA overreactivity above, also activates inflammatory pathways, and even more so in individuals affected by clinical depression [46]. Systemic inflammatory diseases often coincide with depressive symptoms [47,48] and conversely, pro-inflammatory challenge trials in healthy subjects may cause interim psychopathologies similar to clinical depression [49]. Moreover, at least a third of patients with depression show a prevalence of inflammation in regard to measurable C-reactive protein (CRP) levels [50] or other pro-inflammatory cytokines. Besides CRP, associations with interleukin-6 (IL-6) concentrations appear to be the most robust [51], but tumor necrosis factor-alpha (TNF-α) and interleukin-1β (IL-1β) also appear to play major roles [52]. This growing evidence on the strong influences of the immune system on emotions has led to the establishment of immunopsychiatry as a new sub-specialty [53,54].

One of the high-throughput methodologies available today in order to gain broad insight into the human proteome is an antibody-based proximity extension assay (PEA). PEA technology evolved from a proximity ligation assay [55] and was first described in 2011 [56] and commercialized by Olink. The so-called ‘Target’ platform was published in 2014 and offers immunoassay panels with more than 90 targets per assay [57]. Its even higher throughput variant with Next-Generation Sequencing (NGS) was published in 2021 [58] and provides the methodological basis for the so-called ‘Explore’ platform which comes with the capacity for an even larger spectrum of possible protein targets. Hence, Olink Explore has recently been employed for such large projects as the characterization of plasma proteomic profiles in 54219 participants [59] of the UK Biobank project targeting 2,923 unique proteins per participant that were measured with eight 384-plex panels. One of these eight 384-plex panels is the so-called ‘Explore384 Inflammation’ panel containing 368 unique protein assays. Assays like these may help to gain further insight into inflammatory processes in general and specifically, into neuroinflammatory mechanisms of clinical depression. Among these protein markers, there are 14 C-C motif chemokines and 38 interleukins or their subunits.

##### Lipidome

Lipids may act as physiological pro- or anti-inflammatory signals [60,61], depending on their type and composition, thus, linking them to the inflammatory proteome. Furthermore, specialized pro-resolving lipid mediators play a central role in the physiological termination of inflammatory processes [62].

Long before the field of lipidomics came to light, the analysis of individual lipid species in Greenland Eskimos provided first insight into their role as possible mediators of health and disease [63] in the 1970s. First observations on altered lipid profiles in patients with depression were made in the 1990s [64] and indices that were derived from a small set of fatty acids, like omega-3 index [65], have become well-known descriptors for potential risk in different areas of human health [66].

Coinciding with the emergence and evolution of high-throughput methods, the concepts of ‘lipidome’ [67], functional lipidomics [68] and lipidomics [69] were postulated and the field began to flourish along a large U.S. American funding initiative [70] referred to as the LIPID MAPS consortium.

Comprehensive serum lipidomic profiling has already shown that it may achieve and even exceed the reliability of conventional diagnostic biomarkers, for example in the detection of pancreatic cancer [71]. Along this trend, quite recent work has shown that lipid compositions in psychiatric diseases show clearly distinguishable alterations [72,73]. As of now, however, these lipidomic signatures largely overlap between schizophrenia, bipolar disorder and depression making them general trait markers for broadly being affected by a psychiatric disorder.

##### Gut microbiome

Over the last twenty years, many valuable studies have shown how the gut microbiome is linked to HPA-axis regulation and the immune system [74]. Its interactions with the brain via a multi-facetted gut-brain-axis created substantial hopes towards relevant biomarkers for clinical depression and its monitoring [75]. Convincing preclinical work on interactions between the modern gut microbiome and stress, anxiety and depression began not later than with preclinical work by Sudo et al. in 2004 [76] and continued in many further interesting preclinical studies [75]. The first human case–control data from 37 psychiatric patients with depression versus 18 neurology patients without depression [77] was submitted by Naseribafrouei et al. in autumn of 2013, shortly after the registration of the trial described, here. The study described first differential abundance data with a relatively increased order of Bacteroidales and a relatively reduced family of Lachnospiraceae, in patients with depression which was often confirmed in later reports by others [78], though differential abundance results from patients with depression mostly remain incongruent.

It may be important to clarify the term (human) ‘microbiome’. The concept indeed embraces all microorganisms that inhabit all surfaces and cavities of the human body besides the human eukaryotic cells, themselves. Hence, until recently, the use of the term ‘gut microbiome’ has truly only addressed the bacterial faction of the microbiome, i.e. the human gut bacteriome and has been used interchangeably as it is here. On one hand, there is growing awareness that microbial interactions in the gut should by far not be limited to its bacteriome and that future investigations should also take into account the other biological factions like the virome, the mycobiome, the archaeome and the parasitome as well [79]. On the other hand, merely investigating the gut bacteriome has shown to be such a highly complex endeavor that the research field has mostly remained on the level of describing bacterial compositions and possible associations [78]. However, it would be essential and desirable to establish functional, i.e. causal, connections between the gut and the brain, for example by at least linking bacteriome-specific metabolomic analyses or even metatranscriptomic and metaproteomic investigations [80]. The experimental design constructed for this trial, however, focusses on exploratory descriptive approaches.

##### MR neuroimaging

Neuroimaging has yielded promising predictive biomarkers for depression. Many of them originate from volumetric and functional MRI (fMRI) analyses [81], for example hippocampal volume, task-based-amygdala activation or functional connectivity (FC) to the dorsolateral prefrontal cortex. Pioneering neuroimaging work for clinical depression was performed with positron emission tomography (PET) using [15O]-labelled water. The amygdala had been an early target in imaging of clinical depression [82]. In 1992, PET with [15O]-labelled water showed increased regional cerebral blood flow (rCBF) in the left amygdala and left ventrolateral prefrontal cortex (VLPFC) in depressed patients [83]. The central role of the amygdala in social-affective behavior [84] and the discovery of a universal link between specific human facial expressions and corresponding emotions [85] made way for first ground-breaking work in 1996 in which [15O]-water PET was combined with the presentation of fearful versus happy faces yielding significant rCBF increase in the left amygdala [86]. A few years later, these findings were reproduced with fMRI [87] and the same study also showed longitudinal reduction of amygdala activation as a treatment effect of the antidepressant sertraline after eight weeks of therapy. A research group in the UK focused their efforts on task-based fMRI (tfMRI) of short-term effects of antidepressants with emotional faces for several years. In single fMRI acquisitions, they could show that two to three hours after a single dose of citalopram or mirtazapine, amygdala activity was reduced in healthy volunteers in response to fearful faces [88,89]. Later, they reported that escitalopram (versus placebo) had reduced amygdala hyperactivity in depressed patients after seven days [90]. These studies, however, did neither compare medications directly nor employ longitudinal fMRI data acquisition during treatment periods. There are different variations of tfMRI with negative emotional faces. The general paradigm, though, is considered robust as shown in a metaanalysis of 105 fMRI studies [91]. As an example, the Hariri variant of this paradigm [92] was included as the core tfMRI acquisition in the imaging extension of the UK Biobank [93] with one hundred thousand participants.

Resting-state fMRI (rs-fMRI or rfMRI) provides the basis for investigating functional connectivity (FC). It is understood as statistical dependencies between remote neurophysiological events within the brain and, thus, continues to address the concept of functional segregation [94]. In clinical depression, there appear to be several common patterns of change in functional connectivity like increased connectivity between the salience network and the anterior default mode network (DMN) [95] and these changes may even identify subtypes of clinical depression [96].

A relevant limitation of FC is that it can only provide for averages of complex neurophysiological events over space and time. Hence, striving to quantify changes in FC measures over time has led to dynamic functional connectivity (DFC) [97]. Within the last few years, investigating DFC in clinical depression has revealed some first insights including how DFC characteristics of certain brain regions may relate to specific phenomenological traits of clinical depression [98].

Water diffusion MRI (i.e. diffusion tensor imaging, DTI) enables fibre tracking (i.e. tractography) in the brain [99] and, thus, provides the basis for investigating structural connectivity (SC). It has already proven to be an interesting source for additional biomarkers for depression in itself, like reduced global fractional anisotropy (FA) in acutely affected patients with depression [100].

Combining structural and functional connectivity leads to the human connectome [101]. It not only helps to mutually validate findings from the other modality [102] and to establish links between both [103]. It also provides additional value beyond the sum of its parts of human brain function [104]. Understandably, both MRI modalities are also part of large imaging endeavors like the UK Biobank imaging extension.

#### Antidepressants

Antidepressants remain a first-line recommendation for the treatment of adult patients suffering from moderate to severe clinical depression. This said, there is growing controversy on the efficacy of antidepressants versus placebo [8,9,105,106] which begins to reflect in recent updates of national guidelines for treatment of depression. This trial, however, does not aim at judging on treatment efficacies, but focusses on the exploration of differential effects of three fundamentally different pharmacological agents, escitalopram, mirtazapine and agomelatine.

Escitalopram is the *S*-enantiomer of the racemat citalopram [107]. It is a highly selective serotonin reuptake inhibitor (SSRI) [108] which makes it a potent representative [109] of the largest class of second-generation antidepressants.

Mirtazapine, a 6-aza analogue [110] of mianserin [111], is primarily known for its distinct dual mode of action by increasing serotonergic and noradrenergic transmission independent of serotonin and norepinephrine reuptake blockade. It primarily does so by antagonizing at central presynaptic alpha-2 receptors [112]. Furthermore, it antagonizes at serotonergic 5-HT2 and 5-HT3 receptors (5-hydroxytryptamine, 5-HT) as well as histaminergic H1 receptors. Its modes of action make it a so-called atypical antidepressant [113] with different pharmacodynamics than the substances that can be found in the groups of selective serotonin reuptake inhibitors (SSRIs), serotonin–norepinephrine reuptake inhibitors (SNRIs), tricyclic antidepressants (TCAs) or monoamine oxidase inhibitors (MAOIs).

Agomelatine, a naphthalenic analogue of melatonin [114], is an agonist at melatoninergic MT1 and MT2 receptors and an antagonist at 5-HT2C receptors. Its melatonergic transmission originally let it be in the focus as a chronobiotic [115] until its antidepressant effects were revealed [116,117]. Thus, agomelatine also belongs to the small group of atypical antidepressants.

Therapeutic efficacies of all three agents are in similar ranges. A large network meta-analysis including 116477 participants [118] yielded odds ratios (OR) of 1.65 (agomelatine), 1.69 (escitalopram) and 1.89 (mirtazapine) and head-to-head comparisons between these three resulted in ORs of 0.90 and 0.93 for agomelatine versus escitalopram and mirtazapine and 1.04 for escitalopram versus mirtazapine.

While there is no doubt in the results of medicinal chemistry and pharmacological in vitro and ex vivo investigations which group these agents into mechanistic classes, simplistic neurotransmitter models like the serotonin and monoamine imbalance theories of depression, that have been a foundation for the development of antidepressants, have become increasingly less valid [119,120]. This opens up new and broader perspectives on much more complex potential mechanisms of action of these substances. For example, discovery and development of antidepressants is closely linked to their antimicrobial beginnings. The first antidepressant and monoamine-oxidase inhibitor originally was a tuberculostatic agent [121] and contemporary investigations confirmed the remaining antimicrobial activity of antidepressants in vitro [122] and in vivo [123]. Hence, some of the many potential reasons for a comparatively long onset of antidepressant efficacy may be rooted in direct modulation of the gut microbiome or in unknown indirect effects via the enteric nervous system along the gut-brain axis.

### OptiMD as framework for this trial

This trial is embedded in the German research consortium OptiMD (*Novel Strategies for Optimized Treatment of Depression*) which aims to contribute to a better understanding of antidepressant efficacy and to identification of potential response biomarkers as well as to investigate novel treatment strategies for depression [124]. OptiMD has been one of nine large multi-center research consortia funded by the *Bundesministerium für Bildung und Forschung* (BMBF, German Federal Ministry of Education and Research) as part of the national program *Forschungsnetz für psychische Erkrankungen* (FZPE, Research Network for Mental Disorders, [125]). Investigational efforts within OptiMD have been focused on six major branches (called ‘subprojects’ = SP). Gut microbiome (SP3) and MR neuroimaging (SP5) are two of those subprojects that are largely represented in this trial. The consortium comprises seven study centers encompassing nine recruiting university hospitals across Germany (Berlin (SP6), Erlangen (SP2), Essen (SP2), Heidelberg (SP5), Munich (MPI, SP4) and Regensburg (SP1 and SP3), later extended by Aachen, Frankfurt am Main, and Munich (LMU) to support recruitment).

### Objectives

#### Motivation and overview

The central goal of this trial may be introduced with the following question: Is it possible to better understand the neurobiology of clinical depression by simultaneously and longitudinally investigating multiple levels of human physiology with state-of-the-art non-invasive technologies during the first week of antidepressant treatment within the same individuals, i.e. early pre/post deep-phenotyping?

This question is relevant for at least three reasons. First, clinically perceivable therapeutic effects of antidepressants usually only reveal themselves after ten to fourteen days at the earliest, making the first two weeks of pharmacological treatment a clinical blind flight in this respect. Second, a selected antidepressant may not work sufficiently or not at all which then increases the burden of suffering for patients over time. Until today, unfortunately there is no way to predict the efficacy of an antidepressant when prescribing it. Third, clinical depression may not be a single disease entity but a heterogeneous clinical appearance of various underlying pathologies. This implies that patients affected by what we currently call clinical depression as a single entity might need to be assigned to specific subgroups of depression in order to optimize therapy efficacy. The first two aspects mentioned above might be related to this heterogeneity to a certain degree.

Exploring (bio-)technology-based ways to differentiate patients with depression within the first days after diagnosis or through their initial reactions to an antidepressant is related to hopes for a better understanding of disease pathology and higher therapy efficacy in the future.

#### Considerations on objective definitions

Our objectives for this study were not primarily set towards predictive analytics as we intended to be realistic about our goals in relation to achievable sample sizes in regard to the large efforts involved for this trial design. This reflects in the structure of our primary, secondary and exploratory aims. While we intended to at least confirm certain predictive traits of published biomarkers along our secondary objectives, we also hoped that the enormous speed in the development of contemporary statistical methods would assist in gaining further insight into our deep phenotyping dataset later on, as reflected in our exploratory aims.

#### Primary aim

The nature of this study is exploratory and hence, the primary objective of this clinical trial is to explore the differential effects of escitalopram, mirtazapine and agomelatine and placebo in a longitudinal pre-/post-design in which multiple research modalities were performed before and after seven days of double-blind randomized medication (and partly at end of trial) in the same participants. The original focus was on three of the five major modalities reported here: MR neuroimaging, combined dexamethasone-CRH-challenge testing and gut microbiome analyses.

#### Secondary aims

Secondary aims address some predictive analytics in regard to HPA reactivity and MR neuroimaging and descriptive analytics in regard to gut microbiome. They partly address how pre/post-differences as described in primary objectives relate to therapeutic outcome.

The predictive value of HPA activity should be looked at from two perspectives: time course of clinical response as well as overall clinical response behavior depending on reduction of HPA reactivity by the first week of therapy. In other words, do patients with reduction of HPA reactivity by the first week of treatment also show an earlier clinical response during the course of therapy and does a reduction of HPA reactivity by the first week generally indicate higher response rates towards the end of the trial period?

Similarly, the potential predictive value of various MR neuroimaging measures was another secondary objective. Are certain changes in pre/post-MR measures around the first week of pharmacologic therapy related to clinical response behavior and might they be of interest for future investigations as potential predictive biomarkers? Furthermore, does HPA reactivity associate to these measures?

Targeting predictive markers in gut microbiome was too far-fetched at the time of conception of this trial. Therefore, secondary objectives in regard to gut microbiome were kept conservative. Identifying changes in the bacterial gut microbiome that associate with therapy response and might therefore show potential as biomarkers in future studies was a prudent secondary aim.

All of the secondary aims relate to therapeutic response dynamics over an observational period of eight weeks and require the originally randomized antidepressant to be steadily continued as monotherapy after the first double-blinded week with accepted dose elevations from the third week onwards according to the study protocol.

#### Exploratory aims

Exploratory aims address whether it is possible to find links between stress hyperreactivity, inflammation, gut microbiome and brain function in affected patients. Hence, exploratory aims include further exploration into differential effects of escitalopram, mirtazapine and agomelatine versus placebo on extensive sets of longitudinal lipidome and neuroinflammatory proteome markers. Furthermore, they include the exploration of possible associations between all of the investigated modalities depending on available statistical methodology as mentioned above. One of several interesting exploratory aims would be a discovery of links between stress hyperreactivity, inflammation, gut microbiome and brain function in clinical depression.

## Material and Methods

### Ethical standards

The trial complied with the *Declaration of Helsinki* from 2013 [126] and, in retrospect, also conformed with its revision from 2024 [127]. Furthermore, the trial complied with the *Guideline for Good Clinical Practice* (E6(R1), [128]) of the International Council for Harmonisation of Technical Requirements for Pharmaceuticals for Human Use (ICH), as well as with the *Arzneimittelgesetz* in Germany (AMG, i.e. German Medicinal Products Act).

Study protocol, patient information sheets, and informed consent forms were approved on 20 May 2015 by the *Ethikkommission bei der Universität Regensburg* (institutional ethics committee, Regensburg, Germany) and given approval number 15-111-0075. The trial was also registered with and approved by local district authorities and conducted in accordance with local legislation and institutional requirements. All participants provided their written informed consent to participate in this study. A first amendment was approved on 25 May 2016 under the same approval number. A second amendment was approved on 25 October 2017 and given approval number 17-758-101. Details are described in section “Trial status”.

Before, the trial had already been registered with the European Union Clinical Trials Register on 30 July 2013 (EudraCT number: 2013-003370-27) and had received approval by *Bundesinstitut für Arzneimittel und Medizinprodukte* (BfArM, Federal Institute for Drugs and Medical Devices, Prüfplancode (identifier): DMFMRI201303) according to § 42, para 2 of German Medicinal Products Act (*Arzneimittelgesetz*, AMG) on 25 September 2014.

This study protocol follows the SPIRIT guideline and CONSORT statement and other guidelines as described in more detail in section “Conformance statement for reporting guidelines and recommendations”.

### Human subjects, screening and recruitment

The trial was conducted at the Department of Psychiatry and Psychotherapy (Universität Regensburg, Regensburg, Germany) from January 2016 to August 2021.

It was one of seven clinical trials focused on clinical depression conducted in parallel at our center and one of four clinical investigations as part of the German research consortium OptiMD as well as from one from an FZPE consortia-encompassing project (PD-CAN) in all of which our study center participated. One of the seven trials was an investigator initiated single-center trial to assess inflammation and cardiovascular risk in clinical depression that had already been recruiting for more than three years at our study center. Similarly, another one of the seven trials was an investigator initiated single-center trial for reprogramming fibroblasts of human patients with depression into neuronal precursor cells that had been recruiting for about one year at our study center at the time of study initiation. Both trials were additionally served with co-enrollment from this trial as explained in “Axis VII: Co-enrollment in other clinical depression trials”.

All of the four clinical OptiMD investigations were multi-center trials with lead centers in Regensburg (SP3, gut microbiome), Munich (SP4, genome-wide association), Heidelberg (SP5, neuroimaging) and Berlin (SP6, minocycline augmentation). All of these were initiated in Regensburg between January and May 2016. A cross-sectional FZPE investigation with lead center in Potsdam (PD-CAN, Domain Assessment Network Germany) became part of OptiMD-related recruitment and co-enrollment for this trial.

The trial described here, was an exception within OptiMD in that it was primarily designed to be a single-center exploratory deep phenotyping investigation. However, it was well suited and appropriate to fully embed the multi-center projects SP3 and SP5 because of strong conceptual overlaps.

Hence, recruitment efforts at our center were managed in a multistage process guided by the rigor of required eligibility criteria for the different clinical investigations running in parallel. Criteria for the trial described here, were the strictest in regard to drug-naïve inpatients admitted for depression. Hence, this trial was usually the first one to be considered for recruitment in this group of admitted patients.

During the recruitment period of this trial, 669 human subjects affected by clinical depression were screened (328 female, 322 male, 19 with undocumented gender) of whom 189 inpatients and outpatients (83 female, 106 male) were recruited for the five OptiMD-related projects described above. Of these, 70 inpatients were included in this trial (24 female, 46 male). Seven of these seventy participants dropped out, six of them before randomization and one of them during the first week of treatment. Reasons for dropping out were: change of mind in regard to study participation, wish for being discharged from hospital even before randomization, emerging concerns in regard to the risk of being randomized into a seven-day placebo group, one screen-failure, one incipient exclusion criterion in regard to novel deviations in electrolyte laboratory values as well as one decision by discretion of a senior attending psychiatrist to exclude placebo risk and begin open-label treatment. Another participant was not randomized to study medication because the second MRI scan (“post”) was not possible due to retrofitting of a new MR research tomograph.

Towards the end of clinical recruiting, the trial also recruited healthy women and men as an additional control group over a period of four months (November 2020 to February 2021). Altogether, 90 human subjects (58 female, 32 male) had originally shown interest in participating in the trial. Eight months into the COVID-19 pandemic, 65 of these (43 female, 22 male) were still available for screening of whom 25 age- and gender matched healthy volunteers were included in the trial (10 female, 15 male).

### Eligibility criteria

#### Non-naturalistic selection of patient cohort

In our trial ideality, the goal of initial clinical assessments had been very strict and meant to identify inpatients who preferentially were admitted to the hospital with a drug-naïve intermediate to severe first-episode of clinical depression without any psychiatric or somatic comorbidities and expressing no to low or at least well manageable levels of suicidal contemplations. In reality, inclusion and exclusion criteria had been defined in a comparably open manner to enable a reasonable recruitment process while the ideal mentioned before was continuously emphasized and aspired by core trial staff. Thus, patient selection according to this ideal was achieved for a large part of participants. Hence, in comparison to typical psychiatric inpatient admissions, this filtering of potential study participants was far from being naturalistic and it required dedicated and extended time resources for study physicians. It is possible that the distribution of age and gender in the patient cohort relates to this phenomenon.

#### Patients

Eligibility criteria for patients are shown in detail in Table 1. The valid diagnostic manual in Germany has been ICD-10 while some study doctors involved were also skilled in applying corresponding DSM-5 criteria for the diagnosis of depression. Key inclusion criteria were hospital admission because of a primary diagnosis of clinical depression according to ICD-10 (German Modification, Version 2015 [129,130]) without indications of other underlying psychiatric illnesses, an age between 18 and 65, physical health, absence of psychotropic medication for at least three weeks before admission and the ability to provide written informed consent. Severity of depression was implied by hospital admission which related to no less than to a moderate to severe level of disease. Key exclusion criteria were major psychiatric diagnoses beyond depression including the abuse of alcohol or drugs within the last six months, general medical conditions or neurological disorders with particular regard to local or systemic inflammatory processes, uncommon dietary habits, allergies or intolerances to intake of escitalopram, mirtazapine or agomelatine, non-removable non-MR-safe metals and implanted stimulators as well as pregnant or nursing women.

**Table 1:** Inclusion, exclusion and discontinuation criteria.

**Figure 1: Schedule of interventions and assessments (SPIRIT 2025-conformant timeline)**
|  |
| --- |
| **Inclusion criteria**<br>• age 18 - 65 years<br>• inpatient psychiatric treatment<br>• diagnosis of clinical depression with a minimum duration of two weeks [1]<br>• indication for antidepressant therapy is independent of trial participation<br>• voluntary hospital admission independent of trial participation<br>• capability to understand the scope, details and consequences of trial participation<br>• willingness and capability to provide written informed consent<br>• physical health<br>• right-handedness<br>• eligibility for magnetic resonance imaging based on MR safety criteria<br>• female patients of childbearing age: negative blood pregnancy test and willingness to use a reliable method of birth control during trial |
| **Exclusion criteria**<br>• involuntary psychiatric admission<br>• schizophrenia, addiction, eating disorder or any other kind of psychiatric disorder as primary diagnosis<br>• abuse of alcohol or drugs within the last six months<br>• lacking capability to understand the scope, details and consequences of trial participation<br>• increased risk for suicidal behavior or endangerment to self or others [2]<br>• treatment with psychotropic medication within the last three weeks (five weeks for fluoxetine)<br>• treatment with antibiotics or corticosteroids<br>• contraindications, known allergies or intolerances to intake of escitalopram, mirtazapine or agomelatine<br>• somatic or neurological disorders [3]<br>• abnormal blood parameters with clinical relevance [4]<br>• uncommon dietary habits which may also lead to malnutrition<br>• non MR-safe metals that cannot be safely removed and implanted stimulators or pumps<br>• undeflectable agoraphobic experiences during imaging<br>• pregnancy or ongoing breast-feeding |
| **Discontinuation criteria**<br>• participants may withdraw voluntarily from the trial at any time and without any specific reasons [5]<br>• worsening of psychiatric condition [6]<br>• emergence of active suicidal behavior [2]<br>• safety concerns, for example because of adverse effects [7]<br>• serious non-compliance to study protocol [7]<br>• incorrect inclusion in trial (i.e. lacking fulfillment of inclusion criteria or presence of exclusion criteria)<br>• novel emergence of exclusion criteria, for example pregnancy<br>• Neutropenia control during antidepressant treatment:<br>if neutrophil granulocytes show a lower concentration than $1.0 \times 10^9/l$, blood lab must be repeated within 24 hours<br>if repetition confirms low neutrophil count, the following steps need to be followed:<br>◦ participant needs to be excluded from trial and lab finding need to be documented at least as adverse event<br>◦ complete differential blood counts need to be performed weekly until normalization<br>◦ during ongoing neutropenia doctors and patients need to comply to common neutropenia safety measures (for example avoiding invasive procedures, avoiding people with obvious signs of infection)<br>◦ during ongoing neutropenia and development of fever or other symptoms implying the occurrence of novel infectious processes, patients need to get in touch with the attending psychiatrist and a complete differential blood count should be taken, immediately |

[1] depressive episode in bipolar disorder would have been allowed but was not included in trial
[2] as partly reflected in HAMD item no. 3 with three or more points
[3] with particular regard to systemic inflammatory diseases and gastrointestinal passage
[4] Definition of abnormality in laboratory values: either exceeding twice the upper limit or falling below half the lower limit of normal reference values
[5] Participants who withdrew from the study should undergo a dedicated examination by the investigator. They should be asked for possible reasons of discontinuation and the presence of side effects. Unexpected or severe adverse events or side effects should be followed up.
[6] as reflected in a 25%-increase of HAMD21 sum values relative to baseline HAMD21-measurement
[7] by discretion of attending psychiatrist

#### Healthy subjects

Healthy volunteers interested in study participation were first asked to provide written answers to a specifically designed one-page screening questionnaire followed by a screening phone or video conversation. Major points of interest in the questionnaire and later on were presence of depressive mood states or other potentially psychiatric phenomena, character of use of alcohol, tobacco and other drugs, type of diet, intake of nutritional supplements or any kind of medication, in particular the use of antibiotics within the last six months and willing or unwilling fluctuations in body weight. Further key criteria were major or minor surgical interventions or invasive procedures like gastroscopy or colonoscopy within the last six months, signs of gluten sensitivity, irritable bowel syndrome or other irregular bowel behavior, previous difficulties during possible examinations with MRI, CT or PET with special focus on possible signs of agoraphobia and presence of non-removable non-MR-safe metals.

Later on, mental health was formally screened with the MINI International Neuropsychiatric Interview [131] and continued detailed history taking. Similarly, physical health was verified with detailed medical history taking, physical examination, blood work including pregnancy testing and measurement of vital signs including ECG. Possible illegal drug usage was assessed by urine testing.

The complete recruitment and study period for healthy participants was coinciding with the second wave of the COVID-19 pandemic in winter of 2020/2021. This required additional screening for any signs of previous or current possible infection and, later on, for vaccination status and possible side effects by vaccination.

For reasons of personnel safety and study quality, all healthy participants were PCR-tested for SARS-CoV-2 in the morning of planned investigations without any positive results in participants or staff throughout. One participant had already received the first SARS-CoV-2 vaccination with an mRNA agent (with no perceivable side effects) while another had already received the second (with no perceivable side effects during the first and minor side effects during the second). In these cases, study investigations were scheduled at least two to three weeks after vaccination.

### Blinding and randomization

Randomization and blinding of study medication was performed by the pharmacy of LMU University Hospital Munich (affiliated to Ludwig-Maximilians-Universität (LMU), Munich, Germany). White hard gelatine capsules (iphas Pharma-Verpackung, Würselen, Germany), size 0, containing 2 % titanium dioxide (E171) [132] were filled using a Feton capsule filling machine (Feton International, La Louvière, Belgium) with one of three licensed antidepressants and a prepared mixture of mannitol Ph. Eur. grade (E421), highly dispersed silicon dioxide Ph. Eur. grade (E551) and red iron oxide (E172) for verum capsules or just the prepared mixture without active substance for placebo capsules. At the time, a randomization list was generated using a Web-based pseudorandom number generator (www.randomization.com) provided openly by Gerard Dallal from Tufts University [133]. The randomization plan was created on 19 October 2015 by randomizing 80 subjects into two blocks with 40 patients each. Seed number ‘11195’ would have reproduced the same plan.

Study dosage was one capsule in the morning and one capsule in the evening for the first seven days of antidepressant treatment for all participating patients. One of the two daily capsules was identical to placebo capsules, i.e. without active substance. Escitalopram 10 mg (Lundbeck, Hamburg, Germany) was given in morning capsules while mirtazapine 30 mg (AbZ Pharma, Ulm, Germany) or agomelatine 25 mg (Servier Deutschland, Munich, Germany) were given with evening capsules. All four parallel trial groups were balanced and distributed equally such that the chance of receiving placebo during the first seven days was 25 %. Assignment of study medication was masked to participants and all members of personnel including prescribing study doctors, study nurses, ward staff and doctors on call.

### Trial safety

#### Principles of clinical etiquette and authentic informed consent during clinical depression

Some core principles were generally guiding the members of the core trial team in interactions with patients who had expressed basic interest in study participation or who had already agreed to participate. These bedside manners and values were kept up during the full trial duration and also in regard to obtaining authentic informed consent in the beginning. This etiquette during study-related interactions included letting the patient guide the flow of information and interaction, taking time and taking it slow, describing all aspects of study participation with radical transparency, repeating information until full satisfaction of patient, adapting to diurnal variations of depressed mood, motivating interested patients to seek exchange with other study participants on ward, encouraging the consulting of friends and family and offering visits to and dry-runs at sites of investigation, for example, visiting the MRI facilities and being placed in MR bore without measurements.

#### Medical clearance

A routine process of medical clearance is applied to all psychiatric patients who are admitted to our hospital for the first time. It usually includes somatic history taking, full medical and neurological examination, ECG, routine brain MRI for first admissions and an initial set of several routine laboratory values. This process was slightly adapted and extended for study participants. As described below, clinical routine head and brain MRI was implemented into research imaging sessions to lessen the load on study patients in regard to diagnostic procedures. History taking was well extended and partly included retrieval of previous medical records including possible previous neurological images. On the research ward, further described in the next section, initial routine laboratory evaluation has generally been more comprehensive for all patients admitted there. Furthermore, a smaller routine set of lab values has usually been repeated each week independent of study participation. Retrieval of previous medical records and exchange with previously and currently treating doctors extended to simple metallic implants as, for example, orthodontic retainers in cases when patients could not provide sufficient information on MR-safety of their implants.

#### General patient safety

The Department of Psychiatry and Psychotherapy had furnished a ward specialized in conducting clinical research with depressed inpatients. This included a higher staffing ratio, specifically trained personnel and continuous presence of at least one study nurse and one study doctor during daytime hours. Furthermore, study doctors were given dedicated resources to allow for sufficient time in interactions with study patients. Open doors for patient inquiries and at least once daily and sometimes multiple daily interactions with study patients ensured sufficient knowledge about their current condition and requirements, particularly in regard to possible strain caused by study measures or control of adverse effects by medication. Moreover, all study patients were offered supportive conversations immediately after trial start with a trained psychologist twice a week during the first two weeks of study participation in addition to the usual set of therapeutic components being initiated with admission. The same psychologist continued common weekly psychotherapy with these study patients on ward afterwards. This early supportive measure was installed for all participating patients to address a 25 % chance to receive placebo during the first seven days of medication.

The course of the trial showed that study patients usually appreciated the change, diversion and increased supervision related to study participation during the first two weeks after hospital admission – a time during which pharmaceutical and psychological interventions could mostly not yet show their therapeutic effects.

#### Routine blood markers during trial

All patients treated on the research ward described above receive weekly routine blood testing which usually comprises 19 basic medical screening parameters aiding in medical clearance and monitoring of possible pharmaceutical side effects: creatinine, urea, sodium, potassium, calcium, γ-GT, AST (GOT), ALT (GPT), creatine kinase, glomerular filtration rate (GFR), C-reactive protein (CRP), leukocytes, erythrocytes, hemoglobin, hematocrit, MCV, MCH, MCHC, and thrombocytes.

These routine parameters have been extended for patients who had decided to participate in one of the larger clinical studies on this ward. This extended set of 44 blood parameters was taken at the beginning and end of a clinical study to further ensure patient safety and to support the initial screening process. The extended blood routine laboratory adds another 25 parameters for men and 26 for women: ureic acid, cholesterol, triglycerides, HDL, LDL, bilirubin total, amylase, lipase, iron, ferritin, transferrin, transferrin saturation, total protein, neutrophils, lymphocytes, monocytes, basophils, eosinophils, HbA1c, Quick, INR, PTT, TSH, free triiodothyronine (fT3), free thyroxine (fT4) and β-HCG (i.e. serum human chorionic gonadotropin, for women of childbearing potential, on first day of study).

Furthermore, a standard analytical urine drug screen at the beginning of a clinical trial searched for four drug classes: amphetamines, benzodiazepines, cannabis and opiates. In cases of relevant deviations in creatinine concentration in urine, measured drug values were corrected accordingly.

#### Pharmaceutic safety

Continuous training and regularly providing study-specific information to all personnel on ward and to all doctors who might be on call after regular hours was one of several measures for study patient safety. This was supplemented by specific trial-related documentation in the integrated hospital information system for each study patient including contact information of trial staff and details on possible proceedings.

Sealed emergency envelopes with unblinding information for the respective drug numbers had also been prepared by the pharmacy of the LMU university hospital and were stored in a dedicated locked cabinet on the ward where study patients were treated. All nurses of this ward were trained to provide information and access to those envelopes for doctors on call upon request at all times. Furthermore, all potential doctors on call were regularly informed on the criteria, conditions and proceedings related to this and all ongoing trials, for example during central morning briefings. In case clinical condition of a patient allowed for it, doctors on call where asked to first call the study doctor primarily responsible for the patient before making changes to medication or opening emergency envelopes. During the whole period of the trial, the need for pharmaceutic changes did not occur outside regular daytime hours and an emergency envelope only needed to be opened once on request by a senior attending psychiatrist who excluded a patient on the fifth day of study medication to exclude risk of placebo treatment.

#### Adverse events

Long established, commonly used and ready-made first-line antidepressants escitalopram, mirtazapine and agomelatine were used as study medications in this trial in a monotherapy protocol along their main indication and as part of national treatment guidelines. However, blinding of study medication during the first seven days and a chance of placebo required additional and strict conformity to the German Medicinal Products Act (*Arzneimittelgesetz*, AMG) and related regular reporting to the German Federal Institute for Drugs and Medical Devices (*Bundesinstitut für Arzneimittel und Medizinprodukte*, BfArM) including the reporting of possibly occurred severe adverse events (SAE).

Adverse events (AE) were controlled on a near daily basis (assessment on weekend days only in part) and recorded from informed consent to after the end of the clinical trial depending on severity and time of occurrence. During the whole period of the trial, a single SAE occurred which was assessed to be unlikely related to study medication: four weeks after entering the trial and having been randomized into placebo group, subsequently receiving agomelatine 25 mg and after two weeks of administration of 50 mg according to study protocol, the patient began reporting intermittent malaise and fluctuating but slowly increasing diarrhea. After two weeks of conservative but increasingly insufficient therapy, the patient was transferred to a gastroenterology department where a pancolitis was observed and classified as a likely symptom of colitis ulcerosa. The patient was therefore immediately treated with antibiotics and cortisone. According to the patient and treating doctors, it was a first-time diagnosis. After sufficient observation in gastroenterology, the patient was transferred back to our department. Gastroenterologists did not see therapy with agomelatine 50 mg as a source of the pathology and therefore continued to prescribe it in their department, as well. After transfer back to us, it was continued at a stable dose and without need for additional psychiatric medication until final discharge without changes in medication.

#### MR safety

MR safety was a major consideration during this trial. The majority of MR scanning was done in a newly installed 3 Tesla MR tomograph dedicated for research. Strict user regulations were established and required special training and certification for everyone intending to participate in MR scanning operation. Hence, operator skills were evolving through different certified levels of expertise, maturity and rank. Therefore, an MR operator team needed to be composed of a so-called ‘level-1’ and a so-called ‘level-2’ operator with a pre-defined distribution of roles and responsibilities and a corresponding set of standard operating procedures. Furthermore, a senior supervising radiology technician was continuously available for solving technical errors in radiological routine at all times during research scanning.

An administrative workflow was established to manage incidental findings that also might have posed (previously unknown) contraindications for trial continuation. Abnormalities noticed by the research MR operating team in clinical scans were immediately to be discussed on the phone with the head of the neuroradiology department. Vice versa, the neuroradiological department prioritized the assessment of clinical scans that were acquired first, such that possible contraindications for trial continuation could immediately be communicated. Only a single neuroradiological abnormality has been detected in one of the 89 participants for MR scanning: it was a small cyst located in the pineal gland in one of the patients presenting without any clinical symptoms. It did not pose a contraindication for trial continuation as assessed by our neuroradiology department. The patient was asked to return as outpatient for a neuroradiological follow-up six months after.

A study nurse that usually knew the patient from study administration was escorting the patient from ward to MR facility and remaining in direct proximity in the MR control room for the whole duration of measurements after which the study nurse escorted the patient back to ward. This ensured continuous observation of the patient. A study doctor was either on call and available within three minutes by discretion of the study nurse or MR operator team. Alternatively, one study doctor was participating in many of the measurements as a certified MR operator with frequent and direct interactions with the patients and healthy participants.

Detailed requirements for MR safety were assessed by a study doctor during the verification of inclusion and exclusion criteria in the very beginning. This process was repeated with three different screening questionnaires at different time-points asking for non-removable metals, implanted valves, stimulators and pumps in different ways. This multistep process owed to different projects being combined and additionally provided for safety by redundancy.

#### Infection control during COVID-19 pandemic

Safety of study participants during the COVID-19 pandemic was of particular concern besides caring for the general safety of patients while changes in general infection control guidance required continuous adaptation. Therefore, on our campus, non-COVID-related clinical research was shut down completely for four months until July 2020. After that, reinitiating clinical research projects was challenging on various levels and hence, slow. Three patients were included in this final recruitment phase from then on. On the other hand, all of the 25 healthy subjects were completely recruited and participating during the second wave of the pandemic from November 2020 to February 2021. For our clinical trial, infection control was divided into three areas of research activity: first, for inpatients on ward, second, for healthy control subjects and, third, for all participants entering the MR research facility.

Besides general infection control measures, all staff and participants were continuously wearing FFP2-masks throughout with an exception for MR scanning as described below. Furthermore, all staff was monitoring themselves for possible signs of infection and performing daily antigen tests in the morning before starting work and additionally, before interacting with a different study participant. Also, research staff members were eligible for low-threshold, immediate PCR-testing in case they did not feel safe about perceived symptoms or possible contacts. Correspondingly, all participants were informed in advance about self-monitoring practices and asked to immediately report possible signs of infection by phone or to a nurse on ward. All members of the core trial team, and in particular study nurses, were specifically trained in all procedures and agents for disinfecting materials, surfaces and cleaning research spaces. Moreover, all rooms and spaces were frequently vented, either by keeping windows continuously open when outside temperatures allowed or by rigorously venting at least every 30 min. Whenever study procedures allowed for it, a minimum distance of 1.5 m was kept up during interactions. Beyond that, work roles were split as described below.

A core idea for additional safety of study participants was the separation of research and clinical spaces, roles of involved team members, workflows and participants, including the full separation of healthy participants from patients. Thus, for example, study doctors who were in charge for study patients on ward were not interacting with healthy participants. Additionally, a separate building was retrofitted to become a dedicated research environment for healthy participants, only. Furthermore, regulations for using the research MR facility were adapted to account for spatial separation of workflows as well as separation of operator roles such that only a single operator had contact with a participant with a distance below 1.5 m and only this operator was allowed to enter the MR room, itself. All participants were required to wear their FFP2-mask until the MR coil was placed above their head as a last step before driving their bed into the MR bore, such that scanning procedures themselves were performed without masks on participants. Correspondingly, they were again provided with a mask immediately after exiting the MR bore.

This comprehensive set of infection control measures appeared to work well, as to our knowledge, there did not occur any SARS-CoV-2 infections among study patients, healthy participants or research staff members during the whole trial period and follow-ups.

#### Trial oversight, monitoring and sponsorship

This study has been designed as a single-center trial coordinated by the Department of Psychiatry and Psychotherapy, Universität Regensburg, Regensburg, Germany. During recruitment phase, a clinician scientist, two study assistants and two psychiatry residents, all of them trained to conform to Good Clinical Practice (GCP), formed the core of the clinical research team for this trial. This team and the principal clinical investigator met once of week, together with other participating staff from the ward where the trial was conducted. These meetings served, among others, to discuss the well-being of study participants, state of recruitment, trial quality control and possible improvements in workflows.

Trial monitoring was assigned to a senior attending psychiatrist from a neighboring ward with dedicated experience in performing clinical trials as a principal investigator himself. There has neither been a dedicated trial steering committee nor a public involvement group for this trial and no further stakeholders than the ones mentioned above.

The trial was sponsored by the municipal administration of the neuroscience campus were the Department of Psychiatry is located, i.e. by medbo KU, a short name for **Med**izinische Einrichtungen des **B**ezirks **O**berpfalz – **K**ommunal**U**nternehmen (Anstalt des öffentlichen Rechts) at Universitätsstrasse 84, 93053 Regensburg, Germany. The German institution name translates to ‘medical facilities of the Upper Palatinate district in Bavaria’, Germany – a municipally owned enterprise governed by public law and represented by chairman of the executive board, Helmut Hausner. It comprises multiple medical facilities spread across the whole district, with Regensburg campus being the largest, offering 530 beds for psychiatric inpatients, and several other neuroscience-related disciplines. For better understanding of sponsor affiliation, academic clinical infrastructure in Regensburg is unique in that several departments of the university’s medical faculty are not located at the university hospital itself but decentralized on other clinical campuses of the same city. Thus, medbo Regensburg campus exclusively houses the majority of academic neuroscience departments like psychiatry, neuroradiology, neurology, pediatric psychiatry and even forensic psychiatry departments for adults and children.

#### Insurance policy for all participants

All clinical trials that need to be registered according to the German Medicinal Products Act (*Arzneimittelgesetz*, AMG) require formal insurance for all participants. For this, HDI-Gerling Industrie Versicherung AG (HDI = Haftpflichtverband der Deutschen Industrie), now HDI Global SE, was contracted via our *Zentrum für Klinische Studien* (Center for Clinical Studies) at Universitätsklinik Regensburg to provide insurance on a year-wise basis for all participating study patients and healthy volunteers (insurance policy number: 57 010314 03010). Maximum coverage for each participant was set to half a million Euro and total coverage for the whole project was 50 million Euro. There have been no insurance claims at any point until today.

### Experimental Design

#### Overview of trial design and time-course

The design principle of this single-center trial is a technology-focused pre/post deep-phenotyping approach around the first week of antidepressant medication within the same drug-naïve inpatients who needed to be admitted to a psychiatric ward because of clinical depression, most of them for the first time in their lives. Here, “pre” denotes a baseline before initiation of antidepressant medication and “post” the end of the first week of medication. Additionally, some investigations were repeated for a third time at the end of the trial period which was set to 60 days in order to be able assess clinical trajectories of participants – corresponding to exactly eight weeks of antidepressant medication per protocol.

The pharmacological trial environment has been structured and controlled in four balanced treatment arms during the first seven days of medication by double-blinding it and randomizing participants to receive one of three commonly prescribed antidepressants – escitalopram 10 mg, mirtazapine 30 mg, agomelatine 25 mg – or placebo. These antidepressants were chosen for their clearly distinguishable pharmacodynamics from each other, in particular in regard to their distinct effects on the HPA-axis, while all of them are valid first-line treatments for clinical depression.

One quarter of randomized participants would receive placebo within the first seven days of treatment. Besides many other trial safety procedures described in section “Trial safety”, risk of placebo was addressed by initiating specific psychotherapeutic support from the very beginning of the trial over two weeks for all participants. It was transformed into regular psychotherapy from the third week onwards, similar to the psychotherapeutic regimen of patients in a usual clinical inpatient setting.

Medication was unblinded on medication day eight (i.e. study day twelve). From that day on, placebo-patients continued with open-label agomelatine 25 mg, thus merging two of the four arms with a time lag of seven days. A structured scheme of optional increases in dosage over the course of treatment after two weeks of stable initial dose had been prepared in the protocol and were decided by discretion of the senior attending present at the time.

While the investigational focus of this trial has been on the first eleven days, the duration of the whole trial has been set to 60 days in order to encompass exactly eight weeks of study medication. It is a common duration for observation of antidepressant effects in clinical trials within a hospital-setting and allowed for suitable documentation of clinical trajectories of participants in reference to investigations during the first eleven study days. Furthermore, a third longitudinal and slightly smaller set of investigations was repeated at the end of trial (i.e. excluding imaging and dex/CRH-testing). This was supposed to allow for observation of contrasts in certain parameters that were not necessarily expected to change significantly during the first week of medication. Moreover, possible changes in parameters on a longer-term basis was supposed to create a contrast in reference to the first week of medication.

The trial was controlled in two ways. On one hand, one of the four balanced, parallel patient study groups was receiving placebo during the first seven days of randomized, double-blind medication and therefore served as a positive control arm. On the other hand, age- and gender-matched healthy volunteers were later included in the trial as a supplement to serve as an additional single time-point negative control group. All investigational modalities performed in patients at baseline before medication were also performed in healthy volunteers, except dexamethasone/CRH-testing.

An overview of assessments over the course of the trial is depicted in SPIRIT-conformant Figure 1. The extensive set of applied longitudinal-investigations for deep phenotyping can be categorized into five major technology-focused axes of investigation: 1.) various event-related and resting-state MR neuroimaging including brain connectome measurements, 2.) analysis of stress regulation along the HPA-axis including copeptin and morning cortisol, 3.) gut microbiome composition, 4.) inflammatory proteome markers with potential relevance for neuroinflammation and 5.) two sets of lipidomic analyses. Furthermore, participants were co-enrolled in concomitant trials were reasonably possible and suitable to the design of this trial which are summarized in a separate axis.

**Figure 1:**
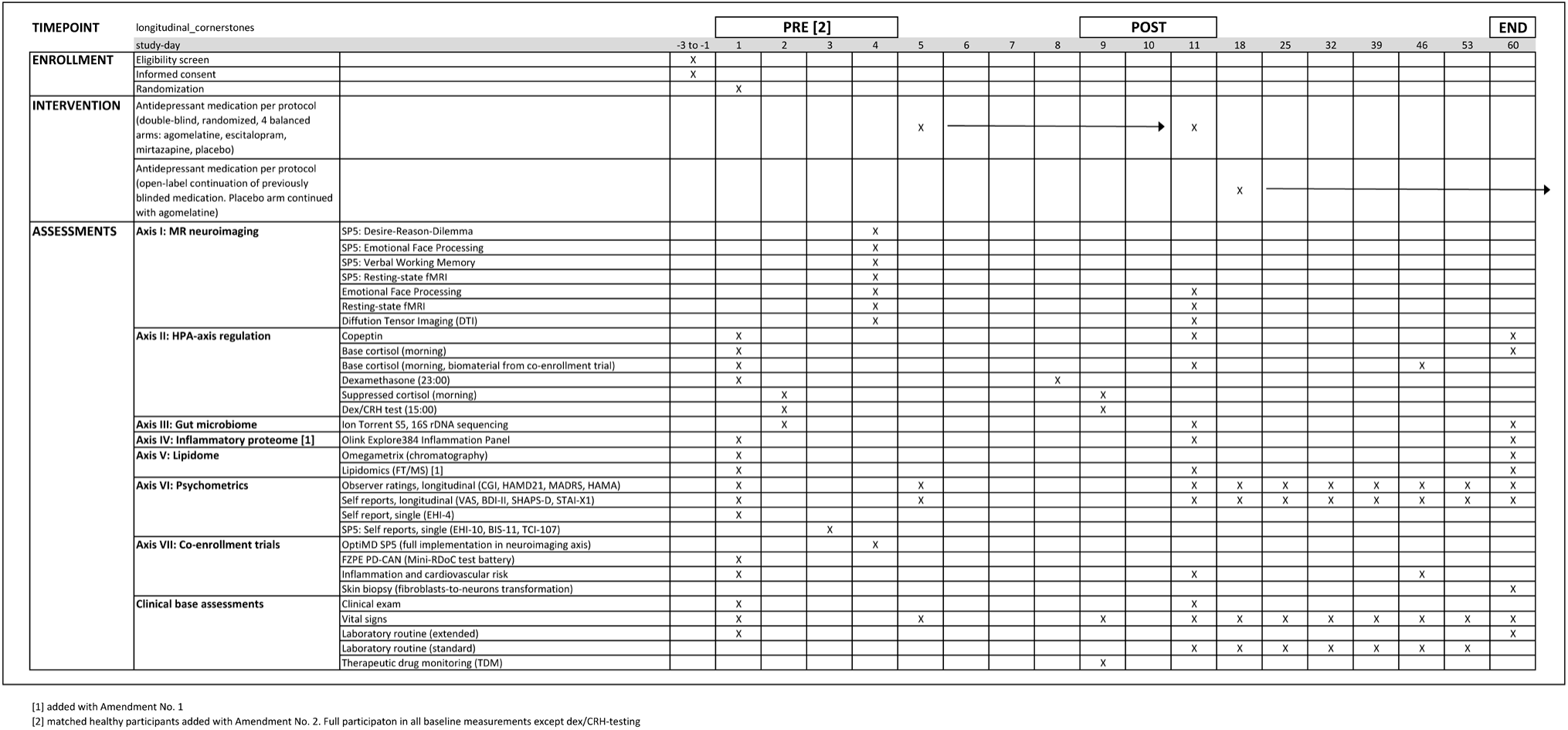
Schedule of interventions and assessments (SPIRIT 2025-conformant timeline)

Pre/post-testing batteries have been the same with exception of imaging for OpiMD subproject 5, which had been designed for single acquisitions at baseline, and except Omegametrix-based fatty acid analyses as recommended by the head of the Omegametrix laboratory.

#### Medication

Participants were randomized into four treatment arms to receive escitalopram 10 mg/d (Cipralex, Lundbeck, Hamburg, Germany), mirtazapine 30 mg/d (AbZ Pharma, Ulm, Germany), agomelatine 25 mg/d (Valdoxan, Servier, Munich, Germany) or placebo for seven days followed by agomelatine 25 mg/d. Blinding was lifted after seven days. Open-label medication usually originated from a different manufacturer than for blinded medication. For non-placebo groups, doses of medications were allowed to be increased after two weeks of treatment (for escitalopram to 20 mg/d, for mirtazapine to 45 mg/d and for agomelatine to 50 mg/d), i.e. from study day 19 onwards. For the placebo group, a corresponding increase in dosage of agomelatine from 25 to 50 mg/d was allowed per protocol from study day 26 onwards.

Additional psychotropic medication was based on clinical necessity and restricted per protocol to zopiclone (up to 15 mg/d) and zolpidem (up to 20 mg/d) given in case patients required sleep medication and lorazepam (up to 3 mg/d) in case inner tensions and agitation required medication in addition to psychotherapeutic support measures. When clinically reasonable, benzodiazepines were avoided on days of dexamethasone/CRH-testing and fMR-imaging and, if possible, one or two days before them.

#### Unblinding procedure

Two different procedures were used for unblinding of randomized study medication. Originally, unblinding had been prepared by therapeutic drug monitoring (TDM) based on chromatographic techniques with a small blood sample taken on study four or five. This approach did not show to be sufficiently reliable and therefore needed to be changed during the course of the trial, as further described in “Protocol deviations during and after first week of double-blind medication”.

Hence, a specific procedure was established for the study team to receive unblinding information while preventing knowledge of placebo intake. According to study protocol, agomelatine 25 mg were openly prescribed on study day twelve in two of the randomized groups: evidently, in the agomelatine group, but also as first prescription after seven days of placebo. Therefore, on the day of unblinding, a study doctor and a study nurse brought the sealed emergency envelope with unblinding information for the respective drug number to the administration office of the department which was not involved in performing the clinical trial. Two secretaries there, were trained in the following procedure: receiving the sealed envelope, opening it and only telling the study doctor and study nurse which medication needs to be continued open-label from then on without showing the envelope to any of them. Afterwards, the opened enveloped was locked away in a specified cabinet by the secretary until the end of the trial.

The complete list for unblinding study medication as provided by the pharmacy of LMU University Hospital Munich was officially opened on 8 October 2025 for quality control and verification of individual unblinding.

### Sample size estimation

In this exploratory, longitudinal deep phenotyping investigation, sample size estimations primarily focused on the acquisition of fMRI data in patients, in particular on amygdala responsiveness (in terms of BOLD signal change) to presentation of emotional faces before and after the first seven days of antidepressant medication. The inclusion of 25 matched healthy volunteers as an additional control group occurred later during the course of the trial through an amendment. Hence, these were not part of the original sample size estimations.

A total patient sample size of 60, with 15 participants per group, was estimated from published literature. In particular, studies originating in the research groups led by Mark A. Mintun and Catherine J. Harmer provided orientation regarding effect sizes to be expected. It may be noteworthy, though, that it used to be uncommon to report standardized effect size estimators in neuroimaging studies [134]. Still, the studies mentioned below were conceptually very similar to our plans and were therefore providing helpful orientation towards effect size estimation. In 2001, Sheline et al. [87] investigated eleven depressed patients and eleven control subjects with longitudinal presentation of emotional faces in tfMRI eight weeks apart with antidepressant therapy in between. In post-hoc comparisons, they could show increased left amygdala activation highest to fearful faces in depressed patients versus controls at baseline which was significantly reduced among patients after antidepressant therapy (F(1,10) = 5.8, p < 0.05)). Similarly, in 2012, Godlewska et al. [90] investigated 39 depressed patients randomized into two groups to receive double-blinded escitalopram 10 mg (N = 19) or placebo (N = 20) and compared these to 17 healthy controls. All participants completed tfMRI with fearful and happy faces once at the end of a seven-day treatment period for patients. The post-hoc analyses of patient data revealed increased right amygdala activation to fearful faces in the placebo group in comparison to patients treated with escitalopram (t[40] = 2.729, p = 0.009).

These reported post-hoc comparisons indicated effect sizes of 1.52 [95% CI: 0.09, 2.90] for within- and 0.86 [95% CI: 0.21, 1.51] for between-comparisons for Cohen’s d, corresponding to 0.37 [95% CI: 0.02, 1.00] and 0.16 [95% CI: 0.02, 1.00] for (partial) η^2^ – implying potentially large effect sizes in terms of Jacob Cohen [135].

In our study design, the primary objective was to observe differential effects of escitalopram, mirtazapine, agomelatine and placebo in a longitudinal pre-/post functional MR neuroimaging design in which task-based fMRI with emotional faces was acquired before and after seven days of double-blind randomized medication.

Therefore, our original sample size estimations were primarily based on an ANOVA with repeated-measures for the comparison of four groups over two time-points. We remained conservative and assumed large effect sizes at the lower end of ‘large’ as described by Jacob Cohen [135], making for an η^2^ of 0.14. Using this assumption for a between-groups comparison with a desired statistical power of 0.8 and a standard significance level of 0.05, yielded a sample size per group of 14 while a within-comparison yielded 15 participants per group. All calculations were performed in SPSS (SPSS Inc., Chicago, Illinois, USA) and in R (R Foundation for Statistical Computing, Vienna, Austria). Considering possible drop-out rates together with challenges in workflows and technology, a recruitment of 80 patients (20 per group) was planned.

Previous experience with longitudinal dex/CRH tests one week apart indicated that the sample sizes estimated here should be well sufficient for this modality, as well. In regard to the gut microbiome modality at the time of trial conception, it was next to impossible to estimate sample sizes for gut microbiome in common microbiology research while first biostatistical foundations for sample size calculations had been laid on the grounds of the Human Microbiome Project applying Dirichlet-multinomial modelling [136].

Simulations for estimating microbiome metrics were deemed unpromising at the time. Reasonable simulations in the microbiome field continue to be challenging today and were so even more, then. Among the many reasons for this, the three most prominent ones are: sequence counts at OTU-level (operational taxonomic unit) are sparse and highly overdispersed and they are phylogenetically related to each other [137]. Hence, good simulations are typically performed with some real data at hand. At the time of trial conception, there were neither pilot data nor publicly available data from related studies available. Moreover, there was a strong heterogeneity of sequencing, preprocessing and analysis methods with a high speed of evolution and change in these fields.

Therefore, expert level experience was the primary orientation and it confirmed that the above estimates should also provide for sufficient power of gut microbiome analyses.

At the time of writing this protocol, about thirteen years after initial sample size estimations, we attempted a retrospective power analysis for gut microbiome data given the estimated sample sizes above. For this, we sought to derive effect sizes from psychopharmacomicrobiomics studies using antidepressants in human participants and found seven relevant studies published since 2019 [138–144]. This approach failed because of at least four major obstacles. First, method and results descriptions were mostly too sparse, incomplete or inconsistent as to serve for comprehension which specific analyses were performed and how. Second, some typical microbiome metrics like alpha and beta diversities were sometimes not reported at all or only in parts. Furthermore, central information of the described study protocol, for example associations of microbiome metrics with given medication were not reported. Third, the two studies that had made their sequencing data publicly available were providing incomplete corresponding metadata making it impossible to recalculate any relevant measures ourselves. Fourth, corresponding authors of three studies that appeared most promising for our goal did not reply to our inquiries.

Beyond lacking details for effect size estimation, the sample sizes of these study designs can be viewed as a general compass. Our planned sample sizes are well in range with these numbers even though our design was most likely conceived much earlier. Six of the seven mentioned studies described longitudinal sampling of which three had their focus on escitalopram [138,139,141] and one contained an escitalopram intervention group [144] while the others were either focused on vortioxetine [143] or naturalistic observations with prescribed psychotropics classified into three large functional categories [142]. Escitalopram group sizes were 11 [139], 16 [144], 17 [138] and 30 [141]. The study mentioned last reported that all of the recruited patients had become responders to escitalopram and chose a second measurement flexible in time depending on the onset of response definition. Hence, a group size of 15 with an overall patient group size of 60 and 25 healthy controls appeared to fit the higher quality end of these published studies.

### Axis I: Neuroimaging

#### General overview of MR scanning workflow

Patients participated in two longitudinal MR scanning sessions one week apart on study-days 4 (pre) and 11 (post). The first session was on the day before the beginning of antidepressant treatment and the second one on the seventh day of antidepressant therapy which was the last day of medication blinding (Figure 1). A total of 64 patients participated in MR imaging at day 4 and 56 patients at day 11 (Table 2). Additionally, 25 matched healthy subjects participated in MR imaging for a single time-point including the full trial scanning protocol of the first imaging day for patients.

**Table 2:**
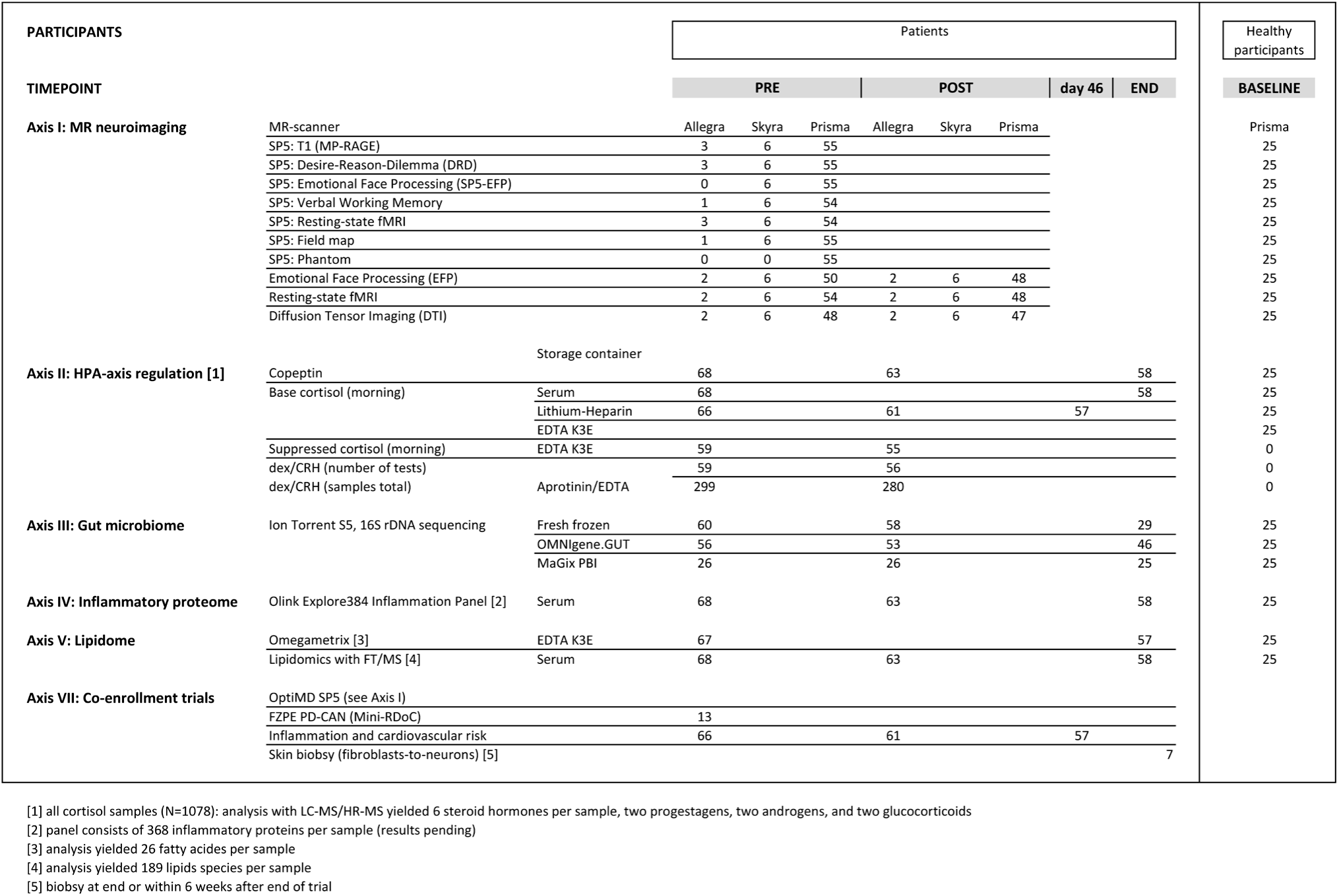
Trial participation: overview of available raw samples. Sample numbers represent presence of samples which have not yet been controlled for quality and validity.

Medical clearance for clinical depression in our hospital included a standardized neuroradiological imaging battery with structural MRI assessments of the brain, at the least for patients who are admitted for the first time, as has been the case for this trial – this kind of medical clearance is independent of any participation in research projects. In order to reduce strain on patients participating in our trial and to ease and simplify their clinical workflow, this routine imaging was performed by our research MR operating team (often including a study physician) at the very beginning of the first scanning session on the MR research tomograph, itself. Thus, most trial participants (i.e. 57 of 64) had their neuroradiological routine assessments combined with research acquisitions immediately afterwards. Clinical acquisitions were directly sent to the Department of Neuroradiology for examination. This set of scans comprised T1- and T2-weighted acquisitions, including MP-RAGE (magnetization prepared rapid gradient-echo) and FLAIR (fluid-attenuated inversion recovery) MR images.

The scan session on the first MR imaging day (i.e. baseline, before beginning of antidepressant treatment) contained three principal investigational components: neuroradiological routine as described above, four experiments from a concomitant trial (OptiMD-SP5), as well as the first of two longitudinal acquisitions for this trial, made up of three experiments. For this reason, MR imaging on study day 4 was split in half with a 45 to 60 min resting period for patients and healthy participants in between. They were free to spend this time along their wishes but were asked to refrain from consuming coffee or tobacco. The majority of patients preferred to return to their ward while some took a walk outside on campus. Before this break, patient images for clinical routine (scan time: 18 min) and for concomitant trial OptiMD subproject 5 (42 min) were acquired. Both components only needed to be run once during the trial. After the resting period, the first longitudinal component (pre) of the Regensburg program (65 min) was run. The exact same post-component was repeated seven days later. The order of acquired MRI sequences was predefined in a scanner workflow protocol and was kept the same throughout the trial. The order is reflected in the described sequence of paradigms in the following sections. Beyond minor exceptions, for example while using the Magnetom Skyra scanner in a back-up capacity, all scanning sessions – for patient and healthy participants – began in the early afternoon around half past one.

#### MR data acquisition

MR Imaging data were acquired with three different 3 Tesla scanners from the Siemens Magnetom device family (Siemens Healthcare, Erlangen, Germany). Changes of scanners were caused by retrofitting of a new research MR scanner. Hence, three initial patients were scanned with a 3 T Siemens Magnetom Allegra and a standard birdcage receiver head coil. The MR tomograph was located on the neuroscience campus (Department of Neuroradiology) of Universität Regensburg which houses all clinical neuroscience departments except neurosurgery. During retrofitting, the next six participating patients were scanned on weekends with a 3 T Siemens Magnetom Skyra and a standard 32-channel receiver head/neck coil, located at the university hospital (Department of Radiology) of Universität Regensburg. Thereafter, 55 patients and 25 healthy volunteers were scanned with a 3 T Siemens Magnetom Prisma and a standard 20-channel receiver head/neck coil located on the neuroscience campus of the Universität Regensburg. This MR tomograph is dedicated for academic research and a joined research funding effort of the faculties for medicine and psychology at Universität Regensburg and of the municipal administration of the neuroscience campus were the scanner is located (medbo, medical facilities of the Upper Palatinate district in Bavaria, Germany).

Head motion during image acquisition was restricted by small cushions placed inside the head coils. Imaging with the Magnetom Prisma scanner was conducted at room temperatures of 20–23 °C, air moistures of 36–44 % as well as at scanner Helium levels of 61–74%. Overall comfort of participants during scanning procedures was set out as a primary requirement. They were placed on whole body foam mats, provided with earplugs for noise protection, blankets to keep warm, cushions for comfortable knee tilt as well as for elbows and wrists against discomfort while lying supine. MR-safe prescription glasses (mediglasses, Cambridge Research Systems, Rochester, UK) were offered in case optical corrections were necessary for interacting with experimental tasks on a screen placed at the end inside of the scanner bore. Lights in the MR chamber were dimmed during the whole acquisition time to provide comfort and higher contrast of stimulus presentations.

Stimuli for experimental tasks were generated either using Presentation software (Neurobehavioral Systems, Albany, New York, USA) for experiments performed for co-enrollment trial OptiMD subproject 5, or with ASF (“A simple framework for behavioral and neuroimaging experiments” [145]) based on the Psychophysics Toolbox [146] for MATLAB [147] for an emotional faces paradigm designed in Regensburg. These stimuli were back-projected using a PROPixx projector with a synchronization controller (VPixx Technologies, Saint-Bruno-de-Montarville, Canada) at a refresh rate of 60 Hz and a resolution of 1024 × 768 pixels onto a translucent rear-projection screen (sandblasted acrylic flat plane custom-made by the Department of Chemistry at Universität Regensburg) placed at the inside end of the MR scanner bore posterior to the head. In contrast to common use settings, the matt side of the screen was oriented towards participants in order to prevent light reflexes from the opposing control room.

Participants viewed stimuli through a detachable standard double mirror (Siemens Healthcare, Erlangen, Germany) provided together with the head/neck 20 coil on which it was mounted and adjusted above their eyes. Distance between eyes and mirror were about 15 cm and the distance between mirror and translucent screen was about 90 cm. This made for a visual angle width of about 22.3° and a visual angle height of about 16.5°, corresponding to 41.5 x 30.5 cm. Screen luminance was measured with a luminance meter (Chroma Meter CS-100, Konica Minolta, Tokyo, Japan) and yielded 0.2 cd/m^2^ for black (RGB 0/0/0), 172 cd/m^2^ for gray (RGB 127/127/127) and 367 cd/m^2^ for white (RGB 255/255/255). Brightness scaling, i.e. non-linear relationship between emitted light and perceived luminance, was addressed by using a gamma lookup table for back-projected stimuli on the screen.

An ergonomic five-button response unit for the right hand received responses from participants which were transferred to a console outside of the scanner (Celeritas Fiber Optic Response System, Psychology Software Tools, Pittsburgh, Pennsylvania, USA) and connected to a PC for stimulus control. Solely button presses for index and middle finger of the right hand were accepted. In this context, index and middle finger were also referred to as ‘left’ and ‘right’ fingers. For training outside of the scanner, button presses with index and middle finger of the right hand were mapped to directly neighboring left and down arrow keys on a standard laptop keyboard providing an appropriate simulation for button presses in the scanner.

All functional MR sequences on all three scanners were based on a multiband sequence (epfid2d1_64) provided by the Center for Magnetic Resonance Research (CMRR, Minneapolis, Minnesota, USA). Functional sequences varied depending on the subproject and the experimental task- or resting-state paradigm. They are therefore outlined with the descriptions of subprojects, below. Scan parameters and acquisition times described below refer to acquisitions with 3 T Siemens Magnetom Prisma.

#### OptiMD SP5: Experimental design and MR scan parameters

##### Overview of imaging workflow for OptiMD SP5 (concomitant trial, multi-center)

Concomitant studies for which participants of this trial had been co-enrolled are summarized as a separate investigational axis (VII) in this study protocol. We make an exception, here, for the multicenter MR neuroimaging investigation for OptiMD, also referred to as subproject 5, i.e. OptiMD-SP5.

Three major reasons for this exception are: first, all participants of this trial have been fully co-enrolled to OptiMD-SP5 and contributed to all paradigms of this trial. Second, while inclusion and exclusion criteria for our trial have been stricter than for SP5 and while SP5 has not been designed to be longitudinal, the major investigational setting and focus of both trials has been similar: to investigate drug-naïve depressed patients at the beginning of the trial and to correlate these results with their prescribed antidepressants and clinical trajectories later on. Third, these strong conceptional overlaps motivated us to fully intregrate SP5 as a constant investigational component in our imaging axis. As a consequence, the Regensburg study center has recruited all of its 64 patients and 25 healthy subjects to participate in the complete imaging program of OptiMD-SP5 and, thus, more than half of the originally conceived cohort.

Therefore, the full study protocol for OptiMD-SP5 is also published as part of this protocol, for the first time, here. It is unique in that, besides the paradigm for Desire-Reason Dilemma, none of the paradigms described here have previously been published with these parameters and task materials. Its leading study center is located within the Division for Experimental Psychopathology and Imaging at the Department of Psychiatry at Universitätsklinikum Heidelberg in Germany.

The protocol consisted of a structural 3-plane localizer, a T1-weighted high-resolution anatomical image, three task-based fMRI paradigms, a resting-state fMRI sequence, a field map and a gel-phantom measurement for quality assessment. Generally, MR sequences were always acquired in the same order as predefined in the manual of the subproject and as correspondingly configured in the scanner workflow protocols. Transaxial slices were aligned parallel to the anterior commissure – posterior commissure line (AC – PC line), based on the measurement volume for the ‘desire-reason dilemma’ task. This alignment was automatically copied to all of the other sequences. Special care was taken on comprising the brain from apical dorsal cortex to both temporal lobes inside the measured volume. Task-based paradigms investigated self-control (‘desire-reason dilemma’ (DRD) task), facial emotion processing, and verbal working memory functions. The SP5 MRI experiment took 42 min scanning time per session for participants and 9 min for the phantom.

Prior to the MRI measurement, participants were trained for every task by a study physician in a calm environment on ward on a laptop running the same software and task protocol as used in the scanner. All stimuli for OptiMD subproject 5 were generated using Presentation software (beginning with Version 18.2, Neurobehavioral Systems, Albany, NY, USA).

For each task, participants were instructed according to a standardized protocol. The standardization included identical instructions through predefined texts and introduction slides, both for the training of the tasks and the actual performance in the MR scanner. Study physicians were trained for this standardized protocol before this trial was co-enrolled.

##### Desire-Reason Dilemma (DRD)

Participants performed a modified version of the so-called ‘desire-reason dilemma’ (DRD) [148] as described by Diekhof et al. [149]. This paradigm had also been deployed in a large multicenter imaging project as part of the German research consortium BipoLife [150] for which Vogelbacher et al. [151] has provided an imaging study protocol.

The paradigm uses trained reward stimuli in two experimental conditions in order to investigate self-control in terms of the ability to successfully control desire for an immediate, but suboptimal reward as required in order to achieve a higher-order long-term goal. The paradigm targets changes in regional brain activation when subjects are not allowed to collect predictors of an immediate reward which forces them to resist impulsive desires and which is has given the paradigm its name.

The experimental design requires two components: appropriate operant conditioning outside of the MR scanner establishes stimulus-response-reward contingencies which are required for the second part, the modified self-control task, inside of the scanner.

During operant conditioning, participants learned to associate eight specific colors and responses with either an immediate reward of ten points (*red*, *green*), a neutral outcome of zero points (*blue*, *yellow*, *pink*, *turquoise*) or an immediate loss of ten points (*brown*, *purple*). Button press with index finger (or ‘left’) was mapped to ‘accepting’ the presented color and a color was ‘rejected’ by button press with the middle finger (or ‘right’). Rejecting reward colors and loss colors, both led to a neutral outcome of zero points while it did not matter which button was pressed with neutral colors. During the conditioning phase, participants were encouraged to initially explore all response possibilities for every color in order to finally maximize their overall outcome. During training, squares of the eight different colors were presented twenty times each in a randomized sequence and only continued to progress after a response button was pressed. This supported conditioning each participant in their own learning curve which was particularly helpful for training patients with depression. Colors associated with loss of points were only presented during conditioning in order to prevent a behavioral preference for the left response button which left the self-control task in the scanner with six colors.

After successful conditioning, training outside of the scanner included rehearsing the actual task which was later run in the scanner. The major goal, here, was to acclimatize participants for the final experimental mechanics including its required short reaction times of less than a second. A minimum total score of 500 points per experimental run was set as a required threshold that intended to reflect successful training of the actual task. Not more than three training runs of the task were allowed to achieve this threshold. For depressed patients, this training was sometimes spread over two to three shorter training sessions along the available resources of the patients and their wishes, as described for safety considerations, above.

The actual task contained equally designed blocks alternating in two different modes. In every block, participants were initially presented with a pair of two target colors randomly selected from the four neutral colors (*blue*, *yellow*, *pink*, *turquoise*). The superordinate goal for each individual task block was to collect the two target colors that were defined at the beginning of each block. These could occur more than once within a block and had to be collected upon each occurrence to reach the goal of the block. Failure to accept a target color (superordinate goal) or to answer within 900 ms led to immediate termination of the current task block, the presentation of “Goal failure” and an outcome of zero points. Every block comprised four or eight trials, with a trial being the presentation of a single color. It provided the possibility to acquire 50 points for successful performance of each individual task block. A single trial had a duration of 1900 ms starting with a blank screen (200 ms) followed by a colored square for 900 ms followed by an immediate feedback for 700 ms.

Pairs of blocks were alternating between two different modes or context rules indicated to the participants by an initial slide showing either “Z” or “B”. The latter introduced two consecutive ‘bonus’-blocks, i.e. desire context, while “Z” introduced two consecutive blocks of the reason context. These two different contexts determined how to treat the non-target colors *green* and *red* for an appropriate performance of the corresponding blocks. In the ‘reason context’ (indicated by initial “Z”) all non-target colors had to be rejected regardless of the originally conditioned reward association in order to achieve the superordinate goal. On the other hand, in the ‘desire context’ (indicated by initial “B”) participants were free to additionally select the two conditioned non-target, i.e. ‘bonus’-colors (*green* and *red*) for an immediate bonus, whereas the remaining two unrewarded non-target colors had to be rejected. Bonuses achieved in the ‘desire context’ were added to the 50 points at the end of a block.

The ‘safe’ way to perform in both context rules was to always remain in the ‘reason context’ and consistently reject the two reward colors. Participants were free to decide to do so. Still, they were consistently and repeatedly motivated to strive for reward maximation reflecting in a higher monetary payout after the experiment. This, however, created a ‘dilemma’ for the participants because they had to control their behavioral tendency to respond to the conditioned reward colors during the ‘reason context’ trials.

In the scanner, the experiment was performed twice, making for 40 task blocks in total which were equally distributed over both contexts. Rewarded and unrewarded non-target color targets could occur up to 30 times each and superordinate goal-relevant target colors could appear up to 60 times each in a pseudorandomized sequence with a counterbalanced trial order. Goal failures reduced these numbers because of their early termination a block. Therefore, participants were required to achieve a minimum total score of 500 points per experimental run during training in order to have enough events of the trial types of interest for the subsequent statistical analyses.

All participants received Euro 30 as base reimbursement. Furthermore, total sum of points acquired during both experimental runs were cashed into additional real money according to a profit grading scheme such that participants could earn up to an additional Euro 20 as reward.

The functional sequence for this paradigm consisted of 185 volumes acquired with a T2*-weighted echo-planar imaging (EPI) sequence with the following parameters: 31 axial slices per volume acquired in ascending interleaved order with prescan-normalization and extended shim mode, phase encoding direction anterior-posterior, echo spacing 0.50 ms and pixel bandwidth (BW) 2604 Hz/Px. GRAPPA (Generalized Autocalibrating Partially Parallel Acquisitions) was used as parallel acquisition technique (PAT) with an acceleration factor of 2, voxel size of 3 mm^3^ isotropic with interslice gap of 20 % (making for a spacing between slices of 3.6 mm), repetition time (TR) 1900 ms, echo time (TE) 30 ms, flip angle 70°, field of view (FOV) 192 mm, in-plane acquisition matrix (AM)= 64×64. Initial volumes of functional scans were planned as dummy volumes to account for T1-saturation and will be discarded before image preprocessing. Sequence acquisition time was 05:57 min.

##### Emotional face processing (SP5 variant)

This implicit emotional face processing task was arranged in a blocked design with alternating blocks of three conditions, i.e. sad facial expressions (S), neutral facial expressions (N) and geometric objects (G, rectangle and ellipse). Overall, the experiment comprised 15 blocks repeating this sequence of three conditions (S-N-G) five times. Each block had a duration of 24 seconds and consisted of the presentation of six stimuli of the respective category (e.g. six sad faces) separated by fixation crosses, providing a baseline condition for later analysis. Portrait pictures with sad and neutral facial expressions were selected from the Radboud Faces Database [152] and were presented during the respective blocks in a pseudorandomized order. The original color portrait photographs as provided by the database were transformed into gray-scale without further clippings or other image modifications. Among the available pictures of 39 Caucasian Dutch adults, the face portraits of 18 subjects were selected from the database (9 males and 9 females each) according to similar ratings for attractiveness, intensity, clarity, genuineness; and the valence of the expression. Adobe Photoshop (Adobe, San Jose, CA, USA) was used to generate two scrambled pictures from this set of faces by splitting images into parts, one rectangular and one elliptical that matched the faces stimuli with respect to mean luminance and other visual features. During blocks in which faces were presented, participants had to judge whether the portrait showed a female (right button press) or a male subject (left button press) making this an implicit emotional face processing task in contrast to explicit task types during which participants directly judge the emotions presented by facial expressions. During blocks in which geometric objects were presented, participants had to judge whether the object was a rectangle (right button press) or an ellipse (left button press). Participants had to respond to presented gender or shape of object.

This implicit emotional face processing task was evaluated in prior fMRI case studies which confirmed reliable activation of key brain regions known to be involved in processing emotional facial expressions, such as the amygdala and the fusiform gyrus. Furthermore, at least two other trials [153,154] had already shown the feasible use and validity of the Radboud Faces Database in contexts of depression before this trial was initiated.

Differences between this emotional face processing paradigm and the one configured for the Regensburg study center are shown in Table 3.

**Table 3:** Conceptual differences between emotional face processing paradigms employed in this trial.

|  | **Regensburg** | **OptiMD SP5** |
| --- | --- | --- |
| experimental runs | 3 | 1 |
| acquisition time per experiment [min] | 10:00 | 7:24 |
| total experiment duration [min] | 31 | 8 |
| total volumes per run | 297 | 219 |
| faces stimuli database | Karolinska Directed Emotional Faces (KDEF) | Radboud Faces Database (RaFD) |
| face presentation | clipped black/white face only<br>(behind elliptical passepartout) | full portrait<br>(original from database) |
| stimuli conditions | 6<br>(sad – happy – fearful – angry – neutral – scrambled) | 3<br>(sad – neutral – scrambled) |
| available faces | 32 female, 32 male | 9 female, 9 male |
| number of stimuli | 384 | 48 |
| experimental blocks | 18 (= 6 * 3) | 15 (= 3 * 5) |
| block duration [s] | 16 | 24 |
| baseline presentation | red dot on black<br>(persisting on stimuli) | white cross on black |
| block sequence | randomized | fixed: sad – neutral – scrambled |
| stimuli per block | 10 | 6 |
| scrambled objects | 64<br>(bottom darker – top darker) | 2<br>(ellipse – rectangle) |
| scrambling method | FFT-phase scrambling (details: ...) | manual image splitting and<br>transformations |
| button press logic | index finger = female / scrambled bottom darker | index finger = male / ellipse |

The functional sequence for this paradigm consisted of 219 volumes acquired with a T2*-weighted echo-planar imaging (EPI) sequence with the following parameters: 33 axial slices per volume acquired in ascending interleaved order with prescan-normalization and extended shim mode, phase encoding direction anterior-posterior, echo spacing 0.50 ms and pixel bandwidth (BW) 2604 Hz/Px. GRAPPA was used as parallel acquisition technique (PAT) with an acceleration factor of 2, voxel size of 3 mm^3^ isotropic with interslice gap of 20 % (making for a spacing between slices of 3.6 mm), repetition time (TR) 2000 ms, echo time (TE) 30 ms, flip angle 70°, field of view (FOV) 192 mm, in-plane acquisition matrix (AM)= 64×64. Initial volumes of functional scans were planned as dummy volumes to account for T1-saturation and will be discarded before image preprocessing. Sequence acquisition time was 07:24 min.

##### Verbal working memory

Implementation of this paradigm was considered optional, but consistently run at our study center. Participants performed a modified Sternberg item recognition task with letters [155] (verbal working memory task, M) in alternation with a letter case judgement task (control task, C) similar as described before [156]. In both tasks, four different target letters were randomly chosen from a set of fourteen preselected letters and visually presented in white on a black background for 1.5 s, followed by a delay of 4 s during which a fixation cross was displayed. Then a probe letter was presented for 1 s, followed by a 1 s-fixation cross. Participants had to respond within 2 s whether or not this probe letter matched one of the target letters (left button = match, right button = no match).

Visual presentation of stimuli was matched between both tasks, making the tasks undistinguishable for participants except how they were trained to behave during each task as indicated by the introductory cue. For the memory task, participants were trained for rapidly repeated subarticulatory (i.e. non-motor) rehearsal to remember the letters presented. Letters were taken from a set of fourteen letters consisting of {b, c, d, e, f, g, l, m, n, p, r, s, t, w}. For the control task variant, participants were instructed to focus on the appearance of the probe letter to come in order to be able to press the correct response button as quickly as possible. Target letters were presented in upper case, the probe letters in lower case or vice versa in a pseudorandomized order to prevent participants from using visual memory strategies.

The overall experiment comprised twelve alternating blocks of control and memory task (six each) which began randomly with one or the other. Each block was 24 s long and consisted of three trials of the same task type 3 x 7.5 s, and a 1.5 s-cue at the beginning of each block indicated whether memory tasks or judgment tasks had to be performed in the upcoming block.

The functional sequence for this paradigm consisted of 164 volumes acquired with a T2*-weighted echo-planar imaging (EPI) sequence with the following parameters: 33 axial slices per volume acquired in ascending interleaved order with prescan-normalization and extended shim mode, phase encoding direction anterior-posterior, echo spacing 0.50 ms and pixel bandwidth (BW) 2604 Hz/Px. GRAPPA was used as parallel acquisition technique (PAT) with an acceleration factor of 2, voxel size of 3 mm^3^ isotropic with interslice gap of 20 % (making for a spacing between slices of 3.6 mm), repetition time (TR) 2000 ms, echo time (TE) 30 ms, flip angle 70°, field of view (FOV) 192 mm, in-plane acquisition matrix (AM)= 64×64. Initial volumes of functional scans were planned as dummy volumes to account for T1-saturation and will be discarded before image preprocessing. Sequence acquisition time was 05:34 min.

##### Resting-state fMRI (5 min, eyes open in fixation)

The experiment for OptiMD subproject 5 ended with a resting-state fMRI acquisition with a duration of 5:18 min. Participants were asked to relax, keep their eyes open and fixate on a fixation cross.

The functional sequence consisted of 156 volumes acquired with a T2*-weighted echo-planar imaging (EPI) sequence with the following parameters: 33 axial slices per volume acquired in ascending interleaved order with prescan-normalization and extended shim mode, phase encoding direction anterior-posterior, echo spacing 0.50 ms and pixel bandwidth (BW) 2604 Hz/Px. GRAPPA was used as parallel acquisition technique (PAT) with an acceleration factor of 2, voxel size of 3 mm^3^ isotropic with interslice gap of 20 % (making for a spacing between slices of 3.6 mm), repetition time (TR) 2000 ms, echo time (TE) 30 ms, flip angle 70°, field of view (FOV) 192 mm, in-plane acquisition matrix (AM)= 64×64. Initial volumes of functional scans were planned as dummy volumes to account for T1-saturation and will be discarded before image preprocessing. Sequence acquisition time was 05:18 min.

##### Anatomical images

Both imaging subprojects required anatomical images for later coregistration. These were acquired at the end of the first half of the first scan day.

High-resolution T1-weighted 3D-images were acquired using a magnetization prepared rapid gradient-echo sequence (MP-RAGE) with the following parameters: 160 slices acquired in ascending order with prescan-normalization, phase encoding direction anterior-posterior, echo spacing 3.54 ms and pixel bandwidth (BW) 751 Hz/Px. GRAPPA was used as parallel acquisition technique (PAT) with an acceleration factor of 2, voxel size of 3 mm^3^ GRAPPA as parallel acquisition technique (PAT) and an acceleration factor of 2, voxel size of 1 mm^3^ isotropic with an interslice gap of 50 %, repetition time (TR) 1910 ms, echo time (TE) 3.67 ms, flip angle 9°, field of view (FOV) 250 mm, in-plane acquisition matrix (AM)= 256×256.

High-resolution T2-weighted 3D-images were acquired using a fluid-attenuated inversion recovery sequence (FLAIR) with the following parameters: 192 slices acquired in interleaved order with prescan-normalization, phase encoding direction anterior-posterior, echo spacing 3.54 ms and pixel bandwidth (BW) 751 Hz/Px. GRAPPA as parallel acquisition technique (PAT) and an acceleration factor of 2, voxel size of 0.5 x 0.5 x 0.9 mm^3^, repetition time (TR) 5000 ms, echo time (TE) 386 ms, field of view (FOV) 240 mm, partial Fourier 7/8 and in-plane acquisition matrix (AM)= 256×256.

##### Quality assurance for OptiMD SP5 multicenter imaging

###### Field map

A field map was acquired with a double-echo spoiled gradient echo sequence with the following parameters: 31 axial slices per volume acquired in ascending interleaved order, phase encoding direction anterior-posterior and pixel bandwidth (BW) 260 Hz/Px, voxel size of 3 mm^3^ isotropic with interslice gap of 20 %, repetition time (TR) 418 ms, echo time (TE) 8.27 ms, flip angle 40°, field of view (FOV) 192 mm, in-plane acquisition matrix (AM)= 64×64. The field map was based on a multiband sequence (fm2d2r) provided by the Center for Magnetic Resonance Research (CMRR, Minneapolis, Minnesota, USA). Sequence acquisition time was 00:56 min and generated a magnitude image and two phase images. The field map image will then be computed from the two-phase images.

###### Phantom acquisition

MR scanner characteristics were assessed by regular measurement of a MRI phantom. The phantom was a 23.5 cm long and 11.1 cm-diameter cylindrical plastic vessel (Rotilabo, Carl Roth, Karlsruhe, Germany) filled with a mixture of 62.5 g agar and 2000 ml distilled water. Phantoms were built by the Experimental Psychopathology and Imaging Unit at Universitätsklinikum Heidelberg and sent to each participating study center. All study sites, therefore, used the same type of phantom. Participants were given a rest period after MR scanning for OptiMD subproject 5 during which phantom data were acquired. The alignment of the long phantom axis was parallel to the main scanner axis and in the center of the head coil. The measurement volume was manually centered on the phantom with a slice direction perpendicular to the phantom body (see Supplementary Figure S1 in Vogelbacher et al. [157]).

Temporal stability is particular important for fMRI measurements during which MR scanners are highly stressed. Therefore, the MRI phantom was measured with 250 volumes of a similar EPI sequence as used for all other functional sequences in this project. Hence, the functional sequence for this paradigm consisted of 250 volumes acquired with a T2*-weighted echo-planar imaging (EPI) sequence with the following parameters: 34 axial slices per volume acquired in ascending interleaved order with extended shim mode, phase encoding direction anterior-posterior, echo spacing 0.49 ms and pixel bandwidth (BW) 2894 Hz/Px. GRAPPA was used as parallel acquisition technique (PAT) with an acceleration factor of 2, voxel size of 3 mm^3^ isotropic with interslice gap of 20 % (making for a spacing between slices of 3.6 mm), repetition time (TR) 2070 ms, echo time (TE) 30 ms, flip angle 70°, field of view (FOV) 192 mm, in-plane acquisition matrix (AM)= 64×64. Sequence acquisition time was 08:42 min.

#### Experimental design and MR scan parameters for this trial

##### Overview of imaging workflow for this trial (single-center)

Pharmaco-fMRI, i.e. longitudinal MR imaging one week apart around the first week of antidepressant pharmacotherapy in drug-naïve patients with clinical depression in the early afternoon is a core design element of this trial. Hence, this imaging workflow was first run after the resting period on the first day of MR imaging (study day 4) at baseline before beginning of double-blinded, randomized antidepressant treatment. The exact same scan protocol was run for a second time on day seven of antidepressant treatment before lifting of blinding at about the same time of day. The scan protocol comprised three paradigms that were run in the following sequence: first, a long resting-state fMRI with closed eyes, second, task-fMRI with an emotional face processing task that was repeated three times and, third, diffusion tensor imaging.

##### Resting-state fMRI (22 min, eyes closed)

At first, a resting-state fMRI was acquired during which participants were asked to relax, lie still in the MRI scanner for 22 minutes, close their eyes and not think of any specific themes while staying awake. This resting-state fMRI was not only intended for the investigation of functional connectivity but also for dynamic functional connectivity analyses and for the combination with structural connectivity information yielded with diffusion tensor imaging.

The functional sequence consisted of 660 volumes acquired with a CMRR T2*-weighted echo-planar imaging (EPI) sequence with the following parameters: 33 axial slices per volume acquired in ascending interleaved order with prescan-normalization and extended shim mode, phase encoding direction anterior-posterior, echo spacing 0.50 ms and pixel bandwidth (BW) 2604 Hz/Px. GRAPPA was used as parallel acquisition technique (PAT) with an acceleration factor of 2, voxel size of 3 mm^3^ isotropic with interslice gap of 20 % (making for a spacing between slices of 3.6 mm), repetition time (TR) 2000 ms, echo time (TE) 30 ms, flip angle 70°, field of view (FOV) 192 mm, in-plane acquisition matrix (AM)= 64×64. Initial volumes of functional scans were planned as dummy volumes to account for T1-saturation and will be discarded before image preprocessing. Sequence acquisition time was 22:06 min.

##### Emotional face processing (Regensburg variant)

###### Stimulus selection from KDEF database for emotional faces

All facial pictures for this paradigm were selected from the Karolinska Directed Emotional Faces (KDEF) database [158]. This database provides 32-bit color images (i.e. 16.7 million colors) at a size of 562 x 762 pixels per image. They originate from two photographing sessions of 70 adult humans (35 female) between 20 and 30 years of age expressing the seven basic emotions [159], each shown in five different photographic angles making for 4900 available images. KDEF images are provided such that the central horizontal image axis is passing through the eye centers.

At first and before individual assessment of images, only straight profile (‘S’) pictures from the first photo session (‘A’) were chosen for this trial and facial expressions for ‘disgusted’ and ‘surprised’ were omitted. This left 350 stimulus images for five facial expressions which are abbreviated in the KDEF database with the letters noted in brackets: afraid (‘AF’), angry (‘AN’), happy (‘HA’), neutral (‘NE’) and sad (‘SA’). From this standard set, 31 images were excluded (three per condition with exception of sad men), mostly corresponding to three specific participants per gender. Except for the ‘sad’ condition, KDEF participants F19, F20 and F26, as well as M19, M20 and M26 were excluded. In the ‘sad’-condition, the following KDEF participants were excluded: F15, F16 und F31, as well as M10, M12, M30 and M31. In compensation for the exclusion of four men for ‘sad’ condition, one (M3) has been doubled. Furthermore, for M15, ‘afraid’ and ‘angry’ condition were taken from photo session ‘B’ and the same for M12 ‘angry’ and M28 ‘happy’.

This individual selection from the KDEF database made for a set of 319 straight facial pictures of 32 women and 32 men each showing five different emotions, and of which one was doubled to complete the set of stimuli to 320. All of the 64 selected images for condition ‘sad’ were copied for scrambling and added as a sixth additional condition for analytical control, making for a full set 384 stimuli.

###### Image processing of selected KDEF portraits

The original KDEF color images with a size of 562 x 762 pixels and a digital RGB-scan color-depth of 32 bit were further used without changing image and canvas sizes. However, the following graphical modifications were made: first, images were transformed to black-and-white and second, clipped behind elliptical black passepartouts.

Each of the selected KDEF photographs was transformed into black-and-white using rgb2gray within Matlab (version R2015b). This function converts RGB values to grayscale values by calculating a weighted sum of the individual R-, G-, and B-components with the following formula: (0.298936021293775 * R) + (0.587043074451121 * G) + (0.114020904255103 * B).

Black-and-white transformed images were then clipped behind elliptical black passepartouts to reduce possible confounding visual stimuli. All passepartouts were of the same size, with a minor elliptical axis of 300 px and a major axis of 400 px. Ellipses were placed such that their lower vertex coincided with the chin line. This provided for a primary focus on the face, mostly excluding ears and hair and the majority of the neck. As a side effect of this narrow image focus, conscious gender identification became challenging with many participants.

###### Face scrambling

Each of the 64 selected images from 32 females and 32 males, all of them for the ‘sad’ condition, were copied to create a sixth condition with scrambled images, i.e. images in which all identifiable features were obscured. While preserving certain image properties, this also ensures that feature identification by unconscious neurobiological processes is rendered impossible.

For this, Fast Fourier Transform (FFT)-phase scrambling was used. The spatial domain (i.e. pixels) of the image was transformed into a frequency domain by computing a two-dimensional fast Fourier transform (fft2 in matlab). This yields a complex number representation of the image as magnitude and phase, in which the latter represents structural information of the image like edges or shapes. Therefore, it is quite common to randomize phase values for face scrambling in this context, after which an inverse FFT (ifft2, matlab) can be used to re-transform the resulting magnitude/phase maps to produce a scrambled version of the original image.

One half of these images was scrambled such that the bottom half of the image appeared darker after scrambling while in the others, the top half appeared darker. These scrambled images were equally presented behind the same elliptical black passepartouts left in the same position as for the original clipped photograph.

###### Randomization of stimuli and gender

The trial design required each participant to complete three EFP experiments of 10 min each in a row on study day 4 and the same on study day 11 making for a total of six experimental runs. Stimuli and gender to be presented were randomized for each participant and for each experiment individually and independently. Randomized stimuli were prepared immediately before beginning an experiment as part of an ASF experiment workflow with a pseudorandom number generator using Mersenne Twister as generating algorithm and a fixed seed individually derived for each participant from current date and time. Stimuli were arranged in a 64 x 18 matrix (64 individuals in rows and 6 conditions, each repeated 3 times during one experiment in columns). First, conditions were randomized across columns. Any condition was allowed to be presented first, including the ‘scrambled’ condition. Repetition of individual stimuli were prohibited within a block but possible between blocks. Gender was shuffled within each column while preserving gender balance in each block with a binomial probability density function. There was no balancing between randomizations for each of the three experimental runs.

###### Experimental design for this EFP paradigm

This implicit emotional face processing task was arranged in a blocked design with alternating blocks of six conditions, i.e. afraid facial expressions (‘AF’ as used by KDEF database), angry facial expressions (‘AN’), sad facial expressions (‘SA’), happy facial expressions (‘HA’), neutral facial expressions (‘NE’) and scrambled images (‘SCR’) of which each was repeated three times making for 18 blocks. Each block had a duration of 16 s and consisted of ten randomized stimuli with balanced gender from one condition. For example, ten sad faces from women and men were presented for 1600 ms each – the duration of each stimulus (1.6 s) corresponded to 96 refreshed images on the monitor at a refresh rate of 60 Hz. They were interlaced with the presentation of a centered red fixation dot (RGB 255/0/0, diameter 10 pixels) on black background (RGB 0/0/0) for 16 s including beginning and end of experiment. The same red dot was overlaid on face stimuli in the same image center position for improved eye control. Face pictures were positioned such that the image center, and hence the red dot, was located between the eye brows along a central horizontal line passing through eye centers. This made for 19 red fixation dot blocks and an experimental duration of 592 s (i.e. 19 x 16 sec + 18 x 16 sec). This blocked experiment of 10 min duration was run three times in a row in each scanning session.

During blocks in which faces were presented, participants had to perform a gender judgement task making this an implicit emotional face processing task in contrast to explicit task types during which participants directly judge the emotions presented by facial expressions. Female faces required a left button press (index finger of right hand) and male faces a right button press (middle finger of right hand). During blocks in which scrambled pictures were presented, participants had to judge whether there were more dark spaces at the bottom of the image (left button press) or at the top (right button press). Participants had to respond to presented gender or darkness of spaces within slightly less than the time of face presentation (i.e. 1595 ms) to enable a technical tolerance and allow for code to return in time for next stimulus.

Differences between this emotional face processing paradigm and the one used for OptiMD subproject 5 are shown in Table 3.

The functional sequence for this paradigm consisted of 297 volumes acquired with a CMRR T2*-weighted echo-planar imaging (EPI) sequence with the following parameters: 33 axial slices per volume acquired in ascending interleaved order with prescan-normalization and extended shim mode, phase encoding direction anterior-posterior, echo spacing 0.50 ms and pixel bandwidth (BW) 2604 Hz/Px. GRAPPA was used as parallel acquisition technique (PAT) with an acceleration factor of 2, voxel size of 3 mm^3^ isotropic with interslice gap of 20 % (making for a spacing between slices of 3.6 mm), repetition time (TR) 2000 ms, echo time (TE) 30 ms, flip angle 70°, field of view (FOV) 192 mm, in-plane acquisition matrix (AM)= 64×64. Initial volumes of functional scans were planned as dummy volumes to account for T1-saturation and will be discarded before image preprocessing. Sequence acquisition time was 600 s.

##### Diffusion tensor imaging

The final image acquisition at each of the two scanning days was diffusion tensor MR imaging during which participants were asked to relax, lie still in the MRI scanner for 12 minutes, close their eyes while not falling asleep. Beforehand, they were reminded of the changing and louder acoustic noises that might also cause perceptible vibrations of the scanning bed. This MR tractography imaging was intended to be used for the investigation of structural connectivity and for the combination with functional connectivity data described above.

The water diffusion sequence was acquired with a CMRR single-shot echo-planar imaging (EPI) sequence with the following parameters: one DTI acquisition (NEX = 1) with 65 gap-free axial slices covering the whole brain, 64 non-collinear directions (b = 1000 s/mm^2^) and one non-diffusion weighted volume (b = 0 s/mm^2^) in interleaved order with prescan-normalization and standard shim mode, phase encoding direction anterior-posterior, echo spacing 0.66 ms and pixel bandwidth (BW) 1776 Hz/Px. GRAPPA was used as parallel acquisition technique (PAT) with an acceleration factor of 2, voxel size of 1×1×2 mm, repetition time (TR) 10800 ms, echo time (TE) 82 ms, flip angle 90°, field of view (FOV) 256 mm, Partial Fourier 6/8 and in-plane acquisition matrix (AM)= 128×128. Sequence acquisition time was 12:16 min.

### Axis II: HPA-axis regulation, copeptin and morning cortisol

#### Morning cortisol

Morning fasting cortisol in serum was analyzed from two different blood fractions (serum and heparinized plasma) to increase granularity in time such that morning cortisol information became available for four time-points at study days 1, 11, 46 and 60 instead of two. This was made possible through slight shifts and exchanges in sample logistics during the analysis phase after end of recruitment. For one, morning cortisol was taken from blood drawn from patients and healthy participants at the beginning and end of the study (study days 1 and 60) into Serum CAT S-Monovettes: (Sarstedt, Nümbrecht, Germany) that was immediately centrifuged at 1516 g for 10 min at precooled 8 °C before long-term storage at –80 °C in micro tube aliquots. Furthermore, blood from patients and healthy participants was drawn into Lithium-Heparin LH S-Monovettes (Sarstedt, Nümbrecht, Germany) at study days 1, 11 and 46, continuously kept cool on ice until centrifuging at 2877 g for 10 min at precooled 4 °C before supernatant heparinized plasma was stored at –80 °C in micro tube aliquots until analysis. Additionally, blood from healthy participants was additionally treated like suppressed morning cortisol samples and drawn into EDTA K3E S Monovettes (Sarstedt, Nümbrecht, Germany) and centrifuged at 2877 g for 10 min at room temperature before supernatant plasma was stored at –80 °C in micro tube aliquots until analysis.

This made for 126 serum cortisol samples from 68 patients and 184 heparinized plasma cortisol samples from 66 patients. All of the 25 healthy participants provided blood for all three factions (25 + 25 + 25 samples) at the same time-point for quality control and possible normalization of data.

#### Dex/CRH-testing

The so-called ‘combined dexamethasone-suppression / corticotropin-releasing hormone stimulation (dex/CRH) test’ is a dexamethasone suppression test (DST, [160]) extended by an afternoon CRH challenge [23] with a 75-minute follow-up. It is regarded to be a sensitive method for investigating the regulatory capacity of the hypothalamic-pituitary-adrenal (HPA) axis in depressed patients as expressed in cortisol and ACTH (adrenocorticotropic hormone, i.e. corticotropin) responses during the course of the test.

Dexamethasone is a first generation synthetic glucocorticoid, first synthesized in 1958 [161]. Its primary chemical characteristics in relation to cortisone are a methyl-group at C-16 in the tetracyclic steroid core which practically eliminates mineralocorticoid effects and a fluorine atom at C-9α which enhances glucocorticoid activity, making it one of the most potent anti-inflammatory steroids available. Its plasma half-life after oral intake is around 5.5 hours [162] while its so-called biological half-life is described to be in a range of 36 to 72 hours [163,164], thus categorizing it into long-acting glucocorticoids based on its effects on HPA axis suppression and immunologic reactions.

The primary objective of this trial in regard to dex/CRH testing was to explore differential effects in longitudinal dex/CRH test results one week apart in relation to treatment with three different antidepressants originating from different pharmaceutic classes. Furthermore, associations of these longitudinal results with results from other research modalities in this trial might help to gain a more granular understanding of pathophysiological processes in clinical depression.

A dex/CRH-testing environment was set up on the research ward described in “General patient safety”. There, two separate, neighboring rooms were used as a laboratory space for dex/CRH-testing. One was shielded and furnished agreeably including a comfortable bed for the patient to be tested. The neighboring room was a research office equipped with some laboratory equipment. From there, the test was monitored and controlled including immediate blood processing. Both rooms were connected in two ways, only: first, by a soundproof lock in the wall at arm level of the patient bed serving for connected tubing of blood sampling and blood pressure measurements and, second, by discreet, non-recording closed-circuit television including microphones for monitoring patient safety and the course of the experiment. There has been no direct interaction between research personnel and patient during the whole course of the experiment with a single exception: asking the patient for the current condition and possible side effects immediately after CRH challenge.

Two hours after lunch, around 14:00 hours patients were being led to the patient laboratory room and reminded of the setting including cameras and microphones for which they already had provided informed consent as well as of regular blood pressure measurements and possible side effects from CRH bolus injection. A short history was taken of their current and general daily tobacco and coffee consumption until the afternoon. Then, they were asked to make themselves comfortable, rest supine in the bed and stay awake for the whole period of the experiment. Furthermore, they were asked to refrain from potentially stressful influences like particular text messaging on their phone, agitating movies or books and, alternatively, to rather focus on calming media without surprises.

At that point, dexamethasone 1.5 mg (dex) (Jenapharm, Jena, Germany) had been administered orally in the previous evening (at 23:00 hours) for suppression and verified by measuring suppressed fasting cortisol in the morning. Constant intravenous access via catheter in a forearm was established before 14:30 hours and kept patent with a continuous physiological sodium chloride (0.9 %) drip at 20 ml/h. This advance intravenous access served to decouple potential experiences of physical pain or fear and their possible influences on stress reactivity from the test itself. The intravenous catheter was connected to a tubing extension into the neighboring laboratory, from where a CRH bolus would be injected and where blood samples were taken, processed and stored in aliquots.

In the most common and short version of this test applied here, five blood samples are drawn beginning with a baseline sampling in the afternoon at 15:00 hours (followed by 15:30, 15:45, 16:00, and 16:15 hours). At the same time-points, blood pressure is taken. Shortly after the first blood sample (baseline), a bolus of human corticotropin-releasing hormone 100 µg (CRH FERRING, Ferring, Kiel, Germany) prepared in a 2 ml syringe was injected starting at 15:02 hours and flushed with 10 ml sodium chloride over a total period of 20 to 30 sec. Cortisol concentrations at baseline reflect the suppressive effects of dexamethasone intake the evening before. The other four samples, then, show the response to challenge with CRH injection.

Neuroendocrine response behavior after CRH-challenge is most often assessed by measuring the total areas under the curve (AUC), either relative to increase from baseline or relative to ground by adding the areas of the trapezoids spanned up by the data points together with baseline or groundline [165].

While calculating AUC has been the primary plan for this trial, there are more options to investigate dex/CRH-test results. For one, it is possible to take a delta value between the maximum of the curve and its initial baseline. Second, if dex/CRH-testing is performed longitudinally, as it has been for this trial, the difference between the longitudinal curve maxima (i.e. peak cortisol levels) may serve as an indicator for change in cortisol response over time. Third, patients may be stratified into non-suppressors, suppressors and intermediate suppressors [166]. For one, this requires the definition of a threshold for the plasma cortisol concentration measured before CRH challenge similar to a common DST. It is supposed to reflect how far dexamethasone has suppressed cortisol release towards the beginning of the test at 15:00 hours. Two different criteria have been proposed for this threshold: the Carroll criterion at 50 ng/ml derived from a dexamethasone suppression test (DST) [26] which has also been applied by Kunugi et al. [166] and Ising et al. [29] for dex/CRH testing; and the Heuser criterion which was originally reported as 40 ng/ml [25] and bias corrected to 27.5 ng/ml, later [30]. Thus, non-suppressors and suppressors are distinguished in the same way as in a DST: the first show a baseline cortisol above the threshold and the latter are below. Definition of intermediate suppressors as proposed by Kunugi et al. [166] additionally require the cortisol response of a CRH-challenge: their baseline cortisol is below the threshold, but their peak cortisol after CRH-challenge is above.

This dex/CRH test was performed longitudinally twice on study days 2 and 9 (with dexamethasone suppression the evenings before) such that each test was followed by a 2-day recovery period before neuroimaging and gut microbiome sampling. Sixty-one patients went through a first dex/CRH test at baseline of whom 58 also completed a longitudinal second dex/CRH test after one week. Samples from two of the first dex/CRH tests could not be used: one only yielded the first two picks and another has remained below limit of quantification in LC-MS / HR-MS in eight of ten samples. Similarly, samples from two of the second dex/CRH tests cannot be used: the same patient with results below limit of quantification (LOQ) and one with only the first pick available. This made for 299 samples from 59 patients for the first and 280 samples from 55 patients for the second dex/CRH test which can probably be used. Analysis of blood samples is described at the end of this section.

Blood from dex/CRH testing was processed according to a long-standing protocol used by our colleagues at Max-Planck-Institute for Psychiatry in Munich, Germany, in order to maintain comparability with existing large databases. Five milliliters of drawn blood for each sample were immediately dispensed into a precooled test tube (8 °C) prefilled with 150 µl of aprotinin with 11700 Kallikrein-inhibitory units per ml (Calbiochem, cat 616399-500KU until 2017, then Sigma-Aldrich, catalog number A6279; both companies of Merck, Darmstadt, Germany) together with 150 µl of aqueous 4 % Edetate disodium (prepared by our pharmacy) and gently mixed by hand.

These tubes were then centrifuged (Hettich Universal 320, Andreas Hettich, Tuttlingen, Germany) at 1516 g for 10 min at precooled 8 °C. Supernatant was distributed into two aliquots (about 800 and 1500 µl) and stored at –80 °C until analysis for all samples at once.

Additionally, blood for suppressed morning cortisol after dexamethasone intake in the evening was drawn into EDTA K3E S Monovettes (Sarstedt, Nümbrecht, Germany) and centrifuged at 2877 g for 10 min at room temperature before supernatant plasma was stored at –80 °C in micro tube aliquots until analysis. Thus, from 119 performed dex/CRH tests, 59 from the morning of the first and 55 from the morning of the second dex/CRH test were available and analyzed together with dex/CRH test samples as described at the end of this section.

#### Copeptin

Copeptin, or CT-proAVP, a C-terminal glycopeptide of 39 amino acid residues, cleaved from a large precursor peptide together with arginine vasopressin in equimolar ratios, was measured using a B·R·A·H·M·S Kryptor compact PLUS with an automated Thermo Scientific B·R·A·H·M·S Copeptin proAVP KRYPTOR assay (Thermo Fisher Scientific B·R·A·H·M·S, Hennigsdorf, Germany). The latter is an automated immunofluorescent assay [39] for the quantitative analysis of copeptin in human serum or plasma.

Analysis requires a volume of 50 µl per sample. The manufacturer provides a detection limit for the assay of 0.69 pmol/l (with 1 pmol/l = 4.02 pg/ml) and a measuring range (with automatic dilution) of 0.7 to 2000 pmol/l. Functional assay sensitivity as detected by inter-assay precision of 20 %, was calculated as 1.08 pmol/l, located at the upper limit as provided by the manufacturer.

In total, 214 serum samples were analyzed which had been taken in the mornings between 7:00 and 07:30 hours before breakfast and without previous HPA manipulation: one hundred and eighty-nine longitudinal serum samples from 68 patients over three time-points (study days 1, 11 and 60) and 25 samples from 25 healthy subjects. Fasting venous blood from patients and healthy participants had been collected into serum containers (Serum CAT S-Monovettes, Sarstedt, Nümbrecht, Germany) and were immediately centrifuged at 1516 g for 10 min at precooled 8 °C before long-term storage at –80 °C in micro tube aliquots. There was one short freeze-thaw cycle between long-term storage and copeptin analysis.

#### Analysis of cortisol and five further steroid hormones with LC-MS / HR-MS

In total, 1103 cortisol-related samples were analyzed for this axis of investigation by employing liquid chromatography-high resolution tandem mass spectrometry (LC-MS / HR-MS) with a hybrid quadrupole-Orbitrap mass spectrometer and a run-time of 5.3 min offering highly selective quantification as described in detail by Matysik and Liebisch [167] – of these, probably 1078 samples can be used for further analysis. For this project, six of the eight steroid hormones from Matysik and Liebisch were analyzed: two progestagens: 17-hydroxyprogesterone and progesterone, two androgens: androstenedione and testosterone, as well as two glucocorticoids: 11-deoxycortisol and cortisol (without corticosterone and cortisone). Limit of quantification (LOQ) varied by target species and ranged from 0.03 to 1.9 ng/ml serum hormone concentration (with 1.9 ng/ml for cortisol).

First, steroid hormones were extracted from human serum samples by liquid–liquid extraction with methyl-tert-butyl ether (MTBE) [168] along the following protocol. A volume of 100 µl from each sample, as well as from two stripped serum-based and five matrix-based in-house calibrators and three commercial serum control panels were placed into 96-deep-well plates (Corning Life Sciences, Amsterdam, The Netherlands) for sample preparation. Then, 10 µl of a methanolic solution of internal standards was added and incubated for 10 min under slight movement followed by 40 µl 1 M NaCl solution and 5 µl 50 % (w/w) H_3_PO_4_. Extraction was performed with 1000 µl MTBE (VWR, Darmstadt, Germany). After vigorous shaking and centrifugation, 10 min at 3000 g, 600 µl of the upper phase were recovered using a Tecan Genesis robot (Tecan Group, Männedorf, Switzerland) and transferred to another 96-deep-well plate for sample storage. Solvent was removed by vacuum-centrifugation. Then, samples were re-dissolved in 50 µl methanol/water (70/30 v/v) and sealed with chemically resistant sealing mats (Corning Life Sciences, Amsterdam, The Netherlands).

For LC–MS/HR-MS, the system was built with an UltiMate 3000 XRS quaternary UHPLC pump, an UltiMate 3000 RS column oven and an UltiMate 3000 isocratic pump (Thermo Fisher Scientific, Waltham, MA, USA) connected to a PAL HTS-xt autosampler (CTC Analytics, Zwingen, Switzerland) followed by a hybrid quadrupole-Orbitrap mass spectrometer QExactive (Thermo Fisher Scientific, Bremen, Germany) equipped with a heated electrospray ionization source.

A volume of 10 µl from each dissolved sample was injected and separated on a Kinetex 2.6 µm Biphenyl column (50 × 2.1 mm, Phenomenex, Aschaffenburg, Germany) at a temperature of 40 °C. Mobile phase A consisted of methanol/water (5/95; v/v), mobile phase B of 100 % methanol, both containing 0.1 % formic acid and 2 mmol/l ammonium acetate. Gradient elution started at 100 % A with a flow rate of 500 µl/min, followed by a linear increase to 68 % B in 0.1 min, to 71 % B in 2 min and to 82 % B in 2 min. For column cleaning the methanol percentage and flow were increased to 100 % and 800 µl/min within 0.3 min. After flushing for 0.6 min, the solvent composition was changed to 100 % A within 0.1 min and was hold until 5.3 min at a flow rate of 800 µl/min. To minimize contamination of the mass spectrometer, the column flow was directed only from 1.3 to 4.5 min into the mass spectrometer using a diverter valve. Otherwise methanol with a flow rate of 200 µl/min was delivered into the mass spectrometer using an isocratic pump. The ion source was operated in the positive ion-mode using the following settings: Ion spray 3500 V, sheath gas 53, aux gas 14, sweep gas 3 and aux gas heater temperature of 450 °C. Capillary temperature was set to 269 °C and the S-lens RF level to 55.

Data were collected in parallel reaction monitoring (PRM) mode with the following settings: resolution 35000, automatic gain control (AGC) target: 500000, maximum injection time (IT) 100 ms with a multiplex of 2 and quadrupole isolation window of 0.8 *m/z*. Data acquisition and analysis was performed with TraceFinder 3.3 Clinical (Thermo Fisher Scientific, Life Technologies, Darmstadt, Germany), a software module that extracts target ions, generates calibration lines, checks quality controls and ion ratios of quantifier to qualifier ions.

### Axis III: Gut microbiome

#### Overview of microbiome analysis: principles and workflow

##### Microbiome analysis in 12 short steps

A gut microbiome analysis workflow applying next-generation sequencing (NGS) might simply be described in twelve short steps: (1) sampling of stool from participants, (2) (long-term) storage of samples, (3) metagenomic DNA extraction from stool matrix, (4) construction of microbiome-related standards for quantitative PCR, (5) PCR-quantitation of total 16S rDNA for each sample, (6) PCR-amplification of species-specific DNA barcodes, here hypervariable regions within 16S rDNA, followed by (7) composing a sequencing library, including extension of DNA barcode amplicons with technology-specific sequencing adapters and index sequences to distinguish individual samples, multiple PCR-quantification steps for normalization, and pooling of all samples into a single stock, (8) seeding and templating of microbeads to be used for sequencing, i.e. binding of single amplicons from this library to single microbeads, followed by monoclonal amplification of these amplicons at those beads, in order to enable (9) massively parallel sequencing with NGS, followed by (10) bioinformatical cleanup of raw sequencing reads, (11) grouping these reads into similarity clusters and (12) taxonomic assignment of these clusters to bacterial species.

Next-generation sequencing usually refers to second-generation sequencing methods while a fourth generation has already been established, including methods like spatial transcriptomics [169].

Ion Torrent, or Ion semiconductor sequencing technology, as applied here, has been categorized in between second and third generation sequencing methods [170]. It is not based on detection of fluorescent signals like common second-generation NGS-methods but on detection of hydrogen ion release during incorporation of new nucleotides into an extending primer strand.

Many of these roughly described cornerstones require a large body of laboratory methodology and, later, bioinformatical processing, each with several long pipelines assembled together. In our lab, a complete gut microbiome analysis would thus take about six full work days at the bench and two full work days of computational processing (from raw reads to taxonomic assignment including QC validation) if compressed into a single sequential workflow.

##### Certified workflow

The complete gut microbiome analysis workflow, described here, beginning with sample retrieval and storage and ending with taxonomic assignment of bacterial species, has been established in the Institute of Clinical Microbiology and Hygiene at Universitätsklinikum Regensburg, Germany, and is accredited by the German Accreditation Body (DAkkS, *Deutsche Akkreditierungsstelle*, Berlin, Germany) in accordance with the German version of the international quality standard published by the International Organization for Standardization (ISO) DIN EN ISO 15189 [171]. DAkkS accreditation number is D-ML-13084-03-00. It has first been accredited December 9, 2015 and is reassessed every five years, last in January 2025.

Cornerstones of this workflow have first been described [172] in 2016 with deployment of 454 sequencing technology [173] and has since been continuously adapted to incorporate current knowledge, state-of-the-art expertise, and available technology, in particular transitioning to Ion Torrent semiconductor sequencing. Changes to the workflow are reported to the national accreditation body on a regular basis and reassessed accordingly. The state of workflow described here corresponds to summer 2022 during which all microbiome samples of this trial have been processed and sequenced. At the time, standard DIN EN ISO 15189:2014 was current, while it has recently been updated to DIN EN ISO 15189:2022. This is the first time that we publish our certified workflow in methodological detail in a comprehensible manner as part of this study protocol. While samples have been fully sequenced, analysis of sequencing results is pending.

##### Bacterial DNA barcoding with 16S rRNA gene

The central goal in this still most common form of gut bacteriome analysis with next-generation sequencing methods, is so-called bacterial DNA barcoding [174]. This term describes the use of specific DNA marker sequences as universal phylogenetic markers to identify and distinguish individual species. Thus, quantification and taxonomic assignment of bacterial DNA barcodes present in a collected stool sample should enable, as a start, statements on bacterial composition and abundance in the final part of the human gut.

There are several genetic targets that may be used for bacterial DNA barcoding. The one that is most commonly known, the 16S rRNA gene, codes for a component of the smaller (30S) of two subunits of the bacterial (70S) ribosome [175]. Here, the letter “S” denotes Svedberg units that provide a time metric for sedimentation of particles in a fluid and, thus, for their size. For example, 16S describes a sedimentation rate of 16 micrometers per second at one million earth gravities in a centrifuge. Hence, the bigger a particle is, the higher its sedimentation velocity and, thus, the larger its Svedberg unit.

On one hand, 16S rDNA contains multiple highly conserved regions which appear to remain mostly stable across bacterial species throughout evolution; Bacterial PCR amplification primers which bind to these conserved regions are therefore often called ‘universal’ bacterial primers. On the other hand, some of these conserved regions are intertwined with nine so-called hypervariable regions (V1 through V9) that allow for the distinction of different bacterial families, genus and even species and may therefore be used as a DNA barcode [175,176]. However, there has been increasing criticism [177] that extensively applied NGS-methods of today might not be able to account for intragenomic variation within the 16S gene, partly caused by horizontal gene transfer and recombination between bacterial species and, therefore, misinform about species identification. Furthermore, the presence of multiple copy numbers of this gene within a species and various states of chromosome replication might distort abundance estimations. Some of this criticism can be addressed at the work bench as well as during bioinformatical processing, for example by clustering sequence reads into so-called exact amplicon sequence variants instead of previously used similarity cut-off techniques for operational taxonomic units.

##### Overview of microbiome analysis workflow

After having devised and set up a suitable experimental design for microbiome sample generation, a reliable microbiome analysis begins with appropriate sampling, preferably at similar times during common physiological cycles (e.g. time of day, time within menstrual cycle, etc.), without changes in environment, lifestyle and eating habits or irregular intake of other substances and with sufficient knowledge of possible underlying clinical pathologies. Furthermore, conventional, non-invasive rectal stool sampling may not be sufficient or reliable enough for certain research questions such that invasive sampling methods might need to be weighed in. After these considerations, initial stool processing, storage systems for sampled stool and length of storage times are important factors with further impact on analytical results.

In order to receive access to bacterial DNA barcodes like 16S rDNA, very different types of bacterial cell walls first need to be broken open, usually applying a combination of mechanical and chemical methods. This process needs to keep a balance: cell walls from very different types of bacteria need to be sufficiently disrupted while the same process should preserve bacterial DNA as far as possible, i.e. keeping DNA degradation low. Furthermore, DNA extraction should work evenly across all bacterial species within a sample in order to provide for a representative yield of DNA.

Following a reliable and valid extraction of bacterial DNA from stool samples, released nucleic acids need to be purified. After that, multiple steps are required to process the extracted and purified nucleic acids into a so-called sequencing library which can be used for high-throughput sequencing technology.

Two PCR-amplifications form the initial steps of this procedure. Such PCR amplifications can also be seen as ‘target enrichment’: the DNA of interest (target) is identified, and therefore isolated by appropriate primers (fragmentation) and can then be amplified (enrichment). It is helpful to keep in mind that the first is a quantitative PCR and the second are standard PCRs. Each of them is optimized independently for different purposes: qPCR towards accurate quantification, including low PCR error, and standard PCR for accurate coverage. This also explains why different PCR reactions make use of primer sets that appear similar but are not, in detail.

In a first qPCR step, the target is total 16S rDNA for quantification. Universal primers for 16S rDNA are used to quantify all of 16S rDNA copies present among total nucleic acid extracts retrieved from the sample after purification. This quantification enables normalization of originally extracted nucleic acid samples such that they can be diluted to contain equal 16S rDNA copy numbers as input concentrations for PCR amplification of DNA barcodes to follow.

This step also supports quality control of nucleic acid extraction and also may indicate participants with a potentially lower bacterial load. When samples are spiked with control bacteria during sample processing, as they have been here, total 16S rDNA of these bacteria, specifically, is additionally quantified per spike in our lab for quality control of this spiking method.

In a second (non-quantitative) PCR step, the target is a fragment of 16S rDNA – the actual 16S rDNA barcode used as phylogenetic marker. In order to generate a sequencing library, this second PCR amplification step is targeting selected 16S rDNA hypervariable regions which are amplified from the originally purified nucleic acids taken in normalized concentrations (provided by the first PCR) to reduce bias towards samples with higher loads of total 16S rDNA. There are several steps that need to be taken into account for this step.

In contrast to methods like shotgun or whole genome sequencing (WGS), short-read 16S rDNA amplicon sequencing, as used for the majority of NGS methods, typically targets only selected regions of the 16S rRNA gene and therefore fails to capture all nine hypervariable regions present in their full-length sequence. Therefore, a decision needs to be made in advance which of those regions to target in NGS. Each hypervariable region of the 16S rRNA gene offers a different phylogenetic resolution, and the choice of target region is typically guided by desired taxonomic depth and species coverage by deployed primers. While some hypervariable regions allow for higher species-level resolution, others offer broader primer coverage across bacterial taxa, highlighting an inevitable trade-off between specificity and universality in region selection.

Making efficient choices in light of this trade-off depends on additional factors such as the type of samples used for a trial. Moreover, using more than one hypervariable region decimates the number of possible samples within a sequencing run as the number of regions multiply with the number of samples to give the final number of samples in a sequencing run. For example, targeting two different regions doubles the number of samples to be sequenced. Therefore, a further compromise needs to be found for the number of samples that should be analyzed within a single sequencing run versus the number of hypervariable regions of interest that should be analyzed within a single sequencing run for best comparability of samples. Hence, we have primarily selected two hypervariable regions for analysis in parallel for this trial: V3–V4 for a focus on genus-level identification and V1–V3 for potentially better species detection. Moreover, V1–V3 was shown to provide at least reasonable approximation of 16S diversity [178].

For this trial, the second PCRs for 16S rDNA hypervariable regions have been performed with pairs of long fusion primers. In our lab, we have designed 192 individual oligonucleotide fusion PCR primers for each hypervariable region of interest such that a maximum of 192 different samples can be prepared for a single sequencing run. These fusion primers contain four types of nucleic acid sequences. Foremost, all of them contain the same primer sequences necessary to anneal to conserved regions of 16S rDNA located in between the hypervariable regions of interest. Furthermore, all of them contain the same method-specific adapter sequences at each end: at one end, a linker sequence is used later to ensure attachment to solid phase particles that are infused into the sequencing chip and the adapter at the other end is the initiating point for sequencing. The large number of fusion primers is caused by the use of 192 different index sequences that enable the distinction of each sample. A list of recommended index sequences as well as adapter sequences are provided by the manufacturer of the sequencing device.

After each hypervariable region has separately been amplified per sample, amplicons from samples of both hypervariable regions are pooled (and normalized) at equal volumes (i.e. first multiplexing step) and purified using solid-phase reversible immobilization (SPRI). Each pooled amplicon sample is then quantified by qPCR and normalized to achieve equimolar ratios among all samples. Finally, all normalized pools are then combined into a single sequencing library (i.e. second multiplexing step).

Amplicons from this library are bound to solid phase particles for monoclonal amplification (i.e. templating). These templated microbeads can then be infused into a semiconductor sequencing chip for massively parallel sequencing.

A microbiome analysis requires quality control at many stages. It is therefore integrated as explicit or implicit quality control measures.

At the bench, workflows require validation with appropriate controls, such as the use of well-chosen mock communities or by addition of spike-in microorganisms at known quantities to complex samples such as real stool. The least for quality control is to continuously monitor plausibility of intermittent results.

For this project, each sample contains defined amounts of spike-in bacteria serving as internal controls to monitor amplification efficiency and detect potential bias. Every sequencing run also includes negative external controls to identify contamination and mock community samples as positive external controls to assess taxonomic assignment accuracy. Furthermore, multiple purification steps and quality filtering are intertwined.

For this trial, individual 16S rDNA next-generation ion sequencing runs were designed to contain around 160 to 180 stool samples leading to double the size of overall samples for sequencing with two hypervariable regions of interest included.

#### Remark on scientific method descriptions for molecular biology ‘kits’

Microbiome protocol descriptions that follow will show how strongly dependent a (gut) microbiome investigation with next-generation sequencing is on the deployment of a multitude of molecular biology kits and automated, closed machines, of which both no longer allow for sufficient knowledge, insight or even control by the competent researcher [179]. This is particularly true in regard to templating and sequencing chip preparation, and an expression of a serious ‘black box’ phenomenon in scientific research [180,181] that we would shortly like to remark on in order to embed the protocol descriptions that follow into a scientifically relevant context.

Molecular biology kits [182] of today usually are commercial containers or cartridges with multiple sets of packaged biochemical or biophysical materials accompanied by brief instructions on how to deploy these containers. Many of these cartridges serve as inserts for use in fully or at least partially automated laboratory machines which themselves are often sold by the same industry companies that provide the kits. The words ‘container’ and ‘packaged’ already imply that these kits are mostly non-transparent and lacking detailed information of included materials, compositions and applied procedures.

This remark is not meant to address the commercialization of academic molecular biology research, but the scientific consequences of it that severely affect the scientific process, its traceability and its transparency in publications like this one.

More often than not, molecular biology kits have been developed by researchers based at universities, themselves, in order to simplify and standardize the application of molecular biology techniques and to monetize their efforts in addition to academic salaries. In the 1980s, kits were not common [182] and most likely experienced a massive boost by want for application of the polymerase chain reaction technique [180] and related methods for the interaction with DNA and RNA, as is also required in a contemporary microbiome analysis.

Indisputably, such kits have become essential in many areas of the life sciences. They come with increased ease of use, higher throughput and more robust overall quality in clinical molecular diagnostics. At the same time, the evolution of such kits and related biotechnological products have fostered a by now huge and ever growing world-wide market dominated by a few companies who propagate complex molecular biology methodology like simple commercial cake mixes [180,182] while keeping the applied materials and methodologies in secrecy in order to seemingly protect future patent rights or preserve trade secrets [179,182]. This approach might be acceptable for use of such kits in clinical routine diagnostics if the products were underlying appropriate regulations and, correspondingly, if they were accompanied by legally binding guarantees across their full functionality including corresponding enforceable rights. However and in particular for NGS sequencing, the majority of robots and kits rather tend to be classified with standard disclaimers like ‘For Research Use Only (RUO). Not for use in diagnostic procedures’ and, thus, elude such necessary regulations. This appears duplicitous and intensely collides with research in academia which requires background knowledge and transparency for scientific craftsmanship to work.

We have not quite yet reached a state as predicted by well-known biophysicist, Nobel laureate and former Biogen-CEO Walter Gilbert more than three decades ago: “many of the techniques of molecular biology will very soon leave the research laboratories entirely. We will purchase them externally as services; they will not be performed by research scientists.” [180,183], but we appear to be quite close. Attitudes like the one expressed by Walter Gilbert, a man with academic origins in theoretical physics and a forethinker for the human genome project, were probably meant to support the heart of science and to free researchers from cumbersome laboratory work such that they could focus their mental capacities on the results provided by those kits, automated machines and even externalized services. However, this collides with the important principle that a research scientist needs to know and understand the details of the methods applied in order to be able to evaluate them correctly such that results can be published with scientifically neutral reliability.

From our perspective today, however, the industrial development towards a substantially increasing degree of ‘black boxes’ containing complex molecular biology materials and procedures has caused major impediments to the scientific process [179]. Moreover, the high-held industry needs for secrecy appears peculiar as most ‘embodiments’ in patents are mostly based on decades of published academic research by thousands of excellent researchers which in turn often are not even properly referenced.

A major consequence of this phenomenon are published phrases in method sections of scientific research articles like “this kit was employed according to the instructions by the manufacturer”, often without specifying the deployment of the kit, related procedures, components and selected parameters any further. In this way, research scientists tend to participate in a non-scientific process by no longer being able to state the specific materials, compositions and procedures of the deployed kits and in consequence not being able to properly validate them.

Furthermore, whoever has ever read or even followed a technical guideline for a molecular biology kit knows that ‘manufacturer instructions’ usually do not provide for a unique linear algorithm of use as implied by statements mentioned above. In the opposite, it is rather common that manufacturers usually provide different conditions under which a kit can be used and different algorithm pathways and workflows, sometimes depending on preliminary pilot results, intended tasks and applied materials for investigation at hand. Moreover, regular handling of such kits in practice additionally often leads to empirical knowledge on practical details beyond user guides on how to use them most optimally and which is certainly not written into method descriptions.

For the reasons described above, we have put substantial effort into our method sections below, in order to at least describe methodological cornerstones, underlying principles, relevant workflows and original scientific publications. Whenever scientific publications did not seem to entirely fit the method at hand, we refer to patents [179]. While patents contain some of the underlying methodology, they mostly hide the actually applied methods in vast arrays of possibilities and described conglomerations of materials and procedures.

#### Stool sampling

Gut microbiome is dynamic, reactive and showing cyclical changes. Thus, participants were asked to provide a fecal sample in the mornings between 06:00 and 10:00 hours whenever possible. Before, the screening process had already verified that participants would be willing to keep their diet unchanged and stable during the trial.

Patients and healthy participants were personally trained by a study nurse with experience in stool sampling either directly on ward for participating inpatients or at a dedicated neighboring research facility for healthy participants to properly deploy and apply a dedicated sterile paper slip for stool collection (Süsse Labortechnik, Gudensberg, Germany; catalog no. S1000-150) on a toilet seat in a restroom a few walking steps away. Therefore, a study nurse was immediately available to directly assist with procedures when patients required further practical guidance. Stool sampling training included mounting of the sterile paper slip on a toilet set, transfer of an appropriate amount of stool from the paper slip into transparent feces tubes with included spoon (76×20 mm, Sarstedt, Nümbrecht, Germany; catalog no. 80.734.001) as used for common clinical stool diagnostics. This tube was immediately handed over to a study nurse for storage preparation and freezing within 20 min.

Patients provided 147 fresh frozen samples for three time-points at study days 2, 11 and 60 (60 – 58 – 29 samples) as well as 155 (56 – 53 – 46) and 77 (26 – 26 – 25) samples for OMNIgene.GUT and MaGix PBI storage kits, respectively, and as described below. All of the 25 healthy participants provided samples for all three of the storage methods. Two healthy participants provided a second stool sample by discretion of the study physician. This made for a total of 460 stool samples for all applied non-buffering and buffering methods.

#### Sample storage

Proper fecal sample storage is important for a reliable and reproducible microbiome analysis. Stool samples contain, among others, food remnants and digestive enzymes, as well as microbial and scaled off intestinal cells, all of which cause changes in samples before and after defecation. The goal of proper sample handling is to ideally stop these biological processes once a sample leaves the gut and additionally, to protect the sample from chemical processes caused by exposition to atmospheric conditions. Until today, immediate freezing of samples to -80 °C remains a ‘gold standard’ for sample storage [184]. For this reason, in this trial, stool storage had originally been planned as immediately storing unmodified fresh frozen stool at -80 °C. While doing so, it is of note that freezing causes relevant mechanical shear to the samples and long-term frozen storage may therefore cause significant changes to samples [185].

Moreover, frozen sample storage is often infeasible, for example in large projects and in outpatient or field collections. Therefore, many dry and liquid storage methods have been tried for stool preservation at room temperatures in order to increase feasibility and quality of microbiome analyses [184,186–188]. Some of the dry preservation methods are mere drying, for example on fecal occult blood test cards, or with dry swabs for forensic pathology. Chemical recipes for some of the liquid preservation methods may contain 95 % ethanol [189], or sodium hydroxide [190], or (Tris-)EDTA [191], or homebrewed nucleic acid preservation buffers [192], like RNAlater which are mainly based on ammonium sulfate. Commercial products seem to apply similar chemical considerations. It remains unclear what storage solution is ‘the best’, but commercially available OMNIgene-GUT DNA stabilization kit (DNA Genotek, Ottawa, Canada; catalogue no. OM-200) appears to have reached a larger spread by now. However, costs for commercial kits like these still motivate researchers to investigate homebrew solutions.

A preservation medium in which stool may be stored at room temperature for several days should fulfill certain criteria: in the least, bacterial growth, which varies between species and therefore may impact compositional analyses, should be blocked. Furthermore, all DNA contained in a sample should be stabilized, partly by inhibiting DNA degradation that may be caused by enzymes like nucleases or by chemical processes like oxidation or hydrolysis. Thus, well working preservation should, on one hand, be reliable and provide for the same results among replicates of a homogenized stool sample (i.e. technical reproducibility) and, on the other, it should be valid and ideally preserve and represent the bacterial composition and quantities of the original sample.

When this trial was initiated a few years after conception, OMNIgene-GUT had shown first promising preservation results with higher feasibility [193] than immediate freezing. At the same time, our own development for liquid stool preservation [194] was under validation and prepared for scaled production. Hence, we included OMNIgene-GUT about half a year into our trial, and MaGix PBI (microBIOMix, Regensburg, Germany; catalogue no. mgx02.04) was added about one and a half years later. This parallel storage approach has been introduced to directly compare the feasibility and quality of these three storage methods in daily work routine. An amendment with our institutional ethics committee was not required for this technical addition as it did not affect the stool sampling procedure in any way. By the time of writing this study protocol, a variant of OMNIgene-GUT had evolved towards being granted a *de novo* authorization from the U.S. Food and Drug Administration [195].

The safety data sheet for OMNIgene-GUT describes the contents of the buffering solution as 10–30 weight % of ethanol and 1–5 weight % of sodium hydroxide (corresponding to 0.25–1.25 M NaOH). In context of preservation, ethanol prevents bacterial growth and inhibits metabolism [196], while sodium hydroxide helps in rapid lysis of various bacterial cell walls and inactivation of degrading enzymes [190,197]. Furthermore, it provides a first step for DNA extraction [198]. In contrast to preservation media with 95 % ethanol, this solution freezes above -20 °C which might be relevant when comparing this method to freezing non-buffered samples.

Similar to OMNIgene-GUT, MaGix PBI storage solution contains a total of 4 ml of an alcohol-based buffer (20 % v/v ethanol) designed to efficiently stabilize microbial nucleic acids. It also includes a spoon for collecting approximately 1 gram of stool and a 5 mm stainless-steel bead to ensure thorough mixing.

The buffering solution contains bromothymol blue as pH-indicator (stable color in pH-range of 8.0-12.5). Its pH is adjusted to 11.5 with sodium hydroxide (1 % w/v).

Hence, fresh stool that had carefully been collected in large feces tubes was distributed over three storage kits within 20 minutes after sampling without homogenization of the original full sample. Afterwards, two aliquots were prepared for each storage method. Unmodified fresh stool was given into 1.8 ml cryo tubes (CryoPure, Nümbrecht, Germany, catalogue no. 72.379.002) for immediate freezing and the two buffer kits were filled with stool in their accompanying spoons and vigorously shaken by hand until proper buffer immersion. Once kits were properly filled and mixed according to manufacturer guidelines, they were opened and their content was given into two cryo tubes each with sterile spatulas. All of the six aliquots were immediately frozen at -80 °C until analysis. There were no freeze-thaw cycles until the day of bacterial DNA extraction.

#### Construction of external standards for absolute quantitative PCR

##### Principle of standard production for quantifying total bacterial load and spike-in bacteria

External quantification standards for absolute quantitative PCR of either total bacterial load in fecal samples or of total bacterial load in pure spike-in bacterial cultures had been constructed in our lab with the standard-curve method [199] and are continuously being used as internal references for qPCR quantification of bacterial 16S rDNA copies in nucleic acid extracts.

Comparable PCR amplification efficiencies for reference and sample material are key for reliable quantification. Proper PCR amplification is an exponential enzymatic process and, therefore, small differences in PCR efficiency could lead to large differences in PCR product yield and, thus, to relevant discrepancies between standard and sample of interest [199]. Hence, choosing appropriate reference genes for calibration is essential for a reliable quantification method. For this reason, plasmid standards for quantifying 16S rRNA gene copies were generated either from complex bacterial mixtures (i.e. real fecal material, e.g. from humans, mice or rats which had been spiked with bacterial species or not), or from cultured spike-in bacteria as detailed below. The preparation and processing of spike-in bacterial material for standard construction is described at the end of this section.

##### Isolation and amplification of long 16S rDNA fragments

Total nucleic acids were isolated as described in section ‘Isolation and purification of nucleic acids’, below, either from 250 µl of cultivated bacterial species suspended at a turbidity of McFarland Standard No. 2.0 [200], corresponding to about 6×10^8 cfu/ml (colony-forming units per milliliter), or from 50 mg of stool material suspended in 250 µl of buffering solution from a single healthy human donor. McFarland 2.0 turbidities for each bacterial strain were measured from bacterial suspensions in sterile PBS buffer with a densitometer (Densimat, bioMérieux, Nürtingen, Germany). Primers and reaction parameters were identical for both, cultured spikes and fecal material.

Nearly full-length 16S rRNA gene fragments were amplified by PCR using primers S-D-Bact-0008-a-S-16 and D-Bact-1492-a-A-16 (Table 4), as repeatedly being used for the last 30 years and (also known as 27F/1492R or GM3/GM4, [201]) and as recommended by Klindworth et al.[202].

**Table 4:**
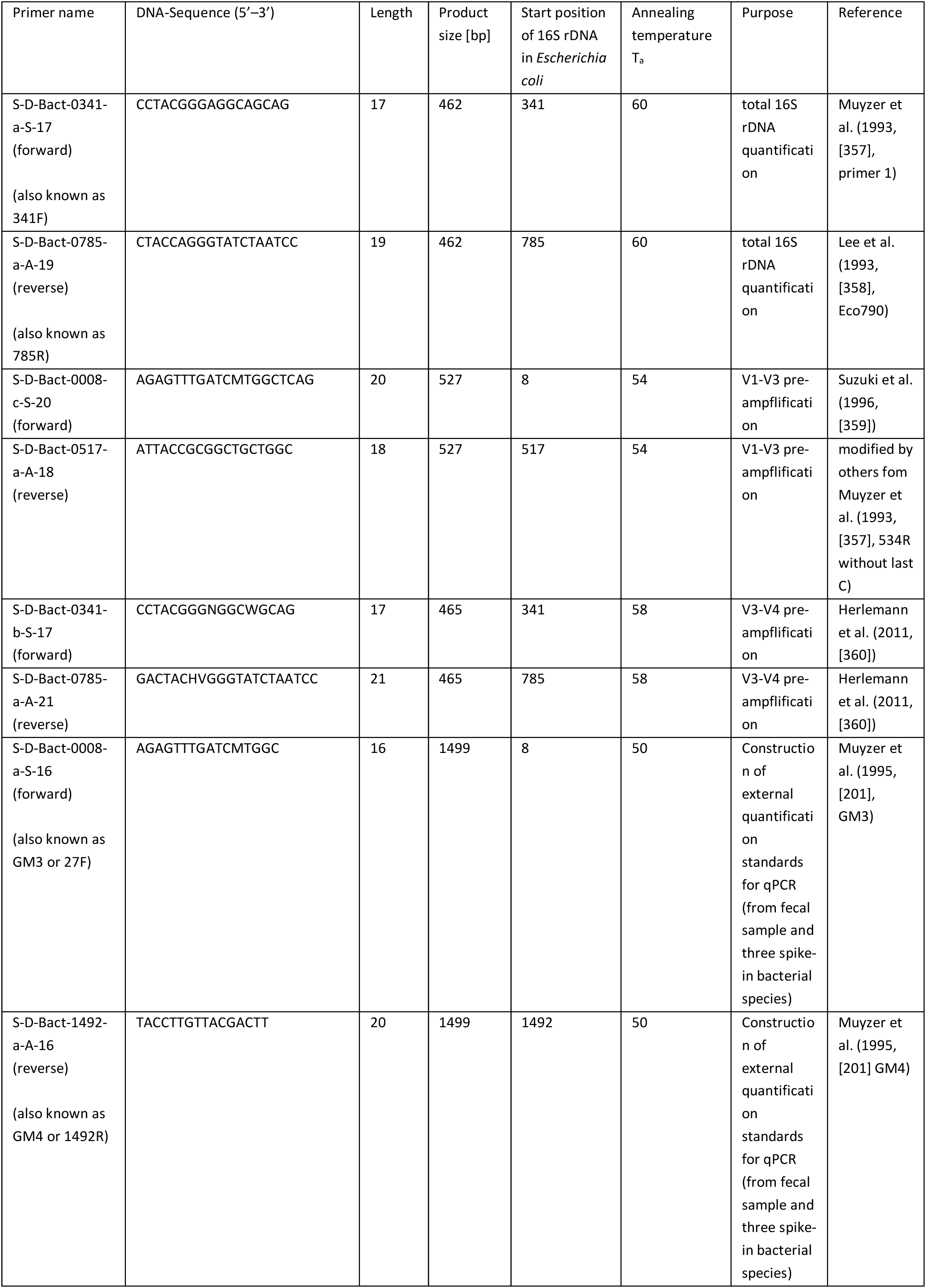
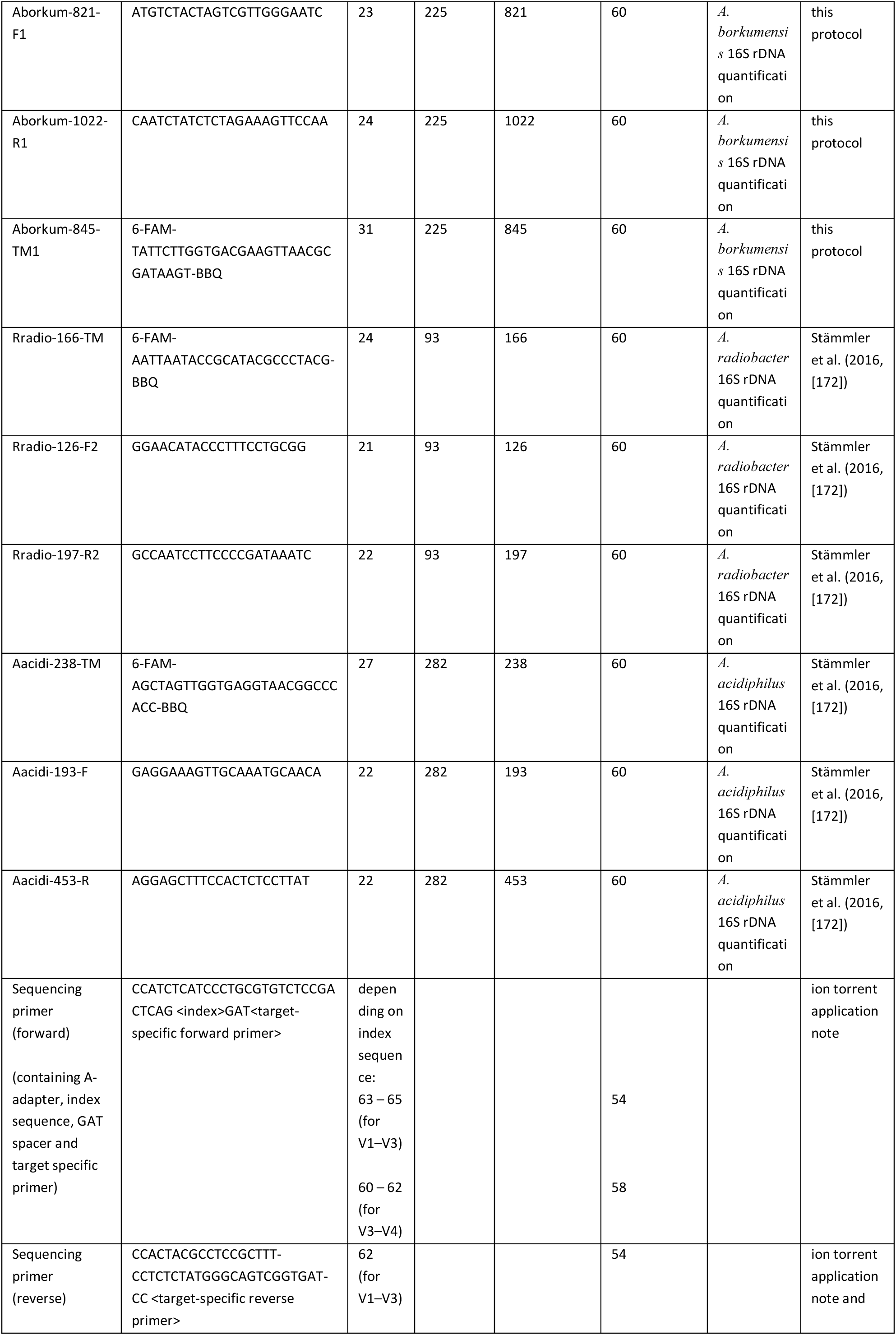
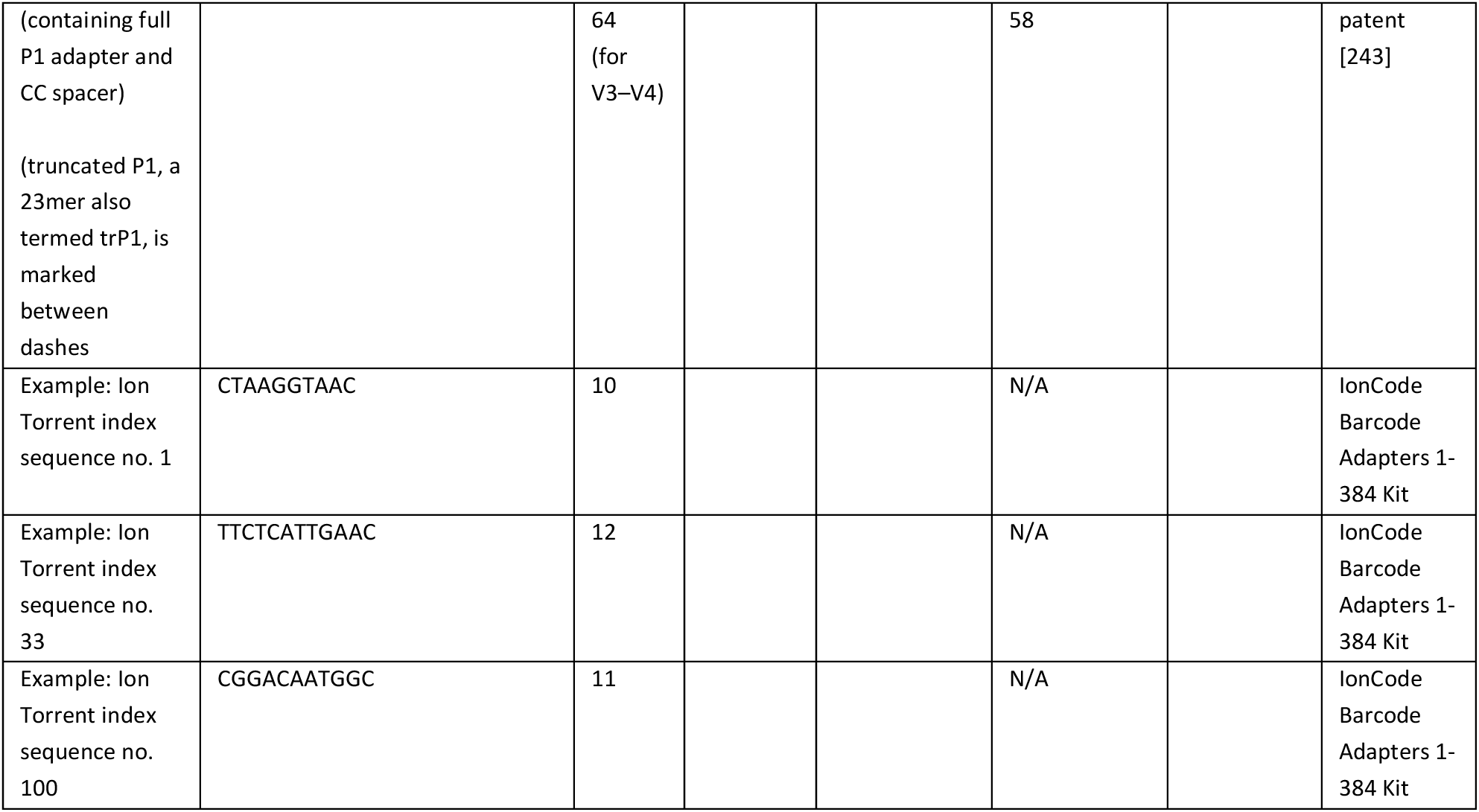
Primers for PCR and sequencing in microbiome analysis. Degenerate primer variants were used for amplicon sequencing to improve taxonomic coverage, while non-degenerate versions were used for quantification to ensure consistent qPCR performance. Degenerate bases follow IUPAC nomenclature (Y = C/T, R = A/G, W = A/T, S = G/C, K = G/T, M = A/C, B = C/G/T, D = A/G/T, H = A/C/T, V = A/C/G, N = A/C/G/T).

Primers were synthesized as Ultramer DNA oligonucleotides in 4 nmol-scale and delivered in a concentration of 100 µM in so-called IDTE buffer (10 mM Tris, 0.1 mM EDTA, at pH 8.0) by IDT (Integrated DNA Technologies, Leuven, Belgium).

They were deployed in Platinum II Hot-Start Green PCR Master Mix (Thermo Fisher Scientific, Darmstadt, Germany, catalog no. 14001013) in which each reaction was set up in a final volume of 50 µl that contained 10 pmol of each primer and 2 µl of template DNA. The master mix provides 1.5 mM MgCl_2_ in its final reaction concentration and was prepared in a final reaction volume of 50 µl, with 22.6 µl PCR-grade water, 0.2 µl of each primer (final concentration 0.4 µM), 25 µl reaction mix solution, and 2 µl of nucleic acid DNA.

The PCR protocol included an initial denaturation step at 95 °C for 3 minutes, followed by 30 cycles of 60 seconds at 95 °C, 15 seconds at 50 °C (annealing), and 45 seconds at 68 °C (elongation), concluding with a final extension at 68 °C for 10 minutes in a Verity PCR thermocycler (Thermo Fisher Scientific, Waltham, MA, USA).

PCR amplicon quality and size was verified with agarose gel electrophoresis in order to confirm expected product sizes and absence of primer-dimers or non-specific amplification products. The applied procedure is described in detail in section “PCR amplification of 16S rDNA variable regions per sample including P1/A-adapter preparation”, below.

Electrophoresis results showed that PCR amplification yielded products of approximately 1500 bp 16S rDNA fragments, with minor length variations due to intrinsic sequence polymorphisms. After PCR cloning, these fragments were quality controlled with Sanger sequencing as described below.

PCR amplicons were purified with solid-phase reversible immobilization (SPRI) using MagSi-NGSprep Plus beads (Steinbrenner Laborsysteme, Wiesenbach, Germany) according to a left side size selection protocol for removing short DNA fragments and primer dimers by choosing a 1.8:1 bead-to-DNA ratio and washing with 200 µl EMSURE-grade 85 % ethanol (Merck, Taufkirchen, Germany) in 96-well full-skirted PCR plates (Sarstedt, Nürmbrecht, Germany; catalog no. 72.1980.700). Purified amplicons were eluted in 50 µl nuclease-free water.

##### PCR cloning of fecal and spike-in bacterial amplicons

For construction of quantification standards, these purified PCR amplicon mixtures were further strongly amplified by insertion into plasmids for cloning by bacteria. PCR cloning like this comes with several helpful advantages in regard to production of standard material and its preservation over longer periods of time. Firstly, using circular DNA in plasmids as quantification standards has been shown to work well as a reference for quantifying linearized DNA of unknown PCR amplicons [203]. Second, it is easy and fast to use for repeated replication whenever standard material is required. Third, this method captures amplicon species in rare abundances well. Fourth, plasmids are known to be more stable in frozen storage than bare amplicons and therefore, fifth, assist in preserving the reliability of quantification standards while in long term frozen storage [204]. Sixth, PCR cloning provides for massive amplification towards very high copy numbers far beyond PCR, and these very high plasmid concentrations are desirable for beginning a broad dilution series in context of the standard curve method.

Therefore, the mixture of 16S rDNA amplicons from a human stool sample and individual spike-in bacteria were cloned with a ready-made kit, pGEM-T Easy (Invitrogen, Thermo Fisher Scientific, Waltham, MA, USA) for so-called TA-cloning [205] following the instructions by the manufacturer. pGEM-T plasmids are derivatives of pGEM-5zf(+) [206,207] with insertion sites additionally flanked by BstZI, a restriction enzyme recognition site. On one hand, Taq DNA polymerases, like the one described for PCR reactions here, add a single deoxyadenosine (A), at the 3’-end of the amplified fragments independent of the amplified template [208]. On the other hand, T-vectors are linearized plasmids with unpaired 3′-T(hymidine)-overhangs at both ends that employ this feature of Taq DNA polymerases in order to match the A-overhangs of the template inserts (i.e. TA-cloning). This enables direct ligation of PCR amplicons into plasmids with a T4 DNA ligase without the need of further enzymes [206]. A 1:1 molar ratio of plasmids to amplicons was used for cloning. In this kit, *Escherichia coli* K-12 derivative JM109 [209] cells with high transformation efficiency for cloning (greater than 1×10^8 cfu/µg DNA) are then used for bacterial transformation [210], i.e. insertion of prepared plasmids into bacterial cells for cloning. The bacteria are made competent for this transformation by thawing them to only 0 °C, keeping them at this temperature on ice in the presence of plasmid buffer with an intermittent gentle and short heat shock at 42 °C [210] for one minute, before incubating them with SOC medium (‘super optimal broth with catabolite repression’) for one hour in an Eppendorf ThermoMixer C (Eppendorf, Hamburg, Germany) at 900 rpm. They were then plated on lysogeny broth (LB) agar plates containing ampicillin at a concentration of 100 µg/ml. Plates were incubated at 37 °C overnight in an incubator (B10, Memmert, Schwabach, Germany).

A vector of this kit allows for a range of insert sizes of 0.5 to 3 kilobases. Ideally, one plasmid should contain a single 16S rDNA amplicon from the complex 16S rDNA mixture with a length of around 1500 bp after transformation. While it is possible that more than one amplicon is inserted into a single plasmid, it does not occur frequently with inserts as large as these. After PCR cloning, Sanger sequencing as described below offers exact sequence identification of these inserts, allowing for selection of plasmids which only contain single and correctly inserted PCR fragments.

High transformation efficiency still means that about 5–10 % of grown bacteria will not contain a plasmid with inserted target. Detection of bacteria with properly inserted amplicons in plasmids is enabled by so-called ‘blue-white’ screening which is genetically made possible via α-complementation of inactive mutant β-galactosidase [211] such that white bacterial colonies indicate transformed cells with included targets in contrast to blue-colored colonies.

##### Quantification of cloned plasmids

Either five (for each spike-in bacterium) or thirty (for fecal amplicons) ‘white’ colonies were selected from a total of about 200 (for spike-in bacteria) or 100 (for fecal amplicons) ‘white’ colonies per plate. For fecal amplicons, more colonies were picked for QC sequencing than used later in case non-valid products were present (e.g. more than one inserted amplicon). Every picked colony was individually grown in 5 ml LB broth supplemented with 100 µg/ml ampicillin (overnight at 37 °C) under vigorous shaking at 200 rpm (KS 3000 i, IKA, Staufen, Germany).

Plasmids were extracted and purified from 1 ml of each individual LB culture using peqGOLD Plasmid Miniprep Kit I (VWR International, part of Avantor, Ismaning, Germany, catalog no. 13-6943-00) according to the spin protocol instructions by the manufacturer without column equilibration and stored at -20 °C. Culture cell density was not explicitly measured before extraction with this kit as 1 ml usually results in cell counts that are within the recommendation of the kit manufacturer.

Plasmid DNA concentrations and purities were analyzed with a NanoDrop ND-1000 spectrophotometer (Thermo Scientific, Darmstadt, Germany). The NanoDrop instrument does not require traditional sample containment like cuvettes but uses liquid surface tension of the sample itself as natural containment such that microvolumes of around one microliter are sufficient for direct spectrophotometric measurement of nucleic acid concentrations [212,213]. It takes ultraviolet absorbance spectra of nucleic acid samples at 260 nm (A_260_) [214], thus providing input for the Beer-Lambert law in order to calculate molecule concentrations. The common extinction coefficient used for double-stranded DNA is 0.020 per mass concentration (ng/µl) per centimeter light path length yielding 50 ng/µl per A_260_ absorbance unit [215]. Furthermore, purity of isolated DNA can be assessed with an absorption ratio of A_260/280_, with a value of higher than 1.8 often considered to be sufficiently pure.

Plasmid copy numbers per microliter were calculated from NanoDrop-measurements following an equation used by many others [203]: copy numbers/µl = (plasmid DNA concentration × 6.022 × 10²³) / molecular weight. Before, DNA concentration had been converted from ng/µl (NanoDrop) to g/µl, and molecular weight was determined from the exact plasmid sequence in g/mol as analyzed with Sanger sequencing described below.

##### Sanger sequencing of cloned plasmids for exact insert characterization

For recovering an exact plasmid sequence, cloned 16S rRNA gene inserts (of 1500 bp lengths, and therefore of nearly full length) were individually verified for each picked colony culture with fluorescence-based dye-linked primer cycle sequencing by capillary electrophoresis in an ABI PRISM 310 Genetic Analyzer (Applied Biosystems, part of Thermo Fisher Scientific, Foster City, USA). The underlying principle is Sanger dideoxy sequencing. In this variant, each of the four dideoxynucleotide terminators is tagged with a different fluorescent dye and provided as part of the ABI Prism BigDye Terminator v3.1 Cycle Sequencing Kit (catalog no. 4337455).

For use of an optimal template concentration, DNA concentration of plasmid samples was measured with NanoDrop as described above and diluted to a concentration of 60 ng/µl before Sanger sequencing. Extracted and purified plasmids were prepared for this Sanger sequencing by adding 60 ng DNA, 1 µl of BigDye Terminator Ready Reaction Mix and 3.2 pmol of the respective primer in a final volume of 10 µl adjusted with PCR-grade water (Roche Diagnostics, Mannheim, Germany; catalog no. 3315959001).

For sequencing of plasmids in both directions, the T7 promotor sequencing primer (5’-d (TAATACGACTCATATAGGG)-3’) and the SP6 promoter sequencing primer (5’-d (ATTTAGGTGACACTATAG)-3’) (both synthesized and HPLC-purified by Biomers.net, Ulm, Germany) were used. Samples were amplified in a PCR thermocycler with the following program: initial denaturation at 95 °C for 15 min, followed by 22 cycles of 95 °C for 1 min, 57 °C for 1 min, and 72 °C for 1 min, with a final elongation at 72 °C for 30 min.

The product of each thermal polymerase reaction cycle was injected and drawn into the instrument capillary by an electrokinetic injection at 2 kV for 75 s. This uncoated capillary (41 cm x 50 µm) was filled with POP-4 (Applied Biosystems; catalog no. 4393715), a so-called performance-optimized polymer no. 4 (an acrylamide-urea based polymer) and heated to 50 °C.

Electrophoretic analysis was carried out at 15 kV and 32 °C for 28 min per sample. Instrument control was performed with the ABI Prism 310 Data Collection software (version 3.1). Sequences from both directions (T7 and SP6) were then assembled and electropherograms verified in SeqMan Pro software (version 7.1, DNASTAR Lasergene, Madison, WI, USA). Consensus sequences were analyzed using BLASTn against the nucleotide collection (nr/nt) database to confirm bacterial species identity based on highest sequence similarity (>99% identity). Additionally, sequence length and alignment coverage were examined to verify single-insert clones and exclude potential concatemers.

Sanger sequencing provided for total plasmid lengths of 1465–1505 bp for transformed fecal amplicons and 1475–1492 bp for transformed bacterial spike-in amplicons. For spike-in bacteria, calculated molecular masses of plasmids were 1409371.2 /mol for plasmids from *A. borkumensis*, 1391503.6 g/mol for *A. radiobacter*, and 1407011.4 g/mol for *A. acidiphilus*, respectively.

##### Construction of standard curves for quantification standards

After sequence identification with Sanger sequencing for calculation of molecular weights, individual cloned plasmid extractions from fecal amplicons were pooled afterwards at equimolar ratios (10 femtomoles each) and filled up to a total volume of 500 µl with TE buffer (10 mM Tris, 0.1 mM EDTA, at pH 8.0).

Each colony consists of clonal offspring of a single transformed original bacterial cell. Therefore, all bacteria taken from one colony should contain a single amplified 16S rDNA amplicon which was confirmed during Sanger sequencing. Hence, a pooled mixture of individually extracted plasmids from 30 picked colonies with inserted fecal amplicons should contain 30 different single amplicons monoclonaly amplified to construct a sufficiently complex PCR standard. For spike-in bacteria, cloned plasmids were taken from a single colony.

Hence, either 1 (for spike-in bacteria) or 20 (for fecal amplicons) different plasmid preparations, as prepared above, were pooled at equal DNA copy numbers according to NanoDrop concentration measurements. Thus, such a pooled mixture contains 1 or 20 different bacterial 16S rRNA gene fragments in balanced numbers.

After pooling, measured total plasmid concentration was used as input for creating a 10-fold serial dilution series of the cloned plasmid mixture [216], ranging from 1×10^3 to 1×10^7 copies/µl. C_T_ values in each of the ten dilution steps were measured in duplicate with qPCR to construct a standard curve.

Quantification was performed using LightCycler 480 Software (version 3.5.1, Roche Diagnostics, Mannheim, Germany) with the 2^nd^-derivative-maximum-method for C_P_-determination. Standard curves were constructed from plasmid standards in duplicate, generating linear regression equations (C_P_ = m × log₁₀[copies] + b, where m is the slope and b the y-intercept). PCR efficiencies ranged from 90 to 110 % (slope: -3.1 to -3.6, R² ≥ 0.98). Unknown sample copy numbers were calculated as: copies = 10^[(C_P_ - b)/m].

Quantities of unknown PCR amplicons can then be extrapolated from this standard curve. For quantification, both the external standard dilutions and the unknown samples are amplified via the same protocol using the same amplification conditions, as described below.

##### Preparation of spike-in bacteria

###### Selection of spike-in bacteria

Three species of bacteria, which usually do not exist in the human gut microbiome and which are well distinguishable from gut bacteria by 16S rDNA next-generation sequencing, were added to all samples in equal 16S rDNA copy amounts according to a protocol that we have called spikein-based calibration to total microbial load (SCML, [172]). All bacterial strains were purchased from DSMZ culture collection (German Collection of Microorganisms and Cell Cultures, Braunschweig, Germany).

Bacterial strains used as spikes here, were *Alcanivorax borkumensis* strain SK2 *DSM 11573^T^* (*A. borkumensis*, GenBank ID: NC_008260.1), a Gram-positive marine bacterium that is hydrocarbonoclastic, i.e. it uses oil hydrocarbons as its exclusive source of carbon and energy [217]. It contains three rRNA gene copies and contributes to the phylum of Pseudomonadota. It was cultivated at DSMZ in Bacto Marine Broth (Difco 2216, DSMZ Medium No. 514 plus 1 % sodium pyruvate) at 28 °C, delivered in a freeze dried stock and cultivated in our lab under aerobic conditions at 30 °C in the same medium.

*Agrobacterium radiobacter DSM 30147^T^* (*A. radiobacter*, GenBank ID: ASXY01000000.1), formerly called *Rhizobium radiobacter*, is a Gram-negative, non-phytopathogenic member of the Biovar I group of *Agrobacterium* found in the soil and the plant rhizosphere [218]. It contains four rRNA gene copies and contributes to the phylum of Pseudomonata. It was cultivated at DSMZ in nutrient agar (DSMZ Medium No. 1) at 30 °C, delivered in a freeze dried stock and cultivated in our lab under aerobic conditions at 30 °C in Luria-Bertani medium (DSMZ Medium No. 381).

*Alicyclobacillus acidiphilus* strain TA-67 *DSM 14558^T^*(*A. acidiphilus*, GenBank ID: PRJDB697) is a Gram-positive, thermo-acidophilic, endospore forming soil bacterium [219]. It contains six rRNA gene copies and contributes to the phylum of Bacillota. It was cultivated at DSMZ in Alicyclobacillus medium (DSMZ Medium No. 402, pH 4.0) at 45 °C, delivered as freeze dried stock and cultivated in our lab under aerobic conditions at 37 °C in the same medium.

###### Storage, culture and quality control of spike-in bacteria

All bacterial cultures were transferred into a microbial cryopreservation system (Cryobank, Mast Diagnostica, Reinfeld, Germany) and stored in our lab at -80 °C until use. Each Cryobank storage vial contains 25 chemically pretreated beads immersed in a cryoprotectant liquid.

Purchased bacterial cultures were revived after arrival by thawing frozen bacterial cells on an agar plate at room temperature and streaking them out with an inoculation loop after which they were incubated at their optimal growth temperature overnight. After cultivation, species was confirmed from a single colony with MALDI-TOF mass spectrometry (matrix-assisted laser desorption/ionization time-of-flight [220,221]) using a MALDI Biotyper (MALDI Biotyper sirius one IVD System, Bruker Daltonics, Bremen, Germany; catalog no. 1890211). A laser within the instrument vaporizes the sample in order to create ions from large and mostly unfragmented molecules whose time-of-flight after (mass-dependent) acceleration yields a protein mass spectrum. Analyzing the protein composition of bacterial cells in this way can be used for rapid microbiological species identification [222] under one minute.

Samples were prepared on polished steel MBT Biotarget 96 targets (Bruker Daltonics; catalog no. 8280784) using the direct transfer method. Briefly, a single colony was applied directly to the target spot, overlaid with 1 μL α-cyano-4-hydroxycinnamic acid (HCCA) matrix solution (MBT HCCA Matrix, Bruker Daltonics; catalog no. 8255344), and air-dried at room temperature. Each measurement included a Bacterial Test Standard (BTS, Bruker Daltonics; catalog no. 8255343) for calibration. Mass spectra were acquired in positive linear mode over a mass range of 2–20 kDa. Spectral acquisition and analysis were performed using MBT Compass software (version 4.0) with the MBT Library (version 7.0). Species identification was based on log(score)-values, with scores ≥2.0 indicating species-level identification, 1.7–1.999 indicating genus-level identification, and <1.7 considered unreliable, according to recommendations by the manufacturer.

Multiple colonies were then picked with an inoculation loop for transfer into a cryopreservation vial which then was gently mixed and cryoprotectant fluid removed before freezing for long-term storage at -80 °C. When needed, bacterial cultures were revived from this cryopreservation system by removing a single storage bead with a sterile inoculation loop in order to thaw the coated cryopreserved bacterial cells on an agar plate in the same way as described above.

###### Construction of standard curves for spike-in bacterial concentration

Whole bacterial cells from these cultures were to be spiked into thawed fecal samples in predefined concentrations. For this, a single colony from the overnight cultures was picked with an inoculation loop and brought into suspension in 5 ml of sterile PBS buffer. Bacterial concentration in such a suspension was estimated with a SmartSpec 3000 spectrophotometer (Bio-Rad, Neuried, Germany, catalog no. 170-2501). Thereafter, 200 milliliters of such a well-mixed suspension was mixed with 800 µl saline solution (0.9 % NaCl), given into semimicrovolume disposable polystyrene cuvettes (Bio-Rad; catalog no. 223-9955) designed for a light pathlength of 10 mm, and optical density (OD) was measured at 600 nm (OD_600_).

Overnight cultures (i.e. 16 to 20 hours) were harvested in stationary growth phase at time of their analysis and use.

In a preparatory step, conversion factors between OD_600_ and number of bacterial cells per milliliter for each spike-in bacterial species had before been derived from constructing species-specific standard curves by microscopic counting of overnight cultures using an improved Neubauer chamber as hemocytometer (Blaubrand Neubauer improved, Sigma Aldrich, a part of Merck, Darmstadt, Germany; catalog no. BR717810). This counting chamber has a standard depth of 0.1 mm and shows a 3×3 grid of large squares of which each has an area of 1 mm² such that each large square denotes a volume of 0.1 µl. Bacteria were counted across the whole chamber without previous staining.

For each bacterial species, a 10-fold serial dilution was microscopically counted in triplicate in correspondence to OD_600_-measurements in an OD-range between 0.1 and 0.3 OD-units. Bacterial sedimentation and aggregation was prevented by gently vortexing the suspension immediately before measurements.

Thus, in our lab one OD_600_-unit corresponds either to 1.45 x 10^9 cells/ml for *A. borkumensis*, or to 1.40 x 10^9 cells/ml for *A. radiobacter*, or to 1.17 x 10^9 cells/ml for *A. acidiphilus*.

###### Final preparation of spike-in bacterial pool

The goal for an external spike-in bacterial standard was to select bacterial concentrations according to a desired number of 16S rRNA gene copies that should be added per sample for each spike. The ribosomal RNA operon copy number database (rrnDB, [223]) provides a number of genomic 16S rRNA operons per bacterium of a species which was used as reference, here. These numbers were mentioned above (3, 4 and 6) and thus, one OD_600_-unit could be transformed into 4.4, 5.6 and 7.0 billion 16S rRNA gene copies per milliliter for *A. borkumensis*, *A. radiobacter*, and *A. acidiphilus*, respectively.

Spike-in bacteria were pooled together in different concentrations such that they would be present in clearly distinguishable copy numbers per sample after sequencing to enable additional quality control, for example in regard to possible deviations from expected results in qPCR.

Hence, quantified overnight cultures were diluted based on their OD_600_ measurements to achieve predefined 16S rRNA gene copy numbers in a total volume of 60 µl, which was then added to the samples as a spike-in pool: 1 x 10^8 16S rDNA copies per sample for *A. borkumensis*, 3 x 10^8 for *A. radiobacter* and 8 x 10^7 for *A. acidiphilus*.

In our lab, these levels of spike concentrations are selected for stool specimens from patients in whom no gastrointestinal abnormalities are to be expected clinically, as for all samples of this trial – in contrast to clinical trials where participants are likely to have comparably lower bacterial loads in their stool such that spike concentrations need to be lowered to prevent later analytical distortions by spike-in bacteria.

Normalizing the amount of spikes to theoretically known gene copy numbers (as available from rrnDB) is the least that can be done for quality control. However, there is inherent uncertainty in regard to 16S rDNA copy numbers in bacteria as several biological aspects cause additional variation [224]. For example, a bacterium may be in a state of replication and, thus, may currently have a variable and unpredictable number of chromosomal copies present in the bacterium. This variability in possible 16S rDNA copy numbers has triggered a few modelling approaches to address these stochastic variations [225,226] but a debate continues about their feasibility and impact.

#### Amplicon sequencing of bacterial ribosomal DNA and read preprocessing

##### Isolation and purification of nucleic acids

Procedures and sample handling were designed and performed to keep technical variabilities to a minimum on all levels of execution. Fecal specimens were thawed at room temperature, distributed and preprocessed in a class II biosafety cabinet to prevent contamination of samples. All of the 460 samples described above were thawed for the first time immediately before extraction of nucleic acids. They were extracted in eight batches over four consecutive days by a single operator with extensive routine in this operation.

For fresh frozen stool without previous buffering (i.e. native), 50 mg of original fecal material were weighed and suspended in 250 µl of sterile PBS buffer. This corresponds to usual stool concentration in the other storage systems. Then, 250 µl of stool suspension were taken from each storage system, either from native stool in PBS or from fecal samples that had already been suspended in stabilization buffer before freezing. Metagenomic DNA was then isolated from this suspended fecal material in the following steps.

Each variant of buffered stool suspensions was directly given into 2 ml tubes already containing Lysing Matrix Y beads (0.5 mm in diameter, yttria-stabilized zirconium oxide beads, MP Biomedicals, Eschwege, Germany). Then, 250 µl of S.T.A.R. buffer (Stool Transport and Recovery Buffer, Roche Diagnostics, Mannheim, Germany), 60 µl of spike-bacteria pool (whole cells, prepared as described above) containing 1 x 10^8, 3 x 10^8, and 8 x 10^7 16S rDNA copies for *A. borkumensis*, *A. radiobacter* and *A. acidiphilus*, respectively, and 50 µl of Proteinase K (Roche Diagnostics, Mannheim, Germany) were added. S.T.A.R. buffer contains detergents that increase lysis efficiency and nuclease inhibitors that prevent DNA degradation. Additionally, it appears that potential PCR inhibitors appear to be limited by the buffer. Proteinase K was used to liquefy the stool matrix while leaving bacteria intact and to prevent aggregation of bacterial cells in frozen storage.

Tubes with these prepared mixtures were then frozen at -20 °C over night (about 20 h) as a first step of mechanical disruption of bacterial cells. The next day, thawed mixtures were incubated at 65 °C for 10 min for Proteinase K treatment. Proteinase K was then inactivated by heating samples to 95 °C for 5 min and letting them cool to room temperature after. A second step of mechanical disruption continued with bead beating on a TissueLyzer II instrument (Qiagen, Hilden, Germany) for 5 min at 30 Hz with the beads already contained in the prepared mixture. Afterwards, additional 500 µl of S.T.A.R. buffer were added, the whole mixture was vortexed and then centrifuged at 4 °C at 20000 g for 5 min (centrifuge: 5427 R, Eppendorf, Hamburg, Germany) in order to pellet insoluble stool particles and beads.

A MagNA Pure 96 nucleic acid isolation robot (Roche Diagnostics, Rotkreuz, Switzerland, catalog no. 06541089001) was used for automated purification of nucleic acids [227] from 500 µl of supernatant from these stool lysates. A MagNA Pure 96 DNA and Viral NA Large Volume Kit (catalog no. 06374891001) was deployed, applying the provided default LV Blood protocol, according to manufacturer instructions and providing a final elution volume of 100 µl. MagNA Pure 96 including the kit are certified for in vitro diagnostic use and are accompanied by a detailed method sheet including compositions of applied solutions [228]. A single kit is designed to isolate, among others, bacterial nucleic acids from up to 1 ml of lysates of human origin and provides for 3 x 96 isolations as input for consecutive qPCR analysis. The manufacturer calls the methodological basis for nucleic acid isolation as MagNA Pure magnetic glass particle (MGP) technology [229,230]. This kind of silica-binding chemistry uses a chaotropic salt, like guanidine hydrochloride, in high concentrations to enable binding of nucleic acids to silica [231] and it also disrupts cells and inactivates nucleases. Magnetic glass particles with bound nucleic acids can then be separated magnetically from the residual lysis mixture after which these are purified in multiple washing steps to remove unbound substances like proteins, debris from cell walls and PCR inhibitors. In a final step, purified DNA is eluted from the glass beads under low-salt conditions [228,229]. For subsequent PCR quantification of total bacterial and spike-in 16S rDNA copies, eluates were diluted 1:100 in nuclease-free water (Carl Roth, Karlsruhe, Germany).

Following automated extraction, nucleic acid concentration and purity were assessed using a NanoDrop-2000 spectrophotometer (Thermo Fisher Scientific, Wilmington, DE, USA; catalog no. ND-2000). Absorbance ratios at A_260/280_ and A_260/230_ were measured to evaluate sample purity. Samples with A_260/280_ ratios between 1.7 and 2.0 and A_260/230_ ratios larger than 1.7 were considered acceptable for downstream applications.

##### Quantification of total 16S rDNA copy numbers per sample

Two different types of quantitative real-time PCR (qPCR) assays were run for quantification of total 16S rDNA copy numbers: one intercolator-based (SYBR Green) singleplex qPCR for total bacterial load in each sample and three individual singleplex hydrolization probe-based qPCR for the three bacterial strains spiked into each sample. The latter are described in the next section.

Total bacterial 16S rDNA from all extracted and purified nucleic acid samples (including spike-in bacteria) was isolated and quantified in a first step of quantitative qPCR. This step is meant to yield the total amount of bacterial 16S rDNA copy numbers per sample. With this information, 16S rDNA copies for each sample can be normalized to a specified concentration for all samples before further analysis – for example, to one million copies per microliter. For this, an external quantification standard, derived from a healthy human stool sample, was constructed, as described above, and included two times in the qPCR run in a concentration range of 1 × 10^3 to 1 × 10^7 copies/µl.

Real-time PCR amplification [232] was performed in a microvolume fluorimeter with rapid temperature control and temperature homogeneity across 384-multiwell-plates [233,234] by real-time monitoring of SYBR Green I fluorescence for every PCR cycle in each reaction well during amplification (LightCycler 480 II, Roche Diagnostics, Mannheim, Germany; catalog no. 05015243001 (for 384)) with white LightCycler 480 multiwell plates 384 (catalog no. 04729749001). It uses an LED for excitation and a charge-coupled device (CCD) camera to collect fluorescent signals. The LightCycler 480 II instrument is classified as RUO and not intended for diagnostic use.

SYBR Green I [235,236], an intercalating [237] cyanine dye, binds to all available double-stranded DNA in a sample and abruptly increases fluorescence when it does so and can thus be used as a surrogate marker for PCR product accumulation (like many other dyes). Hence, the fluorescent signal yields sigmoidal curves as an expression of an initially exponential PCR enzymatic process and its products and depletion of reactants later. Threshold cycle (C_T_), also known as crossing point (C_p_), marks the point at which the fluorescence signal of a PCR product exceeds (“crosses”) the background level along a characteristic sigmoidal amplification curve. It represents the threshold at which the accumulation of product becomes detectable and correlates to concentration of nucleic acids in a sample such that higher nucleic acid concentrations require fewer cycles to threshold, i.e. left-shifted fluorescence curves. C_T_-values were determined by applying the second derivative maximum method and PCR melting curve analysis was performed as described below.

Bacterial 16S rDNA primer pair S-D-Bact-0341-a-S-17 / S-D-Bact-0785-a-A-19 (Table 4) was used as specific quantification primers for total bacterial 16S rDNA applying a protocol similar as described before [172]. Degenerate primer variants were used for amplicon sequencing to improve taxonomic coverage, while non-degenerate versions were used for quantification to ensure consistent qPCR performance.

Klindworth et al. [202] had evaluated multiple primer pairs for 16S rDNA NGS with Roche 454 pyrosequencing and reached a conclusion that this was one of the best primer pairs in their study as it achieved high bacterial domain and phylum coverage making it a good candidate set for quantitative applications. Though their results referred to Roche 454 pyrosequencing with focus on hypervariable region V3-V4, our experience confirms that this primer pair remains feasible for NGS with Ion Torrent and hypervariable regions V1–V3 and V3–V4.

Concentration of primers, concentration of MgCl_2_ and annealing temperature had been optimized for this set of PCR primers and provided best results for standard MgCl_2_ concentration according to the kit manual and at an annealing temperature of 60° C (unpublished data) using the LightCycler 480 SYBR Green I Master kit.

A qPCR mixture with 20 µl final reaction volume was prepared by using LightCycler 480 SYBR Green I Master kit (Roche Diagnostics, Mannheim, Germany, catalog no. 04707516001). This kit uses FastStart Taq DNA Polymerase with an added inhibitor enzyme acting as a temperature-dependent switch that can be reversibly inactivated [238] by elevating temperature. This enables so-called ‘hot-start’ qPCR such that preparation of reactions can be performed at room-temperature beforehand.

A master mix was prepared with 10 µl of 2× SYBR Green I Master Mix, 1 µl each of forward and reverse primers (final concentration of 1 µM), and 6 µl nuclease-free water. Eighteen microliters of master mix were dispensed per well on a white LightCycler multiwell Plate 384 (calalog no. 04729749001), followed by addition of 2 µl of nucleic acids (1:100 diluted) extracted from stool samples. Quantitative hot start qPCR was performed over 40 cycles (95 °C for 10 s (denaturation), 60 °C for 15 s (primer annealing) and 72 °C for 15 s (DNA extension) with an initial 10 min hot start at 95 °C. Fluorescence was measured at the end of each extension step at 72° C. Presence of non-specific PCR products as detected by SYBR Green I dye was ruled out by melting curve analysis [239] after amplification with a temperature gradient of 0.2° C/s from 65 to 95 °C. Samples were then cooled down to 40 °C for 30 s. Melting curve analyses were performed to confirm melting points of samples and standards between 85 °C and 92 °C.

An external quantification standard is co-amplified in duplicates during qPCR in various concentrations ranging from 1 × 10^3 to 1 × 10^7 plasmid copies/µl as described above in dedicated control wells on the same plate (in contrast to an internal standard that is co-amplified mixed together with a sample in the same well).

Fluorescence curves from the standard in different concentrations yield different C_P_-values that can then be plotted against the corresponding nucleic acid concentration of the standard in logarithmic transformation.

Comparing C_P_-values from samples with unknown amounts of nucleic acid template to a fitted linear standard curve then provides for the original concentration of template per sample before the qPCR was started

Quantification was conducted as described above, using the second derivative maximum method for calculation of C_P_-values, implemented in LightCycler 480 software (version 1.5.1). Quality control criteria for standard curve acceptance included slope values of -3.1 to -3.6 and amplification efficiency of 90 to 110 %.

Results from absolute qPCR quantification of total 16S rDNA per sample were then used for normalization of nucleic acid content in samples. For this, 5 µl from each sample of the original purified nucleic acid mixtures were picked with semi-automated electronic 8-channel pipette (Voyager 2–50 µl, Integra Biosciences, Biebertal, Germany; catalog no. 4726) with low-retention tips (Integra Griptips 125 µl, catalog no. 3727) individually diluted with nuclease-free water to equal concentrations of one million 16S rDNA copies per microliter for each sample in 1.5 ml screw cap micro tubes (Sarstedt, Nürmbrecht, Germany).

Thus, quantification of total 16S rDNA copy numbers per sample enabled the input of equal concentrations of 16S rDNA copies per sample into a second qPCR step for isolating and pre-amplifying hypervariable regions of interest before sequencing.

##### Quantification of spike bacteria-specific 16S rDNA copy numbers in each sample

Specific quantification of spike-in bacteria contained in complex fecal mixtures requires greater specificity than universal bacterial 16S rDNA amplification. Therefore, we implemented a testing approach that incorporated hydrolysis probes alongside amplification primers. The abundance of three distinct 16S rRNA genes from whole-cell bacterial spike-in standards was quantified using this hydrolysis probe-based approach, with calibration against constructed plasmid standard curves for each spike-in bacterial species. Singleplex qPCR assays were performed for each of the three bacterial targets.

Primers and hydrolysis probes for quantification of 16S rDNA from *A. radiobacter* and *A. acidiphilus* had been designed and evaluated *in silico*, before [172], based on the RefNR sequence collection of the SILVA reference database, release 119, containing 534968 different 16S rRNA sequences. Allowing no primer mismatches, specificity of primers and probes targeting *A. radiobacter* and *A. acidiphilus* DNA provided for specificities of 100 %. Specificity of primers and probes had further been evaluated *in silico* using the Nucleotide BLAST (BLASTn) algorithm [240] against the nucleotide collection (nt) database. Primers for quantification of *A. borkumensis* DNA was added later, following an identical approach.

Thus, loads of spike-in bacteria *A. borkumensis*, *A. radiobacter* and *A. acidiphilus* were individually estimated by quantifying species-specific 16S rRNA gene copy numbers within the isolated DNA of spiked samples by qPCR with corresponding species-specific primers and hydrolysis probes on a LightCycler 480 II instrument on white LightCycler 384 multiwell plates in reference to three included external quantification standards of various known 16S rDNA concentrations. The threshold cycle (C_T_) was determined as described above for universal bacterial 16S rDNA. Primer pairs Aborkum-821-F1 / Aborkum-1022-R1 were used for *A. borkumensis*, 126-F2 / 197-R2 for *A. radiobacter*, and 193-F3 / 453-R3 for *A. acidiphilus* in spiked fecal nucleic acid extracts.

Primers for spike-in bacteria were synthesized and HPLC-purified by Biomers.net (Ulm, Germany). Hydrolysis probes were then synthesized at 3 nmol scale by TIB Molbiol (Berlin, Germany) and labeled with 6-carboxyfluorescein (FAM) fluorophore and BlackBerry quencher 650 (BBQ).

Concentration of primers were optimized by titration in the range of the kit manufacturer recommendations after PCR amplification of 16S rDNA targets from DNA extracts of human and murine fecal specimens. Concentration of MgCl_2_ and annealing temperature had been optimized for these three sets of PCR primers and provided best results for 4 mM MgCl_2_ and 60° C.

A qPCR mixture with 20 μl final reaction volume was prepared by using LightCycler 480 Probes Master kit (Roche Diagnostics, Mannheim, Germany, catalog no. 04707494001) which provides a hot-start reaction mix with FastStart Taq DNA Polymerase designed specifically for detecting DNA targets with hydrolysis probes during qPCR with the LightCycler 480 II system.

The final reaction mix contained 4.4 µl PCR-grade water, 1.6 µl MgCl_2_ stock solution (final 4 mM), 0.5 µl of each primer (final 0.25 µM for each), 1 µl for each probe (final 0.1 µM each), reaction mix 18 solution, and 2 µl of template DNA.

qPCR was conducted over 40 cycles (95 °C for 30 s (denaturation), 60 °C for 30 s (primer annealing) and 72 °C for 30 s (DNA extension)) with an initial 10 min hot start at 95 °C.

Quantification was conducted using the second derivative maximum method for calculation of C_P_ values, implemented in LightCycler 480 software version 1.5.1. Quality control criteria for standard curve acceptance included slope values of -3.1 to -3.6 and amplification efficiency of 90-110%.

Individual quantification standards per bacterial strain were added to dedicated control wells in five different concentrations for external co-amplification as described above. After qPCR, template DNA was normalized to 1×10⁶ 16S rDNA copies per microliter by dilution with nuclease-free water in a 96 well-plate.

##### PCR amplification of 16S rDNA variable regions per sample including P1/A-adapter preparation

All procedures described above have laid the foundation such that quantitatively spiked sample material after nucleic acid purification in normalized total 16S rDNA concentrations is available for building a 16S rDNA sequencing library. Library amplicons were generated by isolating and amplifying V1–V3 and V3–V4 hypervariable regions of bacterial 16S rRNA genes from a total of ten million bacterial 16S rDNA copies per sample, as prepared before. Each hypervariable region of interest was amplified in a separate PCR reaction. During these two amplification processes, the amplicon products were additionally prepared for the sequencing system to be used. For Ion Torrent sequencing, this meant extending the ends of sample amplicons by two components: first, index sequences in order to distinguish amplicons originating from a single sample after sequencing, and second, a P1/A-adapter system per amplicon used for Ion Torrent technology from templating to sequencing as described below. No kit was deployed for building the sequencing library.

Hence, 192 individual large fusion PCR primer pairs were designed for Ion Torrent-specific sequencing for each hypervariable region such that a maximum of 192 independent samples could be analyzed in a single sequencing run.

Primer pairs for amplifying V1–V3 and V3–V4 hypervariable regions were verified based on sequence coverage and phylum spectrum evaluation with TestPrime 1.0, an online service of the SILVA project. This service is offered according to the *in silico* PCR primer performance evaluation method published by Klindworth et al. [202] and allows inspection of per-taxon coverages for individual primer pairs.

All evaluations were based on RefNR sequence collection of the SILVA 16S ribosomal RNA reference database (SSU release 119) with no and one primer mismatch allowed and a 0-mismatch zone of 3 bp length at the 3’ end.

Entering V1–V3 primer pair S-D-Bact-0008-c-S-20 (5’- AGAGTTTGATCMTGGCTCAG-3’) / S-D-Bact-0517-a-A-18 (5’-ATTACCGCGGCTGCTGGC-3’) provided 66.6 % matches for region coverage. Entering V3–V4 primer pair S-D-Bact-0341-b-S-17 (5’-CCTACGGGNGGCWGCAG-3’) / S-D-Bact-0785-a-A-21 (5’- GACTACHVGGGTATCTAATCC-3’) provided 94.7 % matches for region coverage.

Primer pairs for isolating and amplifying hypervariable regions were: forward primer S-D-Bact-0008-c-S-20 and reverse primer S-D-Bact-0517-a-A-18 for hypervariable region V1–V3 (Table 4) and primer pair S-D-Bact-0341-b-S-17 (forward) / S-D-Bact-0785-a-A-21 (reverse) for hypervariable region V3–V4. Primers were synthesized as Ultramer DNA oligonucleotides in 4 nmol-scale and delivered in a concentration of 100 µM in so-called IDTE buffer (10 mM Tris, 0.1 mM EDTA, at pH 8.0), by IDT (Integrated DNA Technologies, Leuven, Belgium).

These bacterial forward and reverse primers were extended to large fusion primers by the following modifications (Table 4). Reverse primers were extended by an additional 3’-linker sequence (full P1-adapter, 23+18 bp) such that templates could be attached to the surface of proprietary silica microbeads later (i.e. P(articles)); forward primers were fused with an Ion Torrent-specific index sequence (10 to 12 bp, see examples in Table 4) for later identification of each sample and with a final 5’-sequence (A-adapter, 30 bp) which provides for primer annealing during final sequencing (i.e. A(amplification)). A set of 384 possible index sequences is provided and strongly recommended for use with Ion sequencing technology. For users of Ion Torrent sequencers, these linker sequences can be found in the Ion Torrent Browser (also see: IonCode Barcode Adapters 1-384 Kit, Thermo Fisher Scientific, Darmstadt, Germany, catalog no. A29751).

Length of forward primers was variable because of variable length of index sequences. Hence, these long fusion primers had a total oligonucleotide length of 63 to 65 (V1–V3) and 60 to 62 (V3–V4) for forward and 62 (V1–V3) and 64 (V3–V4) for reverse primers.

Amplicon sequencing with these long fusion primers was performed according to the following protocol: concentration of primers, concentration of MgCl_2_, and annealing temperature had been optimized for this large set of long PCR fusion primers and provided best results for 1.5 mM MgCl_2_ and 54° C (V1–V3 primers) or 58 °C (V3–V4 primers), respectively.

A qPCR mixture was prepared with Platinum II Hot-Start Green PCR Master Mix (2X) (Thermo Fisher Scientific, Darmstadt, Germany, catalog no. 14001013) according to the 3-step protocol. The master mix provides 1.5 mM MgCl_2_ in its final reaction concentration and was prepared in a final reaction volume of 40 µl, with 8 µl PCR-grade water, 1 µl of each primer (final concentration 0.5 µM), 20 µl reaction mix solution, and 10 µl of template DNA (DNA normalized to 1×10⁶ 16S rDNA copies per microliter according to total 16S rDNA qPCR quantification results). This hot start qPCR was performed over 30 cycles (94 °C for 30 s (denaturation), 54/58 °C for 45 s (annealing) and 68 °C for 45 s (extension) with an initial 2 min hot-start at 94 °C and a final extension step at 68 °C for 5 min in a Verity PCR cycler (Thermo Fisher Scientific, Darmstadt, Germany) using twin.tec PCR Plate 96 (Eppendorf, Hamburg, Germany).

PCR amplicon quality was assessed by agarose gel electrophoresis in order to confirm expected product sizes and absence of primer-dimers or non-specific amplification products. Final PCR products were directly loaded onto a 1.5 % (w/v) agarose gel (Sigma Aldrich, part of Merck, Darmstadt, Germany; catalog no. A9539) with a DNA-gel loading dye (Thermo Fisher Scientific, Life Technologies, Darmstadt, Germany; catalog no. R0611) together with a DNA ladder standard (GeneRuler 100 bp, Thermo Fisher Scientific, Life Technologies, Darmstadt, Germany; catalog no. SM0243). Electrophoresis run time was 40 minutes at an electrical potential of 6000 mV/cm in 1x TBE as running buffer (diluted from 10x TBE, Sigma Aldrich, part of Merck, Darmstadt, Germany; catalog no. 574795).

Ethidium bromide 1 % solution (Carl Roth, Karlsruhe, Germany; catalog no. 2218.2) had already been added to the agarose gel at a concentration of 0.5 µg/ml for in-gel staining of amplicon bands. Ethidium bromide is a DNA intercalating fluorescent dye that can be visualized with an ultraviolet (UV) light source in a UV transilluminator (Gel Doc EZ Gel Documentation System, Bio-Rad Laboratories, Feldkirchen, Germany; catalog no. 8212).

Amplicon lengths were estimated in relation to added DNA ladder standard. Electrophoresis results showed that PCR amplification yielded products of approximately 631 bp for V1–V3 and 571 bp for V3–V4 for the respective 16S rDNA fragments, with minor length variations due to intrinsic sequence polymorphisms.

Individual V1–V3 and V3–V4 PCR reactions (20 µl each) were then pooled for each sample with a semi-automated electronic 8-channel pipette (Integra Voyager 2–50 µl) with low-retention tips (Integra Griptips 125 µl). Using equal volumes was sufficient for normalization of both target regions.

This pool was then purified and filtered for amplicon size (see below) with solid-phase reversible immobilization (SPRI) using MagSi-NGSprep Plus beads (Steinbrenner Laborsysteme, Wiesenbach, Germany) with a manual 8-channel pipette (Rainin Pipet-Lite Multi Pipette L8-200XLS+, Mettler-Toledo, Giessen, Germany; catalog no. 17013805/ with Pipette Tips TR LTS 200µL F 960A/10; catalog no. 17014963)) according to a left side size selection protocol for removing DNA fragments below a target size of about 300 bp by choosing a 1.2:1 bead-to-DNA ratio and washing with 200 µl EMSURE-grade 85 % ethanol (Merck, Taufkirchen, Germany) in 96-well full-skirted PCR plates (Sarstedt, Nürmbrecht, Germany; catalog no. 72.1980.700). Purified amplicons were eluted in 50 µl nuclease-free water.

##### Composing the sequencing library from all samples and variable regions

A ‘sequencing library’ constitutes the final stock solution of preprocessed PCR amplicons that have been pooled together from all experimental samples and targeted genetic regions. Preprocessing, as described above, includes ligation of amplicons with adapter and index sequences for each sample of each genomic target region as well as size selection, purification and quality assessments. Furthermore, concentrations are normalized for each sample and each targeted genomic region in order to achieve an equimolar representation across all samples and regions such to prevent bias during sequencing later.

Precise absolute quantification is a critical prerequisite for accurate sequencing library composition after PCR amplicons for hypervariable target regions have been purified, filtered in size and controlled for quality. This quantitation is required to, first, maximize sequencing coverage uniformity and to, second, ensure optimal template-to-bead ratios during templating. Kits used for this kind of quantification measure total DNA concentration in a sample.

Hence, DNA quantification, as described below, was performed twice; first, in order to normalize concentrations of V1–V3 and V3–V4 16S rRNA gene amplicons across all samples, and second, in order to set the final sequencing library to specified concentration required for optimal template preparation and semiconductor chip loading.

Absolute copy numbers of purified and pooled V1–V3 and V3–V4 16S rRNA gene amplicons, each containing Ion Torrent-specific P1- and A-adapter sequences with unique index sequences for each sample, were estimated with the Ion Library TaqMan Quantitation Kit (Thermo Fisher Scientific, Darmstadt, Germany; catalog no. 4468802) on a LightCycler 480 II (Roche Diagnostics, Mannheim, Germany) with white LightCycler 480 multiwell plates 384 according to instructions for the quantitation kit.

The kit uses AmpliTaq DNA Polymerase Ultra Pure. Furthermore, possible carryover of pre-existing trace amounts of DNA by reamplification is eliminated by initially incorporating dUTP instead of dTTP and injecting starting reactions with uracil DNA glycosylase (UDG, [241]) before PCR cycles to make such contaminating DNA unavailable for the PCR at hand. The kit uses hydrolysis probes that are specifically designed for libraries constructed with Ion Torrent P1- and A-adapter sequences. These probes are linked to FAM (6-carboxyfluorescein) as fluorescent reporter dye and a fluorescent quencher ligated to a minor groove binder (MGB) at the other end of the probe. Furthermore, it uses a proprietary quantification standard provided with the kit (called *E. coli* DH10B control library) for absolute copy number determination (i.e. molarity of pooled PCR products, with defined amplicon length for each species).

Samples were prepared in 384-well plates with a semi-automated electronic 8-channel pipette with low-retention tips as described above. The PCR protocol included an initial UDG incubation at 50 °C for 2 min, followed by a Taq polymerase hot-start at 95 °C for 20 s and then, 40 two-step PCR cycles of 1 s at 95 °C and 20 s at 60 °C.

The provided standard was prepared in five sequential 10-fold dilutions starting from a 68 pM stock concentration. Sample templates were diluted 1:10.000 with a manual 8-channel pipette (Rainin Pipet-Lite Multi Pipette L8-200XLS+, with Pipette Tips TR LTS 200µL F 960A/10).

Based on these quantitative analysis results, individual sample amplicon libraries were systematically normalized to a standardized concentration of 1 nM using automated liquid handling protocols executed on a Freedom EVO 100 robotic workstation (Tecan, Crailsheim, Germany) controlled by custom-developed scripts written with Tecan Freedom EVOware software. This normalization ensures equimolar representation of each sample in the final pooled library. Subsequently, equal aliquots (5 µl) from each normalized sample library were combined to generate a composite sequencing library which then represented every sample for both of the targeted hypervariable regions.

The fully pooled library was quantified a second time using the same DNA quantitation kit with 5 µl of undiluted pooled library as template, followed by dilution to a concentration of 17 pM which is considered optimal and therefore required for downstream automated template preparation and semiconductor chip loading using the Ion Chef automated template preparation system (Thermo Fisher Scientific, Darmstadt, Germany) as described below.

16S rDNA sequencing is considered a *de novo* sequencing method because there are no reference genomes. It is therefore recommended, to include Ion S5 Calibration Standard (catalog no. A27988) for Ion 530 Chip workflows (further described below) to the diluted sequencing library in order to increase base-calling accuracy. It is described as a small panel consisting of known sequence content with comprehensive and uniform representation of long homopolymers up to 10-mers. A volume of 4 µl of the Ion S5 Calibration Standard was added to the final library pool and ‘Enable Calibration Standard’ was selected in the Torrent Browser.

##### Template preparation on forward primer beads for library amplification and chip preparation with Ion Chef robot

###### Templating: goal and terms

Most NGS systems, including Ion Torrent technology applied here, require so-called library amplification as a last step before high-throughput sequencing. It ensures that the sequencing technology is provided with many copies of each single library amplicon in a dedicated compartment in order to generate a sufficient signal for readout during sequencing. Hence, this type of amplification denotes monoclonal amplification, as further described below.

Thus, template preparation serves two goals: first, individually linking single amplicons to separate solid supports (here, microbeads) that fit the sequencing technology (here, chip wells), after which these amplicons are referred to as templates and, second, amplifying each bound template in order to create a high number of identical templates bound to a single solid support such that final sequencing is provided with sufficient monoclonal template material for delivering a significant signal from this amplicon clone.

Preparing microbeads for sequencing usually comprises three procedures which tend to be described with ‘seeding’, ‘templating’ and ‘enrichment’ and which can be deployed in various intertwined orders and multi-step processes. *Seeding* describes a procedure of ideally binding a single library amplicon molecule (of around 500 bp) to an attached forward primer (B) on a single microbead with the P1-adapter end. *Templating* describes the process of amplifying these single amplicons in the confined proximity of their bead in order to generate a monoclonal population such that hundreds of thousands of monoclonal single-stranded amplicons will finally be bound to a single bead. Seeding and templating describe processes that often happen within a single reaction step, at least in part. Thus, a first seeding reaction often also contains first templating which may sometimes cause confusion in the use of terms. *Enrichment* denotes the separation of template-positive beads from unbound, i.e. empty beads, for example by capturing them with additional magnetic beads.

###### Ion Chef robot: overview

The complete process of templating is biotechnologically complex and sophisticated across several reaction steps. For the Ion Torrent sequencing variant employed for this trial, all of these steps were performed within a single, closed and fully integrated, automated device, the Ion Torrent Ion Chef robot (Thermo Fisher Scientific, Life Technologies GmbH, Darmstadt, Germany; catalog no. 4484177), controlled by Torrent Suite Software (version 5.18).

Among others, it includes various reagents containers, a pipetting robot, a thermocycler block, two centrifuges for recovery of templated microbeads before enrichment, a magnetic tube rack for enrichment, and a chip centrifuge for chip loading such that this loaded chip can immediately be transferred to a sequencing machine.

The Ion 520 & Ion 530 ExT Kit – Chef (catalog no. A30670, Thermo Fisher Scientific, Life Technologies GmbH, Darmstadt, Germany), or ExT kit in short, provides all materials for templating and chip loading in the Ion Chef, all of them combined in a few closed cartridges. It only requires one additional kit that provides the sequencing chips, themselves, in our case, the Ion 530 Chip Kit (Thermo Fisher Scientific, Life Technologies GmbH, Darmstadt, Germany; catalog no. A27763). Sequencing chips are loaded into the Ion Chef robot together with all other reagents from the ExT kit before the Ion Chef is started. Hence, the ExT kit contains all necessary reagents for chip loading, as well.

The Ion Chef / ExT kit workflow is designed as a walk-away system requiring no operator intervention after initialization. The workflow requires about 20 minutes of hands-on time for reagent preparation. ExT Kit reagents, libraries, and chips are thawed and positioned at designated locations within the Ion Chef robot. Libraries are diluted (50 µl) into specific reagent cartridge positions and consumables are loaded after which a deck scan is performed. After the operator has selected an appropriate protocol on the display interface, the system runs on its own without further intervention.

A single Ion Chef run for templating and chip loading takes about seven hours. Microbeads are templated and enriched outside of the sequencing chip during the first six hours. Isothermal amplification for templating takes about 1 h, followed by 2 h of washing and particle purification. Magnetic enrichment then takes another 2.5 h. After this, non-enriched and enriched template-positive microbeads may be manually taken from the robot for quality control if needed. The Ion Chef, then requires one additional hour for chip loading of which chip preparation is the longest part. The deck is cleaned in a final step (pipetting tips removed, etc.) for about 30 min, after which chips can be removed for sequencing.

We are not aware of any published applications notes, academic reviews or research on biotechnological details, functionality, performance characterization or validation of the Ion Chef instrument or of the ExT kit. The user guide of the kit states that the Ion Chef instrument uses automated isothermal amplification technology with the kit to prepare 400- to 600-base-read templates. This information vaguely corresponds to its predecessor kit, the Ion PGM Template IA 500 Kit whose user guide additionally emphasized non-emulsion PCR templating (of “500” bp templates) in contrast to the original Ion Torrent sequencing publication [242].

Product marketing by the manufacturer implies that automation with the Ion Chef together with the ExT kit may be the primary difference to its predecessor PGM technology, thus inferring the underlying biotechnology may have remained similar. However, while templating and chip loading with the IA 500 kit together with Ion OneTouch ES has only been semi-automated and required substantially more hands-on time, the whole process only required about one third to half of the time that the Ion Chef robot takes. This difference is probably related to the speed and architecture of the robot with a single pipetting head containing a single pipette. Additionally, it might hint towards different templating chemistry, the details of which, however, were not to be found anywhere.

The use of non-emulsion isothermal amplification (IA) for templating is, more or less, the only scientific method information that a user receives for the deployment of the kit. This nearly complete lack of scientifically relevant information on how our prepared sequencing library is being processed inside this robot motivated us to assemble a method and workflow description of most likely used procedures and materials including applied scientific principles. For this, we used published literature, user guides, available patents [243–246] as well as our experience with its predecessor technology. Unfortunately, we still do not know what is truly happening inside the robot for this important pathway before sequencing. Wherever we are certain about protocol details, we mention them or at least describe the detailed settings, parameters and interim outputs of the instruments.

###### Design of forward primer beads

The final goal of templating for Ion Torrent sequencing is to have generated a multitude of identical single-stranded amplicons linked to a single separate mobile solid support (here, a microbead). The ‘classic’ variant of this design is the following sequence: bead / (5‘-) bead-bound forward primer / P1-adapter / sample amplicon / sample index / A-adapter (3’-tail). These ready-made strands then pose as templates for complementary single-end unidirectional sequencing in chip wells initiated from the free 3’-end of the bound template strand, i.e. from the A-adapter.

The supports used for templating in the context of Ion Torrent technology are mobile silica microbeads that are sized to individually fit into a single well of a sequencing chip. Their proprietary name is Ion Sphere Particles (ISPs, Thermo Fisher Scientific, Life Technologies, Darmstadt, Germany). These microbeads most likely are hydrophilic, paramagnetic acrylamide polymer beads [247]. Their diameter has originally been described with 2 µm [242]. More recent sequencing chips (like Ion 540 and Ion 550) are designed with many more and therefore smaller well positions and thus require smaller beads whose diameter may likely be in a range between 0.1 and 0.3 µm [247]. The chip used here, however, belongs to a product line that continues working with bead sizes in a diameter range of around 2.8 to 3.0 µm fitting into chip wells with a diameter of 3.5 µm for the Ion 530 chip. Each ExT kit batch provides six billion ISPs, usually in a concentration of 70 to 80 million ISPs per microliter. This batch is fully used for a single chip run while it is unclear whether all of the six billion ISPs are used for one chip loading or just a fraction as in the IA 500 kit.

Acrylamide monomers, i.e. the chemical building blocks of these microbeads, had been chemically prepared such that they can reveal carboxyl sites after polymerization to beads in order to enable covalent binding to identical oligonucleotides that are used for capturing. There are different variants of these bead-attached forward primers. The original or basic variant was denoted with ‘B’ probably to clarify that these are covalently bound to ‘B’eads at their 5’-ends (5’-CCT ATC CCC TGT GTG CCT TGG CAG TCT CAG-3’, [243]). Modified attached B-primers have been labelled, among others, with AV4, AV5 and AV6 and patents claim higher sequencing yield with a capture primer like AV4 in contrast to the original one. ISPs are therefore also described as ‘forward-primer beads’ and they come with many hundreds of thousands of such identical proprietary forward primers attached to each bead [243].

###### Seeding and initial templating in limiting dilution

For seeding, binding of a single template to one microbead is achieved by applying thermodynamics and thus working at low input concentrations of template per available microbeads. It is a key requirement for seeding to work and a balancing act of creating a sufficiently high number of template-positive beads to achieve high chip loading, while keeping the binding of multiple (i.e. different) templates to a single bead to a minimum. The latter is essential as the sequencing signal from polyclonal beads cannot be used which in turn costs valuable sequencing chip space.

A practical principle behind choosing appropriate concentration ratios for seeding is an old one and called limiting dilution [248]. It describes diluting homogenously suspended particles in solution (e.g. molecules, droplets, cells, etc.) until a probe of a certain volume either captures one or none of these particles. Biologists have used this principle to estimate the number of bacteria in a solution for a long time [249] and it also is a cornerstone for digital PCR [250]. Thus, the concentration of amplicons is diluted as far down as to reach conditions that provide for binary, i.e. single-or-nothing binding behavior such that either a single or no amplicon may bind to a bead. For such a limiting dilution assay, a single-hit Poisson model can be applied in order to theoretically derive a suitable amplicon molecule-to-bead ratio. Empirically, one DNA molecule of about 500 bp per twenty-five beads (1:25) was reported to provide for a template-positive bead fraction of 98 % which was shown to correspond to theoretical estimates of single-hit modelling [251]. Ion Torrent technology uses the same principle and among an array of multiple mentioned ratios, the feasible ratios described in the publication above are contained therein. The final ratio possibly is in the range of 1:25 to 1:50 and usually needs to be fine-tuned for each method and each lab.

We found a concentration of 17 pM library stock (i.e. 0.51 × 10^9 total DNA template molecules per 50 µl) to be best for our use case after testing a dilution series of our validation library. Thus, a sample library concentration of 17 pM usually resulted in high chip-loading rates of more than 90 % (i.e. more than 90 % of chip wells were loaded with a single template-positive microbead), polyclonality rates of less than 30% and more than 30 % of usable reads.

For limiting dilution to be applied successfully, it is not necessary to have any separate confinements, e.g. by plate wells or by a dispersed phase in an emulsion. Thus, the mere seeding process is well possible in a single solution, i.e. within a continuous aqueous phase. The patents mentioned above describe multiple examples for doing so and it is probable that neither the IA 500 nor the ExT kit apply the principle of confined microspaces for templating reactions.

###### Biotechnological challenges during templating

Seeding, templating and enrichment, however, require some more aspects to consider beyond limiting dilution and the following are just some of those challenges: First, the individual amplification processes for single templates during templating create free amplicons in solution which would diffuse to other beads if not contained. Second, prepared Ion Torrent library templates are double-stranded amplicons with blunt ends at both sides. They contain P1- and A-adapters and their complementary counterparts (A’ and P1’) at their ends. In this situation, binding to beads with already bound forward primers (B) is not possible unless strands are dissociated and modified during seeding and template amplification. Third, when theoretically illustrating a templating reaction, one might assume that one of the many forward primers bound to a microbead might be priming a dissociated single complementary amplicon strand contained in the aqueous droplet together with the bead. This however does not seem to happen sufficiently because researchers like Dressman et al. [252] had already found that priming by oligonucleotides coupled to beads was very inefficient compared to priming with the same oligonucleotides in free solution. Fourth, the same researchers realized that adding free primers to the reaction mix in order to initiate templating might let these free primers compete with bead-bound primers leading to alternate design considerations. Fifth, Vogelstein and Kinzler [253] had observed that some of pre-confined single templates in free solution were not amplified until several cycles of PCR. Sixth, while the template library is ready-made with P1- and A-adapters, it is not yet prepared for enabling the enrichment process for which the adapter sequences require further modifications. Beyond these general challenges in regard to templating, a seventh also needs to be considered: it is a characteristic of 16S rRNA PCR amplicons themselves, that they can form complex secondary structures as is the nature of original full-length 16S rRNA. These secondary structures may interfere with primer annealing and polymerase processivity during templating [254,255]. This phenomenon particularly affects GC-rich regions [256] or stem-loop formations that might compete with adapter-primer hybridization.

These are just some of the reasons why templating requires sophisticated fine-tuning for setting up separate reactions, proper confinements and optimized reaction mix concentrations.

There are different ways to address the first aspect of free amplicon diffusion to other beads during template amplification. Constructing a multitude of physicochemical microreactor compartments for seeding and templating is one way to do so, either by individually locking free single library amplicons (double-stranded) from a sample as well as (hydrophilic) microbeads into dispersed phase aqueous droplets, formed from an emulsion in oil as external phase [257], or by locking seeded microbeads in chip wells for templating before sequencing. Other ways to reduce amplicon diffusion, possibly applied here, is, first, to keep reactions in a limiting dilution, second, to shorten and split reaction steps into a multi-step templating process and, third, to add diffusion-limiting compounds like sieving agents. An advantage of these latter reaction optimizations is that amplification products do not need to be freed from their lock in oil.

All other aspects mentioned above may be addressed by involving modified primers, optimizing concentrations of components as well as number and duration of reaction processes, for example by splitting the template amplification process into two or three individual reaction steps.

While the manual of the IA 500 kit describes a workflow with a single bulk isothermal amplification of 25 min for seeding and templating within the same reaction process, this may have been split up for the ExT kit in the Ion Chef robot as indicated in a patent example [243] that describes a 4 min IA, followed by a 30 min IA, both at 40 °C.

A multi-step reaction like this provides for creating different reaction conditions for each step that can be optimized individually and fine-tuned to each other.

For example, a first and short seeding reaction with free library amplicons (and primers) in limiting dilution would enable seeding with little risk of diffusion. Thus, concentration of initial free template and primers can be kept low and initial template amplification products can also be kept low in order for the majority of these templates to be hybridized to the bead instead of diffusing into neighboring spaces with other microbeads.

Free library amplicons would no longer be added in a second and longer templating reaction which would only amplify already bound templates on microbeads in a bulk amplification. Here, diffusion rate of free template amplification products could be slowed by addition of diffusion-limiting agents. Splitting and optimizing reaction steps in this way would help to reduce polyclonality on beads.

Diffusion-limiting agents may include sieving agents like cellulose polymers with specified pore sizes as used in electrophoresis and which also increase viscosity, as well as drag compounds which directly attach to templates to reduce their mobility. Drag compounds often require template modifications for binding, so-called drag-tags. One example for a drag compound in our context might be Neutravidin that binds to biotinylated A-primers which serve as drag tags. When these primers become part of a template during a first amplification step, Neutravidin may cause increased hydrodynamic drag on a template which is not yet bound to a bead.

A consequence of these optimization attempts is that a first reaction step may lead to no more than 10% of bead-bound primers loaded with template [245] and probably much less. These first bound templates, in turn, further help to reduce template diffusion during the second mass template amplification step. Consequently, a two-step template amplification reduces polyclonality on beads, increases templating efficiency and thus generates more usable sequencing reads. The final number of bound templates is supposed to be in a range of 50 to 500 hundred thousand template molecules per bead [243].

###### Primer extension during templating for building an anchored target strand

Primer **Ex**tension during **T**emplating has possibly provided the name for the “ExT” kit used here, as multiple primer extension steps are performed with this kit for templating. There are different approaches to do so, in order to connect a P1/A-strand (i.e. target strand) of a double-stranded sequencing library amplicon with blunt A/A’- and P1/P1’-ends to a bead with covalently bound forward primer B. The design concept is mostly independent of the amplification method used for construction.

Whatever building process and materials are used in detail, a general construction principle stays constant for seeding and templating Ion Torrent ISPs: modifying and building a complementary amplicon strand that can hybridize with the forward primer (B) anchored on a bead. This strand then serves two purposes: for one, it provides the template for building the target strand starting from the anchor forward primer on the bead. Second, it is used for capturing during enrichment before it is separated (i.e. denatured) from the target strand and washed away.

For this to work, the complementary strand, i.e. A’-x(n)-P1’ needs to be modified at both ends: the free 5’-end (A’) receives a biotin-extension (either directly or indirectly in a consecutive step) and the 3’-tail (ending in P1’) is extended by a complementary sequence (B’) for hybridization with the anchor primer (B) on the bead. Simplified, this makes for three extension reactions: at A’, at P1’ and for building the anchored target strand.

In order to do so, Life Technologies has described four different ways [243,245] of which all can be distinguished by the design and functionality of primers that are used to anneal to P1’-ends. One way is probably the original and first method as applied for Ion Torrent PGM instruments involving a blocked reverse fusion primer with a full B- and P1-sequence, in which a polymerase block is created with a phosphate group at the 3’-end of the P1 sequence of the primer. Another way requires that amplicon P1-ends have additionally been extended by B/B’-sequences during library preparation. A third way uses an unblocked reverse fusion primer with a truncated P1- and full B-sequence (trP1-B) for annealing to the outer (3’) end of P1’ and a fourth uses a similar truncated P1 fusion primer for annealing to the inner (5’) end of P1’ causing mismatch B/P1’-ends. It is unclear which extension method is used here while the first might still be likely.

Modifications at the A-adapter ends are simpler and can be distinguished by the planned capture methods for enrichment, i.e. direct versus indirect capture. This means either direct conjugation of a biotin to an A-primer such that the biotin will be directly linked to the complementary template strand, or coupling of a short oligonucleotide ‘handle’ to an A-primer via a polymerase stop-sequence such that a short biotinylated oligonucleotide can later hybridize to this handle on the complementary template strand for indirect capture during enrichment. It is unclear which capture method is used here, while the ‘classic’ method applied direct capture.

###### Enrichment of template-positive microbeads

A consequence of applying limiting dilution for seeding is a large fraction of template-negative beads of about 70–90 %, i.e. beads to which no amplicon has bound, following the single-or-no-hit principle. Template-negative beads would waste valuable sequencing space, i.e. chip wells, and therefore need to be excluded. Enrichment describes this process from the perspective of template-positive beads.

Similar to limiting dilution, the scientific principle for the most frequent enrichment method applied here is an old one: avidin-biotin technology [258–260] combined with magnetic beads [261,262].

The bond of avidin, as well as of its derivative neutravidin and its relative streptavidin, to biotin is considered the strongest known non-covalent bond [263]. Correspondingly, dissociation of this bond may be chemically challenging in practice. For enrichment here, though, this dissociation is not required as will be described further below.

Magnetic beads are also known by their brand name Dynabeads (as originally developed by John Ugelstad in his academic startup Dynal Biotech in Oslo, Norway, now part of Life Technologies at Thermo Fisher Scientific) and continue to be a gold standard for capturing, isolating, and handling biotinylated molecules. There are many different variants of these beads. A variant that is deployed in the IA 500 kit and that might also be used here are Dynabeads MyOne Streptavidin C1 (Thermo Fisher Scientific, Life Technologies, Darmstadt, Germany; catalog no. 65001). These are described as uniform, superparamagnetic beads of 1.0 μm in diameter with a streptavidin monolayer covalently coupled to the hydrophilic bead surface with overall increased binding capacity and lower sedimentation rate which make them preferential for automated high-throughput applications.

The capturing of template-positive ISPs is designed such that the complementary template strand is the one linked to biotin. This strand is not directly bound to a microbead and only linked to it indirectly via its hybridization to the template strand. Therefore, it can be eluted away after melting it off from the template strand which will include biotin-streptavidin-bound Dynabeads after enrichment. Hence, the strong biotin-streptavidin-bonds does not need to be broken.

As described above, constructing an appropriate complementary amplicon strand during template extension is key for the whole process to work well including enrichment. Furthermore, multiple enrichment steps are possible. If, for example, the templating IA reaction is split up into two as described above, then there might be one enrichment step after each IA step [243].

###### Quality control of template-positive microbeads

Quality control of ISPs during seeding and templating is important for at least two reasons. First, the number of template-positive ISPs needs to be known in order to generate an appropriate concentration for limiting dilution of template-positive ISPs for subsequent reaction steps. Second, it is helpful to determine and filter the ‘size’ of ISPs. Here, size denotes the amount of monoclonal template copies per bead. Ensuring that selected ISP have the same ‘size’ prevents sequencing bias towards beads with higher template loads, later.

Therefore, the goal of this quality control step before mass templating is to select seeded ISPs with a similar number of template copies on each ISP in a defined bead concentration for the mass templating step.

Similar to the Ion OneTouch ES instrument, the Ion Chef robot allows interrupting in order to retrieve unenriched or enriched template-positive microbeads. This enables quality control of templating and enrichment processes and is often performed with a flow cytometer [243].

###### Isothermal amplification: overview

The Ext kit user guide describes deployment of isothermal amplification for templating. The first isothermal amplification (IA) method [264] was published two decades before the first polymerase chain reaction [265] but the principle only got traction when PCR had already been established. Hence, the term ‘isothermal’ was introduced after PCR [266] to emphasize a contrast to repetitive temperature changes (i.e. thermal cycling) used during PCR for denaturing (i.e. melting) double-stranded DNA into single strands by fully opening up the hydrogen bonds between paired bases. Since then, many different types of isothermal amplification have been published and more than half a dozen of these methods are in frequent use for various applications [267,268].

Opening up double-stranded DNA far enough to enable primer annealing to desired extension sites is a key condition for any amplification method to work and full DNA melting at high temperatures is just one of multiple approaches to do so, with other options being the use of sodium hydrochloride, low ion content or displacement with an enzyme as described below. Hence, the thermodynamical principle of so-called DNA-breathing [269,270] is probably essential for all physiological and biotechnological modifications of DNA. It describes ongoing local DNA fluctuations (i.e. breathing sites) in the duplex structure of helical dsDNA around its (fully bound) ground state conformation at room temperature, preferentially at certain nucleotide sequences. These local melting transitions may be conceptualized as one-dimensional first order phase transitions. Thus, DNA breathing is quite likely an important supporting phenomenon for site access to enzymes in most isothermal amplification methods.

Three different amplification methods have been described for Ion Torrent templating over the years. Emulsion PCR [252,257] had originally been reported for their first chip series [242]. However, patents indicate [243,244] that emulsion PCR has mostly been displaced by isothermal amplification [264] which does not necessarily need to be performed in emulsion, at least during a first step. The first of two applied isothermal amplification methods was originally named ‘WildFire in-situ solid-phase isothermal amplification’ with a so-called template walking mechanism [244,271]. This method uses indirect strand invasion by prior strand-nicking as originally described for strand displacement amplification (SDA) [272] even though we could not find a reference to SDA for template walking. However, the isothermal amplification method that has probably been in use for Ion Torrent templating for more than a decade is recombinase-polymerase amplification (RPA) [273,274] which applies direct strand invasion with a recombinase-primer filament.

###### Recombinase-polymerase amplification (RPA) for templating

The method name already implies that two enzymes are central to RPA: a recombinase and a polymerase. The assumed molecularbiological mechanism for the physiological function of a recombinase [275] in DNA recombination and repair is that it binds cooperatively to single-stranded DNA to form a nucleoprotein filament. Assembly of this filament complex requires ATP such that a recombinase also is a DNA-dependent ATPase. This recombinase-ssDNA filament then scans along double-stranded DNA for sequence homology to bound ssDNA. In RPA, primers are used as this ssDNA to form a recombinase-primer filament.

The recombinase that has originally been used for RPA appears to remain the enzyme of choice: uvsX protein from T4-like bacteriophages [276]. This recombinase requires an accessory protein, T4 uvsY, in order to prevent rehybridization of strands dissociated by the recombinase. Thus, RPA is started after primer binding to T4 uvsX in the presence of ATP and uvsY to form a recombinase-primer complex which delivers primers directly to their annealing sites within the double-stranded template at stable reaction temperatures of around body heat (37° C) and even of room temperatures. After this recombinase-primer complex has disassembled following primer annealing and ATP-hydrolysis, the recombinase proteins can be substituted by a single-stranded DNA binding protein gp32 (for T4 Gene 32 Protein), sometimes also called SSB. This protein is required for the ‘d’isplaced strand (D-loop) of dsDNA to stay open during elongation. Hence, it stabilizes this transiently formed D-loop of ssDNA during elongation.

The recombinase disassembly also opens up access to the extension strand for a polymerase. Thus, elongation from the primer annealing site is then performed by a mesophilic DNA polymerase with high strand-displacement activity. One of several possible polymerases can be deployed. A patent states [245] that the originally used large fragment of *Bsu* (*Bacillus subtilis*) DNA polymerase I which thus lacks its 5’ to 3’ exonuclease domain (i.e. exo^-^) as well as T7 DNA polymerase with reduced exonuclease activity and Sau polymerase (exo^-^) would be particularly well suited for an RPA reaction. Moreover, two further polymerases are potential candidates for RPA: *Bst* (*Bacillus stearothermophilus*) DNA polymerase (exo^-^) as used for SDA at higher temperatures [277] as well as for template walking [271], and DNA polymerase from *Bacillus subtilis* phage phi29 [278]. RPA reaction mixes usually contain two polymerases with T7 DNA polymerase as a fixed element. For the ExT kit, it appears likely that the other is *Bst* DNA polymerase (exo^-^) or possibly *Bsu* DNA polymerase (exo^-^).

The design of RPA appears simpler at first, in contrast to methods like SDA or template walking that require template nicking to provide access for a strand displacement polymerase. In RPA, however, a ‘dynamic reaction environment’ [273] is described to be key for it to work well, but also to be challenging. Among others, the management of this dynamic reaction environment requires continuous shifts in equilibria of reactants for recombinase loading and unloading with primers and supplements like a crowding agent [279,280] (here, a high-molecular polyethyleneglycol) together with energy-providing ATP. This may explain why two Life Technology patents include the use of proprietary RPA reaction mixes provided by TwistDx, a company founded by the scientists who published RPA. Furthermore, primers need to have a minimum length of 30 bases for a recombinase like uvsX to work [273].

###### Chip selection criteria and Ion Chef settings

Number of possible sequencing reads per sample is the decisive factor for chip size selection. Selection of an appropriate Ion chip was based on number of sample libraries to be sequenced, together with the average sequencing depth. An Ion 530 chip contains approximately 37.8 million addressable wells and provides for about 15 million sequencing reads per run with an output of 6 giga bases at an average length of 500 bp. It is designed for intermediate scale sequencing applications.

For example, aiming for a minimum of 20000 reads per sample and variable region (i.e. more than 40000 total reads per sample), this chip size will allow for analysis of around 375 samples per run (based on total sequencing capacity of 15 million reads). This read depth ensures sufficient coverage for accurate taxonomic classification and detection of low-abundance microbial taxa in complex microbiome samples.

Sample libraries for a single sequencing run were assembled such that, for example, all fresh frozen samples for both hypervariable target regions were included. Similarly, all samples buffered with OMNIgene.GUT for both hypervariable target regions were pooled in a single sequencing run, etc. This sequencing design made for a range of 320 to 370 samples per sequencing run.

###### A possible reaction workflow for templating in the Ion Chef robot

Our sequencing library is what is often called a ‘complex’ library, in other words, it contains a multitude of different amplicons from different samples and different 16S rDNA hypervariable regions. This library has been prepared with long fusion primers to generate amplicons with standard Ion Torrent A- and P1 library adapters, i.e. with P1- and A-located on the same strand (and A’ and P1’ on the complementary strand). These are first cornerstones for what might happen next.

Synthesizing published information and scientific principles described above together with our experience with predecessor technology like the Ion PGM IA 500 kit, as well as examples from mentioned patents and combining these with the basic configuration of our prepared sequencing library, it still remains unclear what is happening with our samples in the Ion Chef robot during the first six hours of seeding and templating, in detail.

We can be certain about the following aspects: 1.) the primary templating amplification method is isothermal amplification with RPA which 2.) is probably not performed in any emulsion at any point, and 3.) microbeads are fully templated and enriched outside of the chip during the first six hours and 4.) the chip is not loaded with magnetic distribution of template-positive beads but with simple centripetal forces in a chip centrifuge.

Everything beyond that is speculation and exclusively leans on examples in published patents, in particular example no. 4 in [243] which describes a non-emulsion two-step IA reaction (short and long) for seeding and templating of microbeads for a complex library.

The use of RPA for templating, however, does not exclude that initial seeding is performed with conventional PCR before a two-step IA as indicated by example no. 6 in the same patent. Such multiple reaction processes might explain the considerably longer runtime of six hours in the Ion Chef robot in comparison to use of the IA kit in about one third of this time, before.

Hence, initial seeding might be performed either with two conventional but short PCR reactions within a single tube. In the PCR variant, library amplicons with ‘classic’ P1- and A-adaptors are first modified in a two-cycle PCR (e.g. extension for 2 min at 58 °C each) with a pair of soluble forward and reverse primers: a biotinylated truncated A-primer (bio-trA, i.e. direct capture) and an unblocked fusion primer for P1 (trP1-B). Microbeads are not yet present in this mix, but it is already prepared in a limiting dilution like 100 pM. After this adapter-modification step, six billion ISPs are added (in limiting dilution) to the same reaction tube followed by a second PCR step with a single cycle for seeding in limiting dilution (e.g. extension for 5 min at 56 °C) and a reaction stop with EDTA.

Alternatively (or supplementary), a first IA reaction step might be used. Here, the amplicons with ‘classic’ P1-and A-adaptors are mixed with soluble primers for adapter modification together with ISPs in a single step in annealing buffer in a limiting dilution; in this case without any amplification enzymes at first. Amplicons are denatured for 2 min at 98 °C and incubated with primers and ISP for 2 min at 37 °C followed by addition of an RPA reaction mix that only shortly incubates for 4 min at 40 °C.

An annealing buffer (sometimes AB in short) mostly forms the basis for preparing primers in amplification solutions or for simple washing steps in between. Its recipe ensures optimal hybridization of primers (i.e. annealing). A typical composition for an annealing buffer is 10 mM Tris-HCl (to maintain a stable pH during reaction), 50 mM NaCl (to promote hybridization with sufficient ions) and 1 mM EDTA (to prevent degradation by nucleases).

A first enrichment with streptavidin-coated dynabeads (e.g. MyOne Streptavidin C1) then follows this short seeding with or IA or PCR. Washed dynabeads are added to the stopped PCR reaction mix with seeded ISPs and biotinylated templates for incubation up to half an hour at room temperature. After pulling out the dynabead-bound seeded ISPs with a magnet, seeded ISPs are melted off and eluted from dynabeads with a melt solution (e.g. with 125 mM NaOH and 0.1 % Tween-20 detergent in water – as used for IA 500) leaving ISPs, each ideally bound to relatively few monoclonal single-stranded templates. Such an enrichment step could be repeated later, for example, after a first and second isothermal enrichment step.

Whether with or without this PCR for initial seeding followed by first enrichment or not, it is likely that at least two IA reaction steps are performed outside of the chip for templating the complex library in limiting dilution including diffusion-limiting agents with soluble trA and trP1-B primers, with trA either directly linked to biotin or to a handle for indirect capture. The first IA step might be performed for initial seeding alternatively to PCR as described above or perhaps even follow it in a two-step templating process.

After first seeding and enrichment, a second and longer IA reaction might then provide for bulk templating of seeded microbeads with similar reaction parameters but with a longer reaction time, e.g. 30 min at 40 °C. This, then, would go through a second enrichment step before an opportunity for quality control and chip loading.

Such a second isothermal amplification step does not require any free library amplicons in solution and only amplifies already bound templates on microbeads. This process splitting substantially helps to reduce polyclonality on beads.

In the past, the RPA reaction mix has usually been provided as a pellet that needs to be rehydrated for immediate use, probably because of the complex composition of reactants. This has been the case for the original TwistAmp Basic kit as well as for the Ion PGM Template IA 500 kit which both require manual preparation of reaction steps in contrast to complete automation with the Ext kit. The IA 500 kit user guide also remarked that rehydrated IA pellet solution is viscous, indicating the use of diffusion-limiting agents.

The patent primarily used here for reference, also describes the detailed composition of the TwistAmp Basic kit [243] and, at least, of the components of an Ion IA pellet. An Ion RPA solution is described to contain: uvsX recombinase with uvsY, gp32, Bsu DNA polymerase and T7 polymerase besides ATP, thioredoxin, phosphocreatine, creatine kinase and deoxynucleotide triphosphates (dNTPs). In contrast, many more details are provided for a TwistAmp pellet, with some relevant differences being the use of Sau (instead of Bsu, together with T7) polymerase and Trehalose and PEG-35, in a Tris-buffer at pH = 8.3.

Seeded, templated and enriched microbeads are then mixed with Test Fragment (TF) ISPs provided as positive control microbeads with the ExT kit. They are added in an amount that makes for 1 % of total ISP volume.

The Ion Chef robot also automates chip preparation for sequencing for which the ExT kit provides all materials, as well. The mixture of enriched template-ISPs and test fragment ISPs is injected into the chip (here, an Ion 530). This loading step is another essential step; its goal is to ideally fill all of the nearly 38 million addressable chip wells with a template-positive microbead each.

Even distribution of these microbeads across chip wells can be achieved by either with centripetal forces by centrifuging or by magnetic loading in order to seat single beads into their individual chip wells. The Ion Chef has a chip centrifuge built in. Therefore, it is likely that magnetic loading is not deployed; it would require, among others, a magnet that sweeps back and forth below the chip repeatedly to load ISPs into wells. After distribution of ISPs within the chip, beads that have not found a well need to be removed. This is usually done with a foam (e.g. annealing buffer with 0.2 % Triton-X 100) that is injected into the chip. Thereafter, the chip is flushed, dried and filled with annealing buffer.

Chip wells with loaded ISPs then need to be filled prepared for sequencing. In a first step after chip loading, templated ISPs in chip wells are primed by annealing sequencing primers complementary to the A-adapters region. After the chip is dried from annealing buffer, a primer mixture is injected into the chip and thermocycled for annealing (e.g. for 2 min at 50 °C and for 5 min at 20 °C), again, followed by refilling the chip with annealing buffer.

A sequencing enzyme mixture is then injected to replace the annealing buffer and left for incubation (e.g. for 5 min at room temperature) followed by drying and refilling with annealing buffer. The sequencing polymerase originally published has been a Bst polymerase. At that point, the chip is ready to be place into the sequencing system.

At this point, an Ion 530 chip is ideally prepared with single, massively templated ISPs in each of its wells, with sequencing primer annealed on templates and sequencing polymerase present in all wells while the chip is filled with annealing buffer. Each ISP should have several hundreds of thousands of monoclonal templates bound to it and the sequencing chip used here (Ion 530) provides for a total number of 37849615 addressable wells, i.e. fluid-accessible sensors. In this state, the chip is ready for massively parallel sequencing of the ISP-templated amplicon library.

##### Sequencing of library templated ISPs based with Ion GeneStudio S5 Plus system

###### Principle of Ion Torrent sequencing

The Ion S5 sequencing system performs sequencing by synthesis. It detects the release of protons in case a nucleotide is incorporated into a growing DNA strand. Each of the four natural DNA nucleotides (dNTPs) is serially injected into the chip (i.e. ‘flow’) in a flow order that is specifically designed for this sequencing technology to reduce sequencing errors. Each dNTP flow is followed by a wash step. In case an injected dNTP is complementary to the next base on the bead-bound template, it is incorporated by the sequencing polymerase during primer extension on templates. Proton ions are released as a by-product of this dNTP incorporation.

The basic principle of Ion Torrent sequencing technology has been published [242] and it does not appear to have changed considerably since then beyond following Moore’s law [281] such that more addressable chip wells have become available in later chip generations. It applies ion-sensitive field-effect transistor (ISFET) technology [282] with a tantalum oxide layer at the base of each well to provide for ion, and here, proton sensitivity [283]. This layer can detect the release of proton ions, i.e. change in pH, in each well. The measured voltage in each well during each flow is correlated with the number of the same nucleotide incorporated during that flow. The manufacturer therefore advertises the semiconductor sequencing technology as ‘the world’s smallest solid-state pH meter’ [284].

An ISFET chip belongs to a larger group of chemFET semiconductors. In these chemical field-effect transistors, a specialized field effect transistor acts as a chemical sensor. In turn, chemFETs are structural analogs of a MOSFET (metal–oxide–semiconductor field-effect transistor), where the charge on the gate electrode is applied by a chemical process. Thus, in an ISFET, a change in proton concentration within a chip well changes the current through the transistor, accordingly.

###### Ion GeneStudio S5 Plus: configuration

Template-loaded Ion 530 chips were taken from the Ion Chef robot and placed into an Ion Torrent Ion GeneStudio S5 Plus System (Thermo Fisher Scientific, Life Technologies, Darmstadt, Germany; catalog no. A38195) for sequencing. The time between end of Ion Chef runs and start of sequencing runs was less than 5 min. Total sequencing time of one Ion 530 chip in the Ion S5 was four hours.

The (Ion Chef and) Ion S5 sequencer was controlled by Ion Torrent Suite Software (version 5.18). Both, ‘Template Kit’ was set to ‘Ion 520 & Ion 530 ExT Kit-Chef’and ‘Sequencing Kit’ was set to ‘Ion S5 ExT Sequencing Kit’, while ‘Library Kit Type’ was set to ‘Ion Fragment Library Kit’ without further meaning as there was no library preparation on the Ion Chef instrument. Among the two default flow numbers, sequencing runs were configured for 1350 nucleotide flows corresponding to 600 base-pair reads.

The sequencing protocol uses ‘Ion samba’ flow order as default. Flow order is the order in which single dNTPs are injected into a chip. This default flow order consists of a 32-base sequence which is continuously repeated (here, over a total of 1350 flows). It is designed to resist phase errors by providing opportunities for out-of-phase molecules to catch up, and to sample all dimer sequences efficiently. Resisting phase errors particularly improves sequencing accuracy for longer reads. Flow order for resequencing needs to be different from flow order for sequencing. Hence, ‘Ion samba.contradanza’ is recommended for resequencing.

We used ‘Ion samba.contradanza’ flow order together with common other settings like ‘TCAG’ for forward library key, ‘Ion P1B for forward 3’-adapter sequence, and ‘ATCG’ for test fragment key. In our case, contradanza flow order showed slightly better homopolymer accuracy and performance with certain GC-content.

Calibration standard functionality was enabled in the software configuration. Calibration was performed by spiking Ion S5 calibration standard (catalog no. A27988) into the final library pool before templating such that the library pool reached its target dilution together with added standard solution of 4 µl. Adding this standard increases base-calling accuracy in the absence of reference genomes. Well signal normalization was applied to reduce spatial biases across the chip surface.

Base-calling algorithms were implemented with quality control parameters including disabled quality trimming (--trim-qual-cutoff 100), minimum index sequence representation of 10 reads per sample, phasing residual filtering threshold of 2.0, and adaptive quality filtering with slope coefficient 0.080 and offset 1.0. Wells normalization was enabled to compensate for spatial signal variation across the semiconductor chip surface. Primary data processing employed the FileExporter plugin to consolidate sequencing reads including index sequences into a unified FASTQ format while preserving sample-specific identifiers within read headers, thereby facilitating subsequent bioinformatics workflows for taxonomic classification and phylogenetic analysis. Sequencing quality metrics were monitored with Ion Torrent server dashboard. Key parameters included read length distribution, chip loading percentage, usable reads per million wells, and polyclonality rate.

For the three sequencing runs as described below, the quality metrics were: run-1: 51 % usable reads, 29 % polyclonals, 84 % loading. 31905890 library ISPs, 496 bp (median); run-2: 50 % usable reads, 27 % polyclonals, 83 % loading, 31073692 library ISPs, 516 bp (median); run-3: 50 % usable reads, 29 % polyclonals, 86 % loading, 31905890 library ISPs, 497 bp (median).

###### Design of sequencing runs for this trial

After nucleic acids from all trial samples had been extracted over four consecutive days by a single operator, they were stored in 1.5 ml screw-cap tubes at 2–8 °C until library construction. Samples were processed within one week for preamplification of 16S rRNA gene libraries, including quantification of total and spike-in bacterial 16S rDNA copy numbers and subsequent concentration normalization. Following library preparation, nucleic acid extracts from stool samples were frozen at −20 °C for long-term storage. Purified PCR amplicons after preamplification of V1V3 and V3V4 16S variable regions were stored at −20 °C until further library construction including qPCR library quantification, normalization, and pooling.

Sequencing runs were performed according to the following scheme: one run contained a total of 348 samples comprising all fresh frozen specimens (N=174) amplified for both hypervariable regions (V1V3 and V3V4). Another run consisted of 366 samples including all OMNIgene-collected specimens (N=182) for the same two regions. A third run encompassed 324 samples from all healthy participants (N = 25+2) for all three storage methods (fresh frozen, OMNIgene.GUT, and MaGix PBI), again, for both hypervariable regions. For this run, material from both stored stool aliquots (see section ‘Sample storage’) were taken while one aliquot was used for the others.

#### Bioinformatical pre-processing after sequencing

##### Computing infrastructure for microbiome sequencing and analysis

The microbiome analysis division in our microbiology department has established a dedicated computational infrastructure to support 16S rRNA gene sequencing data management and data analysis.

Computing hardware comprised an HP Z8 G4 workstation with dual Intel Xeon Gold processors (36 cores total), 396 GB RAM, four 4-terabyte drives in RAID5 configuration, and dual 1-terabyte SSDs for high-performance operations, operating under Linux Mint 21. Machine control was achieved through network integration between Ion Chef, Ion S5 sequencing platforms, and Torrent Suite Software. FASTQ files were manually downloaded from the sequencing system and deposited into designated processing directories alongside raw data from qPCR measurements, library preparation quantification and normalization protocols, and comprehensive sample metadata. Quality control monitoring utilized automated Torrent Suite metrics including read distributions, base-calling accuracy, and sample-specific performance indicators. Data organization followed hierarchical file systems with structured directories organized by project and sequencing run identifiers. Version control maintained systematic metadata documentation and standardized naming conventions ensuring complete data provenance. Redundancy mechanisms included automated daily incremental backups and monthly comprehensive archival to long-term storage systems, providing data integrity and disaster recovery capabilities while ensuring compliance with research data retention requirements.

##### Preprocessing of 16S amplicon sequencing reads

Signal processing and base-calling was performed using Torrent Suite Software Version 5.18 (as part of Ion GeneStudio S5 Plus system, Thermo Fisher Scientific, Darmstadt, Germany) yielding raw sequencing data in FastQ format without quality trimming. Every bioinformatical processing step from there on may underlie future optimization according to state-of-the-art knowledge at the time of analysis. The following steps reflect the current state. Raw sequences will be filtered for quality with Trimmomatic 0.39 [285] by removing reads with an average Phred quality score [286] of below 15 within a sliding window of 10 bases and removing sequences shorter than 150 bases [-phred33 SLIDINGWINDOW:30:17 MINLEN:150]. In our experience, these parameters yield a read loss of about 30 %. Sequences will then be sorted according to their sample-related index sequences as part of the adapters (demultiplexing) without allowing for any errors with cutadapt 4.1 [287] including removal of 16S-specific primer sequences and sequencing adapters, including A- and P1, as well as sample index sequences.

##### General analysis pipeline for preprocessed sequencing reads

Constructing exact amplicon sequence variants (ASVs, [288,289]), followed by taxonomic classification will be performed in a workflow based on the vsearch package [290] (currently, version 2.30) which is an open-source-variant of usearch [291]. In context of usearch- and vsearch-based algorithms, ASVs are commonly called zero-radius operational taxonomic units (zOTUs, [292]) – conceptually, they are the same.

Basic quality trimming of sequencing reads with prior error filtering according to their Phred quality scores has already been performed with Trimmomatic as described above. This kind of filtering is taking quality scores from single sequences. In contrast, vsearch/usearch offers quality filtering according to posterior error probabilities. Here, it is possible to filter according to expected number of errors that would be observed in the whole collection of sequences and in which the error rate at each position is also taken from the Phred score. Thus, sequencing reads will be removed with more than five expected errors (vsearch --fastq_filter --fastq_maxee 5.0)

Quality-filtered reads will then be clustered into zOTUs by applying an alpha-value of 2 and a minimum size of 5 reads (vsearch --cluster-size 5). Chimeric sequences will be removed using the uchime3_denovo algorithm– this uses uchime2_denovo [293], but with a different default for minimum abundance skew of 16.0 instead of 2.0. Filtered reads with 98 percent pairwise identities will be mapped back to non-chimeric zOTUs applying the usearch_global algorithm [291]. Taxonomy will be assigned in the R language [294], current version 4.5.1, using the IDTAXA classifier of DECIPHER 2.30 [295] together with the All-Species Living Tree database version 10.2024 [296]. Here, a 98 percent bootstrap-cutoff will be used to descend the tree, taxonomy can be reported at each taxonomic level with a confidence value threshold of 40.

### Axis IV: Inflammatory proteome

#### Proximity extension assay technology with Olink Explore 384 ‘Inflammation’ – overview

Proximity extension assay (PEA) technology provided by Olink (Uppsala, Sweden) promised to yield high readout specificity and sensitivity to at below pg/ml-protein concentrations and low dropout rates which appeared more suitable for the delicate blood analytes of this trial than conventional multiplex immunoassays as the material was stored at –80 °C between three and seven years as is common in clinical trials like ours.

Outlining the principles of this method: when a pair of two so-called ‘proximity probes’ – two matched antibodies labelled with a unique set of DNA oligonucleotides – simultaneously bind to a target protein (first step), their bound DNA oligonucleotides have the chance to hybridize at their unique annealing sites in case they are in close proximity (second step). Then, a DNA polymerase extends these hybridizing oligonucleotides (third step) creating a double-stranded DNA template unique to its specific target protein. These unique DNA templates are then pre-amplified and purified for subsequent detection and quantification by Next-Generation Sequencing (NGS, fourth step). In contrast to usual NGS applications, DNA sequences for Olink analysis are known in advance. Hence, in this context, NGS is used to count PEA amplicons. Matched NGS counts then represent original protein concentration for each sample on a relative scale.

#### Sample preparation for Olink Explore

Olink analysis for this trial was performed at the Metabolomics and Proteomics Core Facility at Helmholtz Zentrum München, Germany, the first so-called platinum-level core facility for Olink technology in Germany [297].

In total, 214 serum samples were analyzed which had been taken in the mornings between 7:00 and 07:30 hours before breakfast: one hundred and eighty-nine longitudinal serum samples from 68 patients over three time-points (study days 1, 11 and 60) and 25 samples from 25 healthy subjects. Fasting venous blood from patients and healthy participants had been collected into serum containers (Serum CAT S-Monovettes, Sarstedt, Nümbrecht, Germany) and were immediately centrifuged at 1516 g for 10 min at precooled 8 °C before long-term storage at –80 °C in micro tube aliquots until analysis.

The number of samples required three 96-well plates for sample preparation of which each allows for 88 analyte positions per plate and therefore for 264 measurements in total. The remaining 48 positions were therefore filled in double or triple with selected participants for redundancy and test-retest quality control.

After first thawing and before dispensing to PCR plates, vortexing and centrifuging of this material was repeated at 2000 g for 10 min at precooled 8 °C.

Olink recommends the use of their R package ‘OlinkAnalyze’ for randomization and even distribution of samples across each plate, as described in one of the package vignettes published on CRAN [298]. Hence, we used version 3.4.1 of this package to do so and tested multiple seed definitions (with function olink_plate_randomizer()) until plate layouts and sample distributions yielded homogenous results (with functions olink_displayPlateLayout() and olink_displayPlateDistributions()).

For each of the 264 prepared and randomized serum samples, 50 µl were dispensed into its predefined well position on a 96-well PCR Plate (Thermo Fisher Scientific, catalog number: AB0800, Life Technologies, Darmstadt, Germany). All samples and each plate were cooled on flat ice blocks during pipetting according to the prepared randomization scheme. Ready-made plates were covered with adhesive film (MicroAmp Clear Adhesive Film, catalog number: 4306311, Life Technologies, Darmstadt, Germany) and frozen at –80 °C until transport to Helmholtz Munich on dry ice.

#### Olink Explore pipeline

The framework of the Olink Explore analysis pipeline has been described by Wik et al. [58]. In brief, the first step is to create several dilutions of each sample. This is necessary because serum proteins occur in a very large range of individual concentrations spanning over more than ten orders of magnitude (see Figure 3 in [299]). Therefore, probes for different magnitudes of protein abundance are pooled into blocks (so-called ‘abundance blocks’) and sample serum is diluted accordingly such that the orders of magnitude roughly match before all samples are being pooled for NGS. The required spectrum of dilutions depends on the assay panel being used. Currently, Olink Explore is set up to accommodate for the parallel analysis of around 3000 proteins (Explore 3072) in eight different assay panels which require five different serum dilutions (undiluted (1:1), 1:10, 1:100, 1:1000 and 1:100000) starting with a 1:10 dilution in the first six columns of a 384-well ‘sample dilution plate’ by picking 1 µl from each sample. The 384-plex inflammation assay panel (catalogue number 97500) itself however, only requires two different dilutions (1:1 and 1:10).

In a next step, an immunoreaction is miniaturized such that 0.2 µl of undiluted or diluted sample are brought together with 0.6 µl of incubation mixture containing forward and reverse PEA probes. Probe-protein binding and probe-probe hybridization takes place overnight at 4 °C, followed by a first PCR for probe extension, addition of a first adapter and pre-amplification. After this first PCR step, the four abundance blocks from each sample are pooled such that this first PCR product contains amplicons at similar concentrations from each of the abundance blocks. A second PCR serves for integrating index sequences such that the final PEA amplicons before NGS have a length of 148 base pairs and contain Illumina P5 and P7 adapters, read 1 sequencing primer site (Rd1SP), assay specific forward and reverse barcodes (FBC and RBC), hybridization site between probe arms (Hyb), and a sample-specific index. This is followed by a second and final sample pooling step during which all samples for each assay panel are being brought together – in our case, this means just the pooling of one full assay library before purification and quality control for correct library size (with a peak around 150 bp).

Next-generation sequencing of this library pool is performed according to the Illumina NovaSeq Xp workflow with their NovaSeq 6000 system on one S1 flow cell lane per assay panel (Illumina, Berlin, Germany). Single-read sequencing with a read length of 66 base pairs covers forward and reverse barcode, and the sample specific index. Run-to-run carryover is addressed by a maintenance-wash before every sequencing run. A PhiX control v3 library is added to the sample library before sequencing to balance the fluorescent signal as sequence diversity in Olink libraries is very low.

#### Olink Explore data output

Olink NGS generates sequence reads that contain both, assay and sample information. Only exact matches are included for further analysis. The raw NGS results file contains counts from each sample index matched with the different assay barcodes. Final results are expressed in so-called ‘Normalized Protein eXpression’ (NPX) values derived from matched sequence counts together with one (of three) internal and one (of three) external controls.

The internal control is a so-called ‘extension control’ which is added at a known concentration during the immunoreaction step. In contrast to the usual two PEA proximity probes of which each is coupled to an antibody, the extension control is made of two paired oligonucleotides coupled to a single antibody. This allows for reliable hybridization independent of antigen binding and therefore monitors variation in the extension and amplification step. It is used to adjust the signal from each sample with respect to extension and amplification.

The external control is a so-called ‘plate control’ which is added in triplicate in the last column of the sample plate. In contrast to the Olink Target platform, this plate control is pooled EDTA plasma originating from healthy blood donors supplied by Olink. The median of these plate control triplicates is used to normalize each assay and to compensate for potential signal variation between plates.

First, matched sequence counts for each specific combination of assay barcode and sample index are divided by the number of counts for the extension control with the same sample index and this resulting ratio is then transformed logarithmically. Second, plate control values which have also been normalized during the first step are used to correct for variation between plates by subtracting the plate specific median from every sample of the corresponding plate. This causes NPX values to become centered around zero. Third, intensity normalization may be added by correcting with the plate median. The resulting NPX values are relative quantification units which are logarithmically related to protein concentration in the sample.

Theoretically, 264 samples (= 216 + 48) samples analyzed with Olink Explore 384 Inflammation panel should generate 97152 NPX values for 368 different proteins. Results are currently pending.

### Axis V: Lipidome

#### Fatty acids from erythrocyte membranes (chromatography)

Analysis of fatty acid (FA) composition in erythrocyte membranes was performed with gas-liquid-chromatography (GLC, or simply GC) as previously described [65,300,301] and as has been common for a long time in the field of lipid analysis [302]. It requires the transformation of fatty acids into volatile fatty acid methyl esters (FAME) that can then be detected with GC, either coupled to a flame ionization detector (GC-FID), as described here, or a mass spectrometer (GC-MS). Assessment of fatty acid composition and the derivation of useful indices is highly method-sensitive [301]. For example, results depend on methylation conditions, extraction solvents, columns, peak identification conventions as well as various calibration and corrections measures. Therefore, three international laboratories in Munich, Germany, as well as in Seoul, South Korea and Sioux Falls, SD, USA, have standardized all stages of the analytical procedures and have been employing identical methodologies, sometimes also referred to as ‘HS-Omega-3 Index methodology’ as first mentioned by Harris & von Schacky in 2004 [65]. Omegametrix in Munich (Martinsried, Germany) is one of these reference laboratories for the analysis of fatty acid compositions from erythrocyte membranes were all analyses are quality-controlled according to DIN ISO 15189.

In total, 149 EDTA blood samples were analyzed, with 124 longitudinal samples originating from 67 patients (begin and end of trial) and 25 samples from 25 healthy controls. Fasting venous blood samples had been collected in the mornings between 7:00 and 07:30 hours before breakfast into tri-potassium (K3) ethylenediaminetetraacetic acid (EDTA)-containing containers (2.7 ml EDTA K3E S-Monovettes, Sarstedt AG & Co. KG, Nümbrecht, Germany). These EDTA tubes were immediately shipped to Omegametrix at ambient temperatures as suggested by the laboratory. These storage conditions, for up to five days, are supposed to have no relevant effect on fatty acid compositional results including the omega-3 index [303].

The complete set of samples was analyzed in different GC runs which also caused longitudinal samples from the same subjects to be measured in different runs. Analyses were performed between February 2016 and October 2020 for participating patients and between November 2020 and February 2021 for participating healthy volunteers. Analytical variability was assessed with a quality-control sample of pooled erythrocytes. Thus, the analytical coefficient of variation (CV) was 6-7 %.

After arrival at Omegametrix, red blood cells (RBCs) were isolated by removing plasma and buffy coat after centrifugation. An RBC sample was taken from the center of the remaining RBC sediment for storage at -80 °C until analysis.

RBCs were neither washed before storage nor after, nor were they going through dedicated lipid extraction before methylation. Hence, thawed RBCs were directly methylated following a recipe by Morrison & Smith [304]. Fatty acid methyl esters (FAME) were prepared by acid transesterification. Fatty acids were transmethylated with methanol-containing 14 % boron trifluoride (BF_3_) at 100°C for 10 min in a 1:10 mixture of RBCs and BF_3_. After cooling, the fatty acid methyl esters were recovered by adding hexane and distilled water in equal volumes as BF_3_, followed by shaking and centrifuging to separate layers. An aliquot of the hexane supernatant was analyzed by flame ionization gas chromatography using a GC2010 gas chromatograph (Shimadzu, Duisburg, Germany) equipped with a 100 mm SP2560 fused silica capillary column (0.25 mm i.d., 0.2 µm film thickness, Supelco, Bellefonte, PA, USA) and hydrogen as carrier gas.

Fatty acids were identified using a commercial standard mixture of 22 fatty acid methyl esters representative for erythrocytes (GLC-727; Nu-Check Prep, Elysian, MN, USA) after response factor adjustment (based on calibration curves and setting palmitic acid as 1.0) [305]. In this standard, individual C18:1 trans isomers (C18:1 d6 through d13) cannot be separated with this methodology but appear as two blended peaks that elute immediately before oleic acid. Hence, the areas of these two peaks are summed and referred to a C18:1 *trans*.

Fatty acid methyl ester composition is reported as weight percent of a total of 26 identified FAMEs in erythrocyte membranes. This set of 26 fatty acids comprises six saturated fatty acids (SFA), four monounsaturated fatty acids (MUFA), seven omega-6 polyunsaturated fatty acids (n6-PUFA) and four omega-3 polyunsaturated fatty acids (n3-PUFA). Four of the n6-PUFAs and three of the n3-PUFAs belong to the group of highly unsaturated fatty acids (HUFA). Furthermore, the set includes five *trans*-fatty acids (TFA) of which two are mono-*trans* and three are poly-*trans* fatty acids. Among the 26 fatty acids, 19 belong to the group of long-chain fatty acids (LCFAs, 13-21 C in aliphatic tails) and 7 to very long chain fatty acids (VLCFAs).

In detail, SFA: C14:0 (myristic), C16:0 (palmitic), C18:0 (stearic), C20:0 (arachidic), C22:0 (behenic) and C24:0 (lignoceric); MUFA: C16:1n7 (palmitoleic), C18:1n9 (oleic), C20:1n9 (gadoleic) and C24:1n9 (nervonic); n6-PUFA: C18:2n6 (linoleic, LA), C18:3n6 (gamma-linolenic, GLA), C20:2n6 (homo-gamma-linolenic), C20:3n6 (dihomo-gamma-linolenic, DGLA), C20:4n6 (arachidonic, AA), C22:4n6 (docosatetraenoic = adrenic, AdA) and C22:5n6 (docosapentaenoic, n6-DPA); n3-PUFA: C18:3n3 (alpha-linolenic, ALA), C20:5n3 (eicosapentaenoic, EPA = timnodonic), C22:5n3 (docosapentaenoic, n3-DPA = clupanodonic) and C22:6n3 (docosahexaenoic, DHA = cervonic); mono-TFA: C16:1n7t (palmitelaidic = trans palmitoleic), C18:1t (elaidic = trans oleic) – the latter comprises two blended peaks of C18:1(d6 through d13); poly-TFA: C18:2n6tt (9t,12t) (linoelaidic = trans,trans-linoleic), C18:2n6ct (9c,12t) (cis,trans-linoleic) and C18:2n6tc (9t,12c) (trans,cis-linoleic).

The omega-3 index is calculated as the sum of the two n3-PUFAs EPA and DHA expressed as a percentage of the total of 26 identified fatty acids in erythrocyte membranes. The so-called ‘trans index’ reflects industrially produced trans fatty acids and is calculated as the sum of all C18:1 and C18:2 *trans* species. It contains the two blended peaks described above (C18:1t) together with the three poly-TFAs (C18:2t) expressed as a percentage of the total of 26 identified fatty acids in erythrocyte membranes.

#### Lipid species from serum (mass spectrometry)

High resolution mass-spectroscopic (HRMS) quantitative shotgun lipidomic analyses from serum were performed by direct flow injection analysis (FIA) using a high-resolution (HR) Fourier Transform (FT) hybrid quadrupole—Orbitrap mass spectrometer (MS) QExactive (Thermo Fisher Scientific, Bremen, Germany) equipped with a heated electrospray ionization source. The applied setup has previously been described in detail [306,307].

In total, 214 serum samples were analyzed which had been taken in the mornings between 7:00 and 07:30 hours before breakfast: one hundred and eighty-nine longitudinal serum samples from 68 patients over three time-points (study days 1, 11 and 60) and 25 samples from 25 healthy subjects. Fasting venous blood from patients and healthy participants had been collected into serum containers (Serum CAT S-Monovettes, Sarstedt, Nümbrecht, Germany) and were immediately centrifuged at 1516 g for 10 min at precooled 8 °C before long-term storage at –80 °C in micro tube aliquots until analysis.

Serum samples were first spiked with lipid species which do not naturally occur and can therefore serve as internal standards (IS). The following lipid species were added as such internal standards: CE 17:0, CE 22:0, Cer 18:1;O2/14:0, Cer 18:1;O2/17:0, DG 14:0/14:0, DG 20:0/20:0, FC[D7], GlcCer 18:1;O2/12:0, GlcCer 18:1;O2[D5]/18:0, LPC 13:0, LPC 19:0, LPE 13:0, PC 14:0/14:0, PC 22:0/22:0, PE 14:0/14:0, PE 20:0/20:0 (di-phytanoyl), PI 17:0/17:0, PS 14:0/14:0, PS 20:0/20:0 (di-phytanoyl), SM 18:1;O2/12:0, TG 17:0/17:0/17:0, and TG 19:0/19:0/19:0.

Lipid extraction was performed according to Bligh and Dyer [308] in the presence of spiked IS. A serum volume of 10 µl was used for extraction with a total chloroform volume of 2 ml. Chloroform phase was recovered by a pipetting robot (Tecan Genesis RSP 150, Tecan Group, Männedorf, Switzerland) and vacuum dried. Residues were dissolved in chloroform/methanol/2-propanol (1:2:4 v/v/v) with 7.5 mM ammonium formate. All solvents were HPLC grade.

In total, 189 lipid species were analyzed for each sample. For lipid quantification with high-resolution FIA-FTMS, triacylglycerols (TG, 46 species), diacylglycerols (DG, 11 species), and cholesteryl esters (CE, 13 species) were recorded in positive ion mode FTMS in *m/z* range 500−1000 for 1 min with a maximum injection time (IT) of 200 ms, an automated gain control (AGC) of one million, three microscans, and a target resolution of 140000 (at 200 *m/z*). The mass range of negative ion mode was split into two parts: lysophosphatidylcholines (LPC, 12 species) and lysophosphatidylethanolamines (LPE, 7 species) were analyzed in an *m/z* range of 400–650. Phosphatidylcholines (PC, 27 + 14 (ether) species), phosphatidylethanolamines (PE, 9 + 8 (ether) species), phosphatidylinositols (PI, 13 species), sphingomyelins (SM, 21 species) and ceramides (8 species) were measured in an *m/z* range of 520–960. Multiplexed acquisition (MSX) was used for [M + NH_4_]^+^ of free cholesterol (FC) (404.39 *m/z*) and for the corresponding internal standard FC[D7] (411.43 *m/z*) using a setting of 0.5 min of acquisition time, with a normalized collision energy of 10 %, an IT of 100 ms, an AGC of one hundred thousand, an isolation window of 1 *m/z*, and a target resolution of 140000 [306].

Data processing details have previously been described [307] using ALEX software [309] for peak assignment and intensity picking in FTMS- and MSX-spectra. Data were then transferred to Microsoft Excel 2016 for automated processing with self-written macros in Excel Visual Basic for Applications (VBA). Among other tasks, these macros are written to correct for Type-I and Type-II isotopic effects.

The quantification step comprised multiplication of spiked IS amounts with analyte-to-IS intensity ratios after isotope correction. Lipid species were annotated in conformance with the latest proposal for shorthand notation of lipid structures derived from mass spectrometry [310].

Accurate quantification of lipid species via FIA-FTMS requires correction with individual response factors. Differences in responses between lipid species originate from structural features such as length and double-bond number of acyl chains, both of which were hypothesized to be in linear relationships for the model used for response correction. Calculation of lipid species-specific response factors has been described previously [306].

### Axis VI: Psychometrics

#### Overview of assessment scheme

All clinical assessments including the verification of inclusion and exclusion criteria as well as observer ratings were performed by five, and primarily by two experienced psychiatry residents supervised by two senior attending psychiatrists on a specific ward dedicated to clinical research on depression. All of them had regularly been trained and certified in Good Clinical Practice (GCP) and in the accurate application of the corresponding psychometric tools including supervised rater trainings.

Over the study course of eight weeks, there were ten primary study visits that included observer ratings and self-reportings on study days 1 (entrance of study), 5 (beginning of medication), 11 (last day of double-blind medication period), 18 (weekly continuation), 25, 32, 39, 46, 53 and 60 (end of study). Overall, twelve psychometric tools were used in this trial of which eight were applied every week. Their primary foci were depression and anxiety supplemented by handedness (as relevant for imaging experiments) and a few aspects of personality. Four of these tools were observer ratings while the other eight were self-reports. Participants were introduced to each of them in advance with guidance for appropriate self-reporting. Four observer rating tools (CGI, HAMD21, HAMA, MADRS) and four self-reporting tools (VAS, BDI-II, SHAPS-D, STAI) were employed during each of these ten study visits and are described in more detail, below.

Additionally, a 4-item short version of the Edinburg Handedness Inventory (EHI) self-rating was used on the first day of study while the regular 10-item version was chosen for the first day of imaging on the fifth day of the study. For the same day, the Barrat Impulsiveness Scale (BIS) with 30 items and the Temperament and Character Inventory (TCI) with 107 items were planned but usually given to patients the day before to lessen the load of research activities during the first day of imaging. This made for seven self-reports for study day five.

#### Psychiatric diagnosis and history

Some medical history questions were repeated in a structured self-report questionnaire asking for native language, gender, handedness, highest achieved levels of education and vocational training, possible twin status, self-assessment in regard to addiction and which potentially addictive substances have been consumed. Furthermore, it asked about age at onset of first depressive episode, number of previous depressive episodes, duration since last depressive episode and number of previous psychiatric admissions. Finally, the questionnaire asked for a detailed family history including parents, siblings, grandparents, as well as uncles and aunts.

#### Psychometric tools

##### Observer ratings

Four different semi-structured psychiatric observer ratings in German language were performed ten times during the course of the study and took around 60 to 80 min during each visit depending on patient condition. Severity of disease, change in disease severity and drug efficacy were evaluated with the Clinical Global Impression scale (CGI, [311]) from 1976 which contains four items (first two items with 8 levels (0 to 7) and last two items with 5 levels (0 to 4).

Changes of depression severity were assessed with the standard 21-item Hamilton Depression Rating Scale (HAMD21, [312,313]) as well as with the Montgomery-Åsberg Depression Rating Scale (MADRS, [314]) while changes in anxiety states were assessed with the Hamilton Anxiety Rating Scale (HAMA, [315].

The Hamilton Depression Rating Scale from 1960 with 21 items (HAMD21) consists of the original 17-item scale by Max Hamilton [312] extended by four additional items which he had excluded in the beginning but described them as of potential interest in research, later [313]. Ten items are built of five levels, ten of three and one of four. Maximum score is 63 and minimum score is 0 points. The higher the summed score, the higher disease severity is considered in comparison to previously taken scores. “Markedly ill” as described by a CGI severity score of 5 points corresponds to a HAMD17 sum score between 24 and 26 points [316]. A HAMD21 sum score of below 8 points has been considered as normal range or remission [317].

The Montgomery-Åsberg Depression Rating Scale (MADRS) from 1979 is a 10-item scale in which each item has 7 levels (0 to 6). Maximum score is 60 and minimum score is 0 points. The higher the summed score, the higher disease severity is considered in comparison to previously taken scores. “Markedly ill” as described by a CGI severity score of 5 points corresponds to a MADRS sum score between 31 and 34 points [318]. A MADRS sum score of below 8 points has been considered as normal range or remission [317].

The Hamilton Anxiety Rating Scale (HAMA) from 1959 is a 14-item scale in which each item has 5 levels (0 to 4). Maximum score is 56 and minimum score is 0 points. The higher the summed score, the higher symptom severity is considered in comparison to previously taken scores. In depressed patients, HAMA cannot discriminate between patients with or without additional syndromes of anxiety [319]. However, it has the capacity to reflect significant clinical improvements that may, for example, be caused by antidepressant therapy and, thus, may support response information.

Different frames of reference in time were applied for these observer ratings. The initial visit referred to symptomatology during the last two weeks before hospital admission while the remaining visits referred to each previous visit such that the second visit referred to the shortest period of time of four days while it usually were seven days for all others.

##### Self-reportings

Eight different self-report questionnaires in German language were used during the course of the study of which four (VAS, BDI, SHAPS, STAI) were handed out one day prior to each of the ten primary visits during the course of the study. Equal to observer ratings, participating patients were asked to apply different frames of reference in time for these self-reports depending on the study visit as described above.

A Visual Analogue Scale (VAS, [320,321]) was deployed with three items (mood, inner tension, fatigue), each represented on a discontinuous scale of nine levels (1 to 9) between extremes with the number 5 as middle of the scale indicating ‘balance’.

The revised version of the Beck Depression Inventory (BDI-II, [322]) from 1978 contains 21 items with 4 levels each (0 to 3). Maximum score is 63 and minimum score is 0 points. The higher the summed score, the higher disease severity is considered in comparison to previously taken scores. “Markedly ill” as corresponding to a HAMD17 sum score between 24 and 26 points can be converted to BDI-II scores of 40 to 44 while BDI-II sum scores below 10 behave like HAMD17 scores such that a BDI-II sum score below 7 points may be considered as normal range or remission [323].

The German version of the Snaith-Hamilton Pleasure Scale (SHAPS-D, [324]) from 1998 contains 14 items. Even though each item offers four levels of answers (ranging from “definitely agree” to “strongly disagree”), the scale does not use a Likert-style scoring but assigns 1 point for all two “disagree” levels and 0 points for two “agree” levels. Hence, maximum score is 14 and minimum score is 0 points. The higher the summed score, the stronger anhedonia is considered.

The German X1-anxiety state form from Spielberger’s State Trait Anxiety Inventory (STAI, [325]) from 1981 contains 20 items with 4 levels each (1 to 4). 10 items are phrased with negative connotation and 10 with positive connotation. Maximum score is 80 and minimum score is 20 points. The higher the summed score, the higher the current state of anxiety is considered. A STAI-X1-state sum score below 40 points may be considered as normal range or remission [326].

Version 11 of Barrat Impulsiveness Scale (BIS-11, [327,328]) from 1995 contains 30 items with 4 levels each (1 to 4). Its German translation was evaluated in 2007 [329]. Eleven items asking for non-impulsivity need to be reverse scored. Maximum score is 120 and minimum score is 30 points. The higher the summed score, the higher the level of impulsiveness is considered. Total sum scores between 52 and 71 are considered to be within normal range of impulsiveness while 72 or above is considered highly impulsive [330]. However, extensive factor analytic work over decades showed that only reporting the total sum score of this scale does not reflect impulsivity as the multi-faceted construct that it appears to be according the developer of the scale. Therefore, it was recommended to additionally and at least report subscores from the three second-order factors, namely “Cognitive Impulsiveness”, “Motor Impulsiveness” and “Attentional Impulsiveness” (previously “Non-Planning Impulsiveness”) [330].

The four temperament dimensions of the Temperament and Character Inventory (TCI, [331]) from 1994 with 107 items (out of 240 for the whole inventory) were used in its German translation from 1999 [332]. These temperament dimensions contain “Novelty Seeking” (NS) with 40 items on four subscales, “Harm Avoidance” (HA) with 35 items on four subscales, “Reward Dependence” (RD) with 24 items on four subscales and “Persistence” (P) with 8 items on a single subscale. Each item requires a “yes” or “no” answer which is provided by setting a mark in the corresponding column. “yes” relates to 1 point and “no” to 0 points and 58 items need to be reverse scored (23 NS, 20 HA, 12 RD, 3 P). Maximum total score is 107 and minimum score is 0 points while the total sum average in a German normative sample was 54 points [332]. Additionally, Richter et al. matched healthy individuals from their German normative dataset to 126 depressed inpatients [333] who primarily showed significantly higher scores in the harm avoidance dimension (for example 23.4 ± 4.7 for patients with depression at admission versus 15.7 ± 5.5). Reward dependence axis also showed relevant differences, however, at the subscale level, only. Very similar scale results were reproduced later in a Turkish cohort by Celikel et al. [334].

The Edinburg Handedness Inventory (EHI, [335]) from 1971 contains 10 items with 5 levels each for everyday activities like brushing teeth that require one hand only. Scores range from -100 for extremely left-handed in all specified activities to +100 for extremely right-handed. Criticism of the original inventory has led to various modifications, extensions and reductions [336] with one of them being a 4-item short form by Jamie F. Veale [337] containing four of the ten original items (writing, throwing, toothbrush and spoon). Instructions and response options are slightly different, for example, the 5 levels are represented on a Likert-type answering scale. Still, the laterality quotient is calculated according to the same principle and yields the same ranges as described for the standard 10-item version.

#### Estimating clinical response behavior

Two approaches will be used for estimating clinical response behavior from psychometric data that has been acquired longitudinally over ten time-points according to protocol.

One variant is ‘traditional’: it omits the majority of performed psychometric assessments (here, 8 of 10) and focuses on a static threshold for ‘responder’ definition which is arbitrarily chosen as a decrease in the sum of rating points, usually by 50 % from baseline to end of trial.

Another way is take all available psychometric data into account and to model clinical response curves over the whole period of the trial implementing non-linear decline properties and sufficient freedom for curve shape. Thus, response behavior can be estimated and distinguished in much more granular nuances like response dynamics which should provide considerably more relevant information in relation to performed investigations.

The ‘traditional’ variant will be included for historic reasons while the investigational focus will be on appropriate modelling of the fully available response information from the trial.

### Axis VII: Co-enrollment in other clinical depression trials

#### Introduction

Axis VII is not an inherent axis of investigation of this trial. It appears reasonable, however, to summarize the participation in concomitant trials, here, to facilitate a comprehensible overview of research procedures that were additionally performed with some or all participants of this trial within the trial recruitment period or immediately after.

#### OptiMD SP5 crosslinking

It is considered good practice to crosslink multicenter projects within large research consortia like OptiMD of which this trial is a part. The complexity of the trial design described here caused us to primarily focus on a single-center approach, though, for reasons of feasibility and validity.

However, this trial has been planned with a strong focus on neuroimaging which made it a natural and reasonable choice to crosslink and fully implement the OptiMD neuroimaging branch (subproject SP5, led by co-author OG in Heidelberg) into our planned efforts. The title of this subproject is “MRI biomarkers for the prediction of individual responses to various antidepressants” and some of its scanning protocols are also employed in other German Research Consortia on mental disorders, like BipoLife [151].

Inclusion and exclusion criteria for OptiMD SP5 were completely covered by the criteria of this trial, which were set to be considerably stricter than for SP5 and only included inpatients while SP5 also allowed for outpatients.

Specifically, inclusion criteria for SP5 comprised drug-naïve patients who were prescribed antidepressant monotherapy with either citalopram, escitalopram, agomelatine, mirtazapine or venlafaxine. Patients were then going through the scanning protocol once at baseline before starting medication without further imaging afterwards.

The OptiMD SP5 MRI scanning protocol was similar to the protocol recently described for the German Research Consortium BipoLife [151] and consisted of a structural localizer, a T1-weighted high-resolution anatomical image, three task-based fMRI paradigms, a resting-state functional MRI (fMRI) sequence, a field map and a gel-phantom measurement for quality assessment.

OptiMD SP5 was fully implemented into this trial and all participants of this trial were co-enrolled in it. Therefore, description of its details has been implemented in chapter “Axis I: Neuroimaging”.

#### FZPE PD-CAN crosslinking: Mini-RDoC test battery – Domain Assessment Network Germany

OptiMD is one of several research consortia within an even larger national program, called *Forschungsnetz für psychische Erkrankungen* (FZPE, Research Network for Mental Disorders). A research project within this national FZPE program, called Phenotypic, Diagnostic and Clinical Domain Assessment Network Germany (PD-CAN) had been installed as an independent crosslinking project [338] encompassing all research consortia.

Crosslinking therefore included research consortium OptiMD and its clinical subprojects and hence, this trial. PD-CAN distributed a Mini-RDoC test battery that contained 18 parts (16 tests) structured into three different components along a shell model with required core variables and variables from two optional shells (as described in detail by Förstner et al. in [338]). Our study center was asked to provide Mini-RDoC test batteries for 35 patients with depression for which we fulfilled all three parts of the shell model between January 2018 and December 2019. Of these 35, 13 have been participating in this trial. There had not been a requirement to acquire the data immediately after admission to hospital, but 10 of these 13 patients provided the test batteries at baseline measurements of this trial.

#### Blood markers for inflammation and cardiovascular risk in depression

An ongoing clinical trial at our study center on inflammation and cardiovascular risk in depression [339] as described in section “Human subjects and recruitment” above, had entered recruitment phase in December 2012, a little more than three years before the beginning of recruitment for this trial. It was designed as a successor to a trial that had been recruiting from 2004 to 2008 and focused on genetic and inflammatory markers as risk factors in patients with cardiovascular disease suffering of clinical depression [340,341]. The successor trial focused on proteomic inflammatory markers in patients with clinical depression including possible mediators like altered lipid profiles. Investigation of these risk factors was accompanied by measuring functionality of the HPA-axis with two dex/CRH-tests at baseline and end of a six-week trial course while patients were ‘treated as usual’, making this a field observation trial.

All participants from the trial of this protocol were co-enrolled for the trial above for the majority of blood biomarker investigations which had strong overlap with the goals of this trial. Blood was drawn into three containers for this trial during research or routine blood draws on study days 1, 11 and 46, sparing participants any additional invasive procedures. Samples were provided by 66 patients at the beginning of the trial.

#### Skin biopsies for a cellular model of depression

Another ongoing trial at our study center on mitochondrial dysfunction in cells reprogrammed from fibroblasts biopsied from patients with depression [342] had entered recruitment phase in December 2014, more than one year before the beginning of this trial. Its focus was to better understand cellular bioenergetics in clinical depression.

Towards discharge of participants from inpatient care and, thus, towards the end of this trial, seven patients that had shown particular interest and enthusiasm in research participation were asked about possible further study participation with a skin biopsy. At that point, they had recovered substantially with mostly alleviated depressive symptoms. Thus, patients with very recent trial experience and a much healthier state of mind were able to clearly decide about an invasive procedure asked of them. All of these patients provided well-informed written consent while they were still participating in this trial. One provided a skin biopsy during this trial, another one week after end of this trial and five about a month after its end. These seven participants have been labelled with index numbers 10 to 16 in supplementary table 1 in [342]. They probably are the participants of this trial that have been phenotyped the deepest altogether. With exception of one participant who did not participate in dex/CRH-tests, all seven participants have fully contributed in all investigational axes of this trial over all time-points including the other co-enrolled trials. For microbiome samples, only OMNIgene.GUT storage is available for all time-points.

### Data management

#### Clinical data management

All trial data has been managed by the research group of the principal investigator at the Department of Psychiatry and Psychotherapy, Universität Regensburg, Regensburg, Germany.

Case Report Forms (CRF) for the whole duration of the study had been prepared in print and were stored in dedicated folders per participant. A printed CRF was structured chronologically in chapters by study visit in line with the study protocol. It contained the signed consent form, a study visit check list, all questionnaires from all subprojects, study related vital parameters including ECG, pharmaceutic safety data including adverse event forms, MR safety data and open sections for protocol deviations, drop-out reasons or additional information that was judged relevant to the study like description of peculiarities that might explain aberrant study results, later. All other source data were transcribed from the integrated hospital information system or collected for digital long-term storage until further data curation.

Additionally, an SPSS statistics data file (IBM SPSS Statistics for Windows, Version 23.0, released 2015, IBM Corp., Armonk, NY) had been prepared in advance in order to primarily digitize psychometric results and vital parameters, structured in correspondence to the study protocol. Digitization and logging of data was transferred to and continued in Microsoft Excel 2016 (Microsoft Corporation, Redmond, WA, USA) during the course of the trial. Data curation has been ongoing in Excel, as well, from where data will usually exported in CSV-format for import into other analysis software.

#### Dedicated server and archive infrastructure for imaging data

Reliably preserving, archiving and handling of original MR research imaging data was of high priority in context of data management. Therefore, a dedicated server and archive infrastructure was established for this trial such that MR images in raw Siemens Digital Imaging and Communications in Medicine (DICOM) format could directly be sent from the MR scanner for long-term storage without any modifications to the original DICOM data as produced and sent by the MR scanner control. The complete server infrastructure and storage environment was embedded on premises in the core data center of our IT department which primarily hosts all health data and services for the clinical neuroscience campus and its satellites. Hence, all research imaging data from this trial were handled, stored and backed up with the same strict safeguards and data protection policies that apply to clinical patient data. This approach provided an appropriate safe space for research imaging data before quality control and full anonymization of metadata and images at a later time.

While in theory, the choice of receiving operating system should not influence the data, it was specifically made conformant to the Siemens MR control operating system which used Windows 8 or higher. Therefore, a VMware-Virtual Platform-based server (VMware, Palo Alto, CA, USA) was installed with Windows Server 2012 R2 using an AMD Opteron 6380 (Quad 2.5 GHz, 2 processors, with 16 GB RAM). On this platform, Pmod software [343] base module (Pmod Technologies, now a Bruker division, Fällanden, Switzerland) was used to set up a scientific data management system (SDMS) which includes the functionalities of a standalone Picture Archiving and Communication System (PACS) [344]. Pmod DICOM and transaction servers were permanently running as controlled Windows Services by using an additional software as recommended by Pmod (AlwaysUp, Core Technologies Consulting, Oakland, CA, USA). This setup provided a robust and reliable SDMS environment which has been running without errors or unplanned interruptions since its installation. Furthermore, Pmod SDMS offers configurability such that incoming data is explicitly saved as received without any modifications. This feature was considered a major prerequisite for data preservation and long-term archiving.

Thus, at the end of each MR scanning session, all acquired imaging data were sent to the Pmod DICOM server and additionally saved directly to DVD. Data on the Pmod server were incrementally backed up very evening and fully backed up to tape once a month.

### Data analyses

The scope and complexity of this trial design deems a staged analytical approach. It will begin with descriptive data analyses of demographic data and base parameters of single research modalities as well as using simplest possible test procedures at first, for example to retrieve and compare initial point estimates and to gain insight into distribution of data. Furthermore, the impact of missing data will be examined with sensitivity analyses. Initial analyses will focus on intention-to-treat and then continued with analysis of valid cases.

The type of data that have resulted from the investigations are described for each experiment in the individual axes of investigation, above, including primary possibilities for their analyses.

Generally, initial analyses will focus on cross-sectional data at baseline as a guidance towards characteristics of the recruited cohort. This should be possible with t-tests for mean value differences. A primary goal will be to test for differences between the four randomized groups over the first two or all of the three major longitudinal time-points of this trial, depending on the experiments. In a first approach, this will be tried with repeated-measures ANOVA depending on characteristics of the data. In further stages, various observations will be clustered at several levels in order to develop appropriate statistical solutions for multilevel modeling.

Different software tools will be used depending on the analyses. With certainty, R (R Core Team, 2025, Vienna, Austria) and Python (Python Software Foundation, 2025, Beaverton, OR, USA) languages together with various required packages and libraries will be employed, including larger repositories like Bioconductor (Bioconductor Core Team, 2025, Seattle, WA, USA). These languages will most likely be controlled and scripted in various IDEs (integrated development environments). Furthermore, statistical analysis software like IBM SPSS Statistics as well as Matlab (The MathWorks, Natick, MA, USA), Matlab software suites like SPM as well as FSL (FMRIB Software Library, University of Oxford, UK) might be used. Also, Pmod (Pmod Technologies, a division of Bruker, Fällanden, Switzerland) might be used for processing imaging data, at least for pre-processing steps. Applied tools will properly be reported for the corresponding analyses.

## Discussion

### Introduction

For many decades many researchers across the world have been working on a better understanding of mechanisms behind clinical depression and potentially improved therapeutic interventions. Our comprehension of them seemingly becomes less the more we know in consideration of the increasingly growing complexity and diversity in perspective to this illness. Hence, this trial will, at best, add but miniature pieces to a puzzle that appears far from being completed for a while, still.

There are different approaches to the conceptualization and design of deep phenotyping studies for a better understanding of affective disorders like clinical depression. Ours has been a single-center trial with relatively low numbers as compared to other ambitious trials [345] in this regard. These numbers were still manageable for our study center over a recruiting period of five years. The recruitment phase has successfully been finished without complications despite the extent and complexity of study measures. The majority of patients and volunteers provided positive feedback and appeared to be fine with their participation during the trial and in hindsight. The trial has not yet been called to an end as databases still remain open and some raw data of analyzed biomaterials are pending return.

Even though the first steps in trial design began more than thirteen years ago, it appears that our final study protocol follows state-of-the-art quality even today including highly sophisticated methods for analysis of materials. We are not aware of single-center clinical trials that have endeavored to combine this many modalities of investigation in the same patients that are deemed relevant in clinical depression.

Our angle for this clinical trial on depression has been multi-facetted. Appreciating the participation of humans suffering from depression in an ethical context meant that even an exploratory trial of this complexity should reliably provide results, at least by confirming recently published data and by employing novel technologies. Moreover, combining an extensive set of research modalities within the same human subjects in a longitudinal design should hopefully add new perspectives on already present knowledge.

The majority of study protocols for clinical trials is published at earlier stages, often during the first half of their recruitment phase or even before. This often means that many aspects of trial recruitment, of biomaterial logistics and methodological details of planned analyses are not yet precisely known. Hence, the final state of a study protocol can usually not be reported at these stages. In contrast, this protocol reflects a state that is rather close to the end of trial as defined by completeness of databases. It should not be long before databases will be closed and after all biomaterials have gone through laboratory analyses as planned per protocol – in many parts, only as planned and implemented during amendments and after granted raises for funding during the recruitment phase. It therefore provides the opportunity to report a realistic trial performance including known deviations from protocol. Moreover, all of the investigational methodology has fully been applied by now and its detailed description, here, should serve as a comprehensible reference for future dissemination of results. Furthermore, the state of this protocol can address aspects of criticism that could not have been formulated during the initial phases of recruitment and which usually cannot appropriately be addressed during dissemination of results.

The majority of clinical trial designs are based on compromises, including this study protocol. It should implicitly be clear that we see many strengths and value in our study protocol as described above. The discussion therefore focusses on its potentially critical aspects.

### Length of this protocol

The length of this study protocol has several reasons. These are, on one hand, related to the design of this trial. On the other, they reflect our endeavor for scientific comprehensibility and reproducibility.

First and foremost, the investigational volume of this trial design is extensive and complex: 14 experiments have been performed in the same participating patients, of which 10 experiments have been longitudinal with at least two or even three time-points. All of them were accompanied by 10 regular psychometric assessments over the full course of the trial, supplemented by 5 at baseline. Additionally, 3 co-enrollment experiments have been performed on parts of the study cohort and matched healthy subjects have passed the full investigational program of patients at one time-point, except dex/CRH-testing. Hence, in this deep-phenotyping approach, many different experiments have been performed with the same participants within the same project. In a conventional clinical trial setting, this would correspond to multiple separate study protocols

Second, this study protocol has a high level of completeness and reflects a nearly final state of a clinical trial of which the recruitment period has successfully ended, including two amendments, and of which databases are expected to be near closure. Therefore, all relevant details, including applied methodology, are made available for future reference, here.

Third, while the principles of methodology applied for this trial have previously been published, the majority of methods as applied here, have largely not been published, before. Therefore, they most often represent the only and primary reference of future disseminations of results from this trial.

Fourth, the completeness of applied methods described here, is meant to strengthen scientific transparency and thus scientific reproducibility. From our perspective, properly described methods create the foundation of good science. This appears particularly necessary in light of commonly short method sections for growingly complex methodology that even experts can only decipher in parts and which often are additionally positioned in supplementary materials, only.

The latest CONSORT 2025 statement [346] published on April 14, 2025, reminds clinical experimenters of exactly this scientific attitude. It cites a comment by Douglas G. Altman on the first CONSORT statement from 1996: “Readers should not have to infer what was probably done; they should be told explicitly.” [347]. There, he also says “The CONSORT statement means that authors will no longer be able to hide inadequacies in their study by omitting important information“. Length in text is a natural consequence of fulfilling such reasonable scientific standards.

Fifth, and related to the previous paragraph, description of the investigational axis for microbiome analysis is the longest for several reasons. The trial described here has received major public funding for analyzing the gut microbiome in clinical depression. This is only one of the reasons, why we aspired to appropriately describe a complete gut microbiome analysis workflow as performed for this trial. While this workflow is no longer up-to-date in all of its details, it reflects an accredited state of the time that all samples of this trial were processed and sequenced for the first time. Furthermore, most of methodology in context of microbiome analyses is usually omitted in publications, implying that experts in the field would still be able to reproduce. This however is mostly not true, as we know from many of our own attempts to do so. We intended to contrast this substantial lack in method reporting by providing a complete description here.

Sixth, readability and comprehensibility for experts from very different disciplines has been another goal of this protocol. The trial design and analysis of its results requires substantial multidisciplinarity. Therefore, the expertise of several collaborators of this protocol ranges from analytical chemistry over molecular biology to brain imaging, mathematics and clinical psychiatry. This protocol is written such that these and all other interested experts and potential collaborators should be able to grasp all details and the background of this trial. Again, here, completeness enables readability but causes length.

As an additional note: to improve readability of this protocol, introductory sections have been separated into two categories and positioned in different places of this protocol: those that introduce the background and goals of this trial were positioned in the introduction as should be expected. Furthermore, a number of method-related introductions have been intertwined in the description of investigational axes to improve their context and readability, and hence, a better understanding of the methods presented.

### Trial design

#### Study setting and sample size

This trial has been performed by and within the boundaries of a single clinical study center. The complexity of the trial let us omit a multi-center design for reasons of feasibility and validity. Additionally, extensive requirements for resources and specific expertise did not reasonably allow for a national multicenter approach of this scope at the time of conception.

It is likely that extent and complexity of this trial caused a selection bias of participants. While suffering of clinical depression that caused them to be admitted to a hospital, patients still needed to be able to show interest in our trial and show remaining mental capacity to take part in it. This particularly concerned a required base level of concentration for participation in the extensive neuroimaging axis and its experimental tasks. Furthermore, they needed to be physically healthy and without relevant psychiatric comorbidities. This kind of selection is far from being naturalistic.

The design and approach of this trial is exploratory, only. Hence, causation and function are not targets of this study. The extent, complexity and strictness of inclusion criteria did not make a larger sample size reasonably possible at a single center like ours, even though it is a large one. Sample sizes are at the lower limit of what one might wish for even though trials have proven new insight with even smaller sample sizes. Furthermore, 80 patients were planned to be randomized which was hampered by changes in personnel and the COVID-19 pandemic within an already approved extension of recruitment, leaving 70 participating patients of whom seven dropped out and one was not randomized. Hence, we mostly achieved our goal of 60 completers, though not for all modalities in combination. Moreover, while we managed to perform MR imaging with 64 patients in total, nine of these were scanned on two other scanners due to retrofitting of the latest research MR device, leaving 55 on a single scanner. An overview of available raw sample numbers originating from this trial is presented in Table 2. These samples have not yet passed data quality control, hence valid samples numbers will presumably be a little less.

The consequences of sample size in this study protocol are multi-facetted. The longitudinal design of this trial increases statistical power. Still, it will sometimes not be sufficient when applying conventional statistical methods, especially when trying to combine research modalities or when evaluating multiplex marker panels.

The exploratory aims of this trial possibly comprise the most exciting questions as they are related to the extensive multi-modal nature of this trial design. Our deep-phenotyping approach might additionally enable an exploratory look at potentially sixty N-of-1-trials as a possible alternative of reporting. Furthermore, contemporary unsupervised machine learning methods might provide helpful novel support.

#### Eligibility and recruitment

Formal eligibility criteria were kept to a moderate level of strictness to enable reasonable recruitment. In everyday clinical study routine, the two central study doctors however were asked to be considerably stricter than the criteria would have allowed for in order to keep interfering variables to a minimum. For example, patients with a depressive episode on grounds of a bipolar disorder were eligible by protocol but none of the included participants showed signs of a bipolar disorder.

Also, no formal score level of total HAMD21 results had been set as an entry condition, but an open definition of medium to severe degree of clinical depression with a strong safety requirement in regard to suicidal thoughts and behavior. Study doctors interpreted this consistently well such that 90 % of included patients showed a total HAMD21 score above 20 at baseline with a median around 27.

Furthermore, it would have been possible to recruit patients with a previously ongoing antidepressant treatment if there had been a clinical requirement for switching medication and in case patients would have been willing to pass a flush-out period instead of a cross-tapering regimen. However, to our knowledge there has only been a single patient arriving with a pre-treatment with citalopram 10 mg which had already been begun to be flushed out on another ward before this patient was approached for study participation. In consequence, there was a flush-out period of nine days before beginning of study medication. All other patients have entered this trial truly drug-naïve, which we consider as one of the strengths of this trial.

Microbiome sampling is a central investigational axis of this trial. In regard to microbiome sampling, formal criteria had also been kept open and only asking for an absence of antibiotic treatment without timeline and uncommon dietary habits. In daily clinical study routine, however, antibiotic treatment was not accepted within the last 6 months, at least. Depending on the antibiotics used, this period could have even been longer. Furthermore, a more strict history of food intake and body weight changes was taken. Recent dietary changes, changes in weight beyond 3 kg within the last three months, suspicious food supplements, nutritional intolerances and possible inflammatory syndromes including non-celiac gluten sensitivity were taken into account and usually not accepted.

#### Short duration of pre-post period and interaction of effects

The essence of this trial is to explore early antidepressant treatment effects with multiple modalities of state-of-the-art technology in the same individuals within the first week of antidepressant medication. Even though a number of trials have shown relevant effects in this short initial time span, even within a few hours or a day, it remains a risk in identifying differences between measurements at baseline and the first week, for example for microbiome analyses. This has been one of the reasons for a third time-point at the end of trial which should enable higher contrasts for research modalities that are not necessarily expected to react within the first week of medication.

Setting this primary and short investigational period within the first days of admission meant that various effects of change of environment would interact with the analyses. For example, patients with clinical depression often experience significant initial relief of their symptoms directly after admission to hospital, most often followed by worsening towards their admission state within a few days afterwards. Hence, we would expect to observe these effects between psychometric assessments at study visits on study days 1 (baseline-start), 5 (baseline-pre) and 11 (1-week-post). Moreover, the daily environment and rhythms of patients, their food and human interactions change within minutes after admission.

Furthermore, while this short primary study period required strict timing of investigations, it still caused a potential of interaction between the effects of investigations themselves. This primarily affected influences by dexamethasone/CRH-testing, particularly considering the long biological half-life of dexamethasone. Each of these tests was set to make for an interim period of two days before neuroimaging. While this interaction was constant in time for both time-points (pre and post), it was not for all other investigations. Per protocol, the first microbiome sample was taken about 8 to 12 hours after dexamethasone intake in the morning of the first dex/CRH-test. However, in about one third of patients, the first microbiome sample was taken on the first study day, together with all blood baseline measurements. The second stool sample was planned together with all other post-measurements, i.e. two days after the second dex/CRH-test. Additionally, initial lipid and inflammatory proteomic profiles from blood taken at baseline have not been influenced by dexamethasone intake or dexamethasone/CRH-testing while the post-measurements two days after the second dex/CRH-test may have been. In retrospect, some of these interactions could partly have been remedied by choosing different time-points for certain investigations, for example, by taking the first microbiome sample on study day 4 instead of 2, similar to scheduling of MR neuroimaging.

In regard to seven days of double-blind study medication in which an active substance could either be contained in the morning or in the evening, this also meant that patients might only have had six days of study medication (mirtazapine or agomelatine group) before the second test battery on study day 11 in comparison to those who had received escitalopram in the morning of that day. In retrospect, this discrepancy could partly have been remedied by beginning study medication in the evening of study day 4.

#### Doubling of two experiments

Most aspects that require discussion in regard to the investigations performed for this trial will individually be addressed in corresponding dissemination of results. A few aspects in regard to trial design should however rather be mentioned, here, within this complete overview.

The neuroimaging axis contains two independent task-based fMRI experiments with emotional faces that both follow similar hypotheses. Additionally, it contains two independent resting-state fMRI experiments. These doublings of experiments have their origins in independent experimental planning and full implementation of the OptiMD neuroimaging branch (subproject 5) later into this protocol as a co-enrollment trial, shortly after initiation.

Thus, two similar implicit emotional face processing fMRI-task experiments were performed on study-day 4 at baseline, one from SP5 before the resting period and the longitudinal one from this trial after. While both experiments are similar in their conceptualization, they are quite different in their execution. An overview of these differences in EFP-paradigms is described in Table 3.

Similarly, a (longitudinal) 22 min-long resting-state fMRI with closed eyes had already been part of this trial, when a (single) 5 min-long resting-state fMRI with open eyes was implemented originating from OptiMD SP5. Here as well, the OptiMD SP5 experiment was performed on study-day 4 at baseline, before the resting period and the other after

#### Study design after initial week of double-blind medication

While the initial week of double-blind medication was designed in a balanced four-arm pre/post-design, the design became unbalanced after this first week of medication, i.e. from study-day 12 onwards. Two study arms (placebo and agomelatine) were merged into the same monotherapy with agomelatine, thus retaining a psychopharmacological latency of seven days between them. Overall trial duration, with eight weeks of medication per protocol, was set long enough such that this latency would only cause a weak interference towards the end.

Furthermore, open-label medication from medication day 8 (i.e. study-day 12) onwards brought a risk of bias of study staff in regard to randomization. However, for the whole trial period, study staff would never know the difference between placebo and agomelatine randomization. Also, staff did usually not recall unblinding information in terms of randomized numbers. Therefore, there did not appear to be any bias of staff in regard to randomization overall.

### Protocol deviations during and after first week of double-blind medication

For a variety of reasons, there have been a number of protocol deviations, in particular after the initial week of double-blind study medication. Major deviations particularly affected three areas of trial performance: brain imaging, microbiome sampling and study medication.

The first of these deviations had been known before the trial was initiated. Due to retrofitting of a new research MR scanner, it was clear that MR acquisitions would initially be performed on three different MR scanners from the Siemens Magnetom device family. This implied the risk that neuroimaging data from the first nine recruited patients might not be reasonably usable in later group analyses depending on capacities for transformation of individual imaging data according to scanner calibration and quality control measures – a verdict on this is pending.

Microbiome samples had originally been planned to be immediately frozen at -80 °C as the primary storage method for all samples of this trial until later analysis per protocol. Two other storage methods with buffering solutions were only added later while the trial had already been ongoing. The majority of participating patients was discharged from hospital before the end of trial. Hence, they needed to come in for weekly psychometric assessments and for the third and final time-point of investigation including stool and blood sampling at study-day 60. At the time, acting study staff did not emphasize fresh stool sampling on site at end of trial but distributed the buffer storage methods one week before, once they were introduced into the trial. In consequence, stool sampling showed a large amount of missing fresh-frozen samples at the end of trial (see Table 2).

In section “Unblinding procedure”, a TDM approach was described as the original and initial plan for unblinding medication at study-day 12. TDM analysis focused on measurable escitalopram and mirtazapine serum levels as the half-life of agomelatine is too short for reliable determination in the morning, about ten hours after intake. This planned scheme was theoretically sufficient as no measurable serum levels of escitalopram or mirtazapine should have indicated randomization into agomelatine or placebo arm which both were designed to continue with agomelatine from study-day 12 onwards.

However, the cooperating laboratory of our department did not succeed in measuring the antidepressant substances after four to five days of intake correctly in all cases. This effect was at first noted by various reports of side effects that did not match the TDM-unblinding information. However, reporting side effects in a double-blinded clinical trial that do not fit the randomized substance is nothing unusual. Hence, time passed until we initiated verification procedures which led to a general change of the unblinding procedure nearly two years into the recruitment period and performed as described above.

Among 26 initially randomized patients (with no. 26 being the thirtieth trial participant), ten were wrongly unblinded by TDM. Seven of these were wrongfully analyzed to be escitalopram and two to be mirtazapine. These affected participants therefore received different study medication than planned per protocol from study-day 12 onwards. In regard to clinical pharmaceutical safety, this corresponded to an immediate switch of medication without cross-tapering after seven days of initial antidepressant medication. Besides changes in reported side effects, none of the affected participants were showing complications and none of them provided negative feedback.

Medication per protocol from study week three onwards was also affected by another phenomenon and led to significant additional protocol deviations. Section “General patient safety” described how this trial was performed on a ward that had in parts been furnished for clinical research with depressive inpatients. The close and nearly daily monitoring of depressed inpatients on a ward like this, however, has an immediate impact on the culture of medicating and has usually led to changes in psychotropic medication after about three weeks of unobserved clinical improvements. Regular teaching sessions and training on the protocol of this trial and how it followed official treatment guidelines were not sufficient to change this culture.

In regard to this protocol, there were two major biases in most of personnel on this ward that could not be remedied during the recruitment period of this trial: first, most team members did not feel that agomelatine would be equally efficacious as escitalopram or mirtazapine. Second, the team was not experienced with comparably strict trial protocols that intended to keep a psychopharmacological monotherapy unchanged over a course of eight weeks, besides possible elevations in dosages. Therefore, patient welfare was regularly taken as an argument to individually change the treatment plan as specified per study protocol. Hence, the final investigations at end of trial often do not reflect an antidepressant monotherapy as planned per protocol.

## Summary

Clinical depression, a growingly heterogeneous symptom complex, requires a better understanding of underlying pathology and further optimization of treatment options. Very different kinds of experimentation can be applied for these endeavors.

The extent of information collected longitudinally from the same participants of this trial in four different treatment arms encompasses a broad scope of perspectives onto possible neurobiological mechanisms in clinical depression with the hope that the longitudinal dynamics and individual perspectives can partly be connected.

The investigational modalities that have been implemented in this trial are already known to have substantial links and interrelations. For example, overregulation of the HPA-axis is related to pro-inflammatory processes and these, in turn, interact with lipid profiles. The gut microbiome appears to be equally intertwined in all of these processes with direct and indirect connections to the brain which itself tends to show signs of dysregulation and changed patterns of processing as known from functional neuroimaging in clinical depression.

This trial intends to provide an exploratory deep-phenotyping approach towards this complex disease entity and may possibly hint at further promising investigational efforts.

## Trial and protocol metadata

### Trial registration

The trial was registered on 30 July 2013 at the European Union Clinical Trials Register (EudraCT number: 2013-003370-27) as well as with the *Bundesinstitut für Arzneimittel und Medizinprodukte* (BfArM, Federal Institute for Drugs and Medical Devices, Prüfplancode (identifier): DMFMRI201303). The registration can be accessed at https://www.clinicaltrialsregister.eu/ctr-search/trial/2013-003370-27/DE.

### Funding

A major part of this investigator-initiated clinical trial has been supported by a grant from the *Bundesministerium für Bildung und Forschung* (BMBF, German Federal Ministry of Education and Research) within the German research consortium *Novel Strategies for Optimized Treatment of Depression* (OptiMD) as part of the national program *Forschungsnetz für psychische Erkrankungen* (FZPE, Research Network for Mental Disorders) with grant number 01 EE1401B (*Förderkennzeichen*) over Euro 1 428 748.97. Also, see section “OptiMD as framework for this trial”.

The funding agency had no role in study design and conduct of the trial; collection, management, analysis, and interpretation of data; preparation, review, or approval of the manuscript; and decision to submit the manuscript for publication.

The grant was provided for full funding of subproject 3 within the OptiMD consortium in order to investigate the relevance of microbiome composition for different types and stages of depression, for therapeutic responses therein and for side effects during antidepressant treatment. Hence, this grant covered gut microbiome analyses for all preclinical and clinical OptiMD trials, of which this trial was the major one. Besides covering funding for axis III (microbiome) as described here, the grant was replenished later to also support axis IV (inflammatory proteome) and axis V (lipidome) of this trial because of its suspected close links to microbiome regulation.

Funding for other axes of investigation described here, in particular in regard to axes I (neuroimaging) and II (HPA-axis regulation) was additionally provided by our Department of Psychiatry and Psychotherapy (Universität Regensburg, Regensburg, Germany). This partly included costs for study personnel, laboratory analytics and insurance. There was neither direct nor indirect funding by industry.

### Study protocol

#### Protocol versions

Version 1 as of 26 February 2015 (initiated: 20 January 2016)

Version 2 as of 18 May 2016 (Amendment No. 1: addition of OptiMD-SP5, as well as axis IV and V) Version 3 as of 06 July 2017 (Amendment No. 2: addition of healthy volunteers as additional controls)

Version 4 as of 27 July 2018 (Implementation of EU General Data Protection Regulation, GDPR)

#### Protocol title

The originally registered title for this clinical study, two and half years before initiation, has been “Functional MRI (fMRI) after challenge and treatment with antidepressants and their relation to the clinical course, to the Hypothalamus-Hypophysis-Adrenocortical Axis and the colon microbiome” (EudraCT). Around the same time, the title for subproject 3 within research consortium OptiMD, fully funded by BMBF, has been “Relevance of microbiome composition for subtypes of depression, therapeutic response and side effects during antidepressant treatment”. After substantial amendments and for publication of this protocol, the title was adapted to more clearly state the character and content of this trial.

### Amendments

#### Amendment No. 1

A first amendment was approved on 25 May 2016 by the *Ethikkommission bei der Universität Regensburg* (institutional ethics committee, Regensburg, Germany) under the same approval number as the original approval for the trial (approval number: 15-111-0075). This amendment comprised three requested changes. First, MR scanning time on the first imaging day (study day 4) was prolonged by 45 min in order to accommodate the MR experiments for OptiMD subproject 5 as described above. Furthermore, it contained additional taking of blood samples to enable the analysis of neuroinflammatory and lipidomic markers (as described for investigational axes IV and V, above).

#### Amendment No. 2

A second amendment was approved on 25 October 2017 by the *Ethikkommission bei der Universität Regensburg* (institutional ethics committee, Regensburg, Germany) and received a separate approval number 17-758-101. This amendment asked for the inclusion of matched healthy control subjects as an additional (negative) control group to the already existing (positive) patient control group (i.e. placebo arm). The amendment allowed for the recruitment of 40 healthy control subjects to enable the inclusion of 30 fully completing participants. This number of fully completing participants needed to be adapted to 25 due to impediments related to the COVID-19 pandemic and obligatory MR maintenance work in March 2021 that caused risk of losing comparability of acquired data with the patient cohort.

#### Implementation of the European Union General Data Protection Regulation (GDPR)

More than two years into trial recruitment, a European Union regulation on information privacy was commenced on 25 May 2018. This General Data Protection Regulation (GDPR) deeply affects privacy and human rights law in all countries of the European Union including data management for participants in clinical trials. Binding legal interpretations of this regulation were pending. Therefore, local institutional ethics committees referred to a guidance provided by *Arbeitskreis Medizinischer Ethik-Kommissionen in der Bundesrepublik Deutschland e.V.* (National Work Group of all German Institutional Ethics Committees for Medical Research) on 27 March 2018. For already ongoing clinical trials, it was recommended to supplement informed consent of current and future participants with a standardized two-page information sheet and verify that current and future consent included active confirmation of the new regulation and its consequences for research participants. Furthermore, all research staff, in particular study physicians were required to attend additional training in regard to GDPR and its consequences for good clinical practice in clinical trials. Hence, these proceedings did not require a dedicated formal amendment by the institutional ethics committee.

### Status of recruitment

Recruitment of participants has been completed. The clinical trial was initiated on 20 January 2016 and recruitment ended on 09 August 2021. During that period, 70 inpatients (24 female, 46 male) were included in this trial. Seven of these seventy participants dropped out, six of them before randomization and one of them during the first week of double-blind treatment. Twenty-five healthy, age- and gender-matched control subjects (10 female, 15 male) were additionally included in the trial between November 2020 and February 2021.

### Status of databases

Databases are still open and collection of some data is pending. All available data are pending quality control. Data collection is expected to be finalized by the beginning of 2026. Database locking should therefore be possible in the first half of 2026.

### Generated results

None of the data or results yielded from this trial have been submitted to or published in any academic journal, yet. A small and very limited pilot data set using dex/CRH testing material together with a small first subset of fMRI emotional faces paradigm was used for a doctorate thesis in psychology that was supervised by Mark William Greenlee at Universität Regensburg, Germany, and defended on 14 March 2019. Here, a pilot dataset dex/CRH testing was generated with a preliminary and different laboratory method than for the final and complete cohort of this trial. Data from trials that have been co-enrolled in this trial as described in Axis VII have partly begun dissemination of results.

## List of abbreviations

AE: adverse event
ACTH: (adrenocorticotropic hormone, i.e. corticotropin)
AGC: automatic gain control
AMG: Arzneimittelgesetz (German Federal Medicinal Products Act)
ANOVA: Analysis of Variance
ASV: Amplicon sequence variant
AUC: Area under the curve
AVP: arginine vasopressin (hormone)
BDI-II: Beck Depression Inventory (revised version)
BfArM: Bundesinstitut für Arzneimittel und Medizinprodukte (German Federal Institute for Drugs and Medical Devices)
BIS-11: Barrat Impulsiveness Scale – Version 11
BLASTN: Basic Local Alignment Search Tool for Nucleotides
BMBF: Bundesministerium für Bildung und Forschung (German Federal Ministry of Education and Research)
BW: Pixel bandwidth (MR imaging)
BOLD: Blood oxygen level-dependent (MR imaging)
CCD: Charge-coupled device
CFU: Colony-forming unit (corresponding to a viable single bacterial cell)
CGI: Clinical Global Impression scale
CRF: Case Report Form
CRH: corticotropin-releasing hormone
CRP: C-reactive protein
CONSORT: Consolidated Standards Of Reporting Trials
COVID-19: Coronavirus disease 2019 (see SARS-CoV-2)
CT: Computer Tomography
DAkkS: Deutsche Akkreditierungsstelle (German Accreditation Body)
Dex: dexamethasone
dex/CRH test: combined dexamethasone-suppression/corticotropin-releasing hormone stimulation test
DFC: Dynamic Functional Connectivity
DICOM: Digital Imaging and Communications in Medicine
DIN: Deutsches Instituts für Normung (German Institute for Standardisation)
DLPFC: Dorsolateral prefrontal cortex
DMN: default mode network
dNTP: deoxynucleotide triphosphate
DRD: Desire-Reason-Dilemma (fMRI experimental paradigm)
DSM: Diagnostic and Statistical Manual of Mental Disorders
DSMZ: Deutsche Sammlung von Mikroorganismen und Zellkulturen (German Collection of Microorganisms and Cell Cultures)
DST: Dexamethasone Suppression Test
DTI: Diffusion Tensor Imaging
DVD: Digital Video Disc
ECG: Electrocardiogram
EDTA: Ethylene-Diamine-Tetraacetic Acid
EFP: Emotional Face Processing (fMRI experimental paradigm)
EHI: Edinburg Handedness Inventory
EPI: Echo-planar imaging (MR imaging)
EudraCT: European Union Drug Regulating Authorities Clinical Trials
FA: here primary: fatty acid; may also mean: fractional anisotropy
FAME: Fatty Acid Methyl Esters
FC: Functional Connectivity
FIA: Flow Injection Analysis
FID: Flame Ionization Detector
FLAIR: fluid-attenuated inversion recovery (MR imaging)
fMRI: functional magnetic resonance imaging
FOV: Field of view
FT: Fourier Transform
FZPE: Forschungsnetz für psychische Erkrankungen (German Research Network for Mental Disorders)
GC: Gas-liquid Chromatography
GCP: Good Clinical Practice
GDPR: General Data Protection Regulation (European Union)
GRAPPA: Generalized Autocalibrating Partially Parallel Acquisitions (MR imaging)
HAMA: Hamilton Anxiety Rating Scale
HAMD: Hamilton Depression Rating Scale
HPA axis: Hypothalamic-Pituitary-Adrenal axis
5-HT: 5-hydroxytryptamine
HUFA: Highly Unsaturated Fatty Acid
IA: Isothermal amplification
ICD: International Classification of Diseases
ICH: International Council for Harmonisation of Technical Requirements for Pharmaceuticals for Human Use
IDE: Integrated Development Environment
IL: interleukin
ISO: International Organization for Standardization
IT: maximum injection time
KDEF: Karolinska Directed Emotional Faces (database)
LC: Liquid chromatography
LCFA: Long-Chain Fatty Acid
LED: Light emitting diode
LOQ: Limit of quantification
MALDI: Matrix-assisted laser desorption/ionization
MADRS: Montgomery-Åsberg Depression Rating Scale
MAO: monoamine oxidase inhibitor
MIAPE: Minimum Information About a Proteomics Experiment
MINI: Mini International Neuropsychiatric Interview
MIQE: Minimum Information for Publication of Quantitative Real-Time
PCR Experiments MP-RAGE: Magnetization prepared rapid gradient-echo (MR imaging)
MR(I): Magnetic Resonance (Imaging)
MS: Mass Spectrometer (or Spectrometry)
MTBE: methyl-tert-butyl ether
MUFA: Mono-Unsaturated Fatty Acid
NGS: Next Generation Sequencing
NIBSC: National Institute for Biological Standards and Control (UK)
NIST: National Institute of Standards and Technology (USA)
NPX: Normalized Protein eXpression (value, context: Olink PEA)
OD: Optical density
OptiMD: German Research Consortium “Novel Strategies for Optimized Treatment of Depression”
OR: odds ratio
OTU: Operational Taxonomic Unit
PACS: Picture Archiving and Communication System
PAT: Parallel acquisition techniques (MR imaging)
PCR: Polymerase Chain Reaction
PD-CAN: Phenotypic, Diagnostic and Clinical Domain Assessment Network Germany
PEA: Proximity Extension Assay
PET: Positron Emission Tomography
PUFA: Poly-Unsaturated Fatty Acid
RBC: Red Blood Cell (= erythrocyte)
rCBF: Regional Cerebral Blood Flow
rDNA: Ribosomal DNA
RDoC: Research Domain Criteria
rfMRI: Resting-state fMRI
RGB: Red, green and blue
RPA: Recombinase-polymerase amplification
RUO: Research Use Only
SAE: Severe adverse event
SARS-CoV-2: Severe acute respiratory syndrome coronavirus 2 (see COVID-19)
SC: Structural connectivity
SDA: Strand displacement amplification
SDMS: scientific data management system
SFA: Saturated Fatty Acid
SHAPS-D: Snaith-Hamilton Pleasure Scale – German version
SDMS: Scientific Data Management System
SNRI: Serotonin–norepinephrine reuptake inhibitor
SP: here: Suproject, with project number for OptiMD
SP35: short name for this trial
SPIRIT: Standard Protocol Items: Recommendations for Interventional Trials
SPRI: Solid-phase reversible immobilization
SSRI: Selective serotonin reuptake inhibitor
STAI: Spielberber’s State Trait Anxiety Inventory
STORMS: Strengthening The Organization and Reporting of Microbiome Studies
TCA: Tricyclic antidepressant
TCI: Temperament and Character Inventory
TDM: Therapeutic Drug Monitoring
TE: Echo time (MR imaging)
tfMRI: task-based fMRI
TFA: Trans Fatty Acid
TNF: tumor necrosis factor
TOF: Time-of-flight
TR: Repetition time (MR imaging)
VAS: Visual Analogue Scale
V1-V3: hypervariable region V1-V3 of the bacterial 16S ribosomal RNA gene
V3-V4: hypervariable region V3-V4 of the bacterial 16S ribosomal RNA gene
VLPFC: Ventrolateral Prefrontal Cortex
VM: Virtual Machine
WGS: Whole genome sequencing
WOCBP: Women of Childbearing Potential

## Declarations

### Data availability

Reliable de-identification of participant data is not truly possible for many datasets of this deep-phenotyping trial. For example, microbiome composition provides a fairly unique biomolecular fingerprint for each participant that tends to remain relatively stable over longer periods of time. Hence, the datasets of this study will not be publicly available due to concerns regarding participant anonymity. Reasonable requests for access to specific datasets should be directed to the corresponding author. We will support ethically sound and reasonable requests for collaboration.

### Conformance statement for reporting guidelines and recommendations

Publication of this protocol conforms to SPIRIT 2013 guideline (SPIRIT = Standard Protocol Items: Recommendations for Interventional Trials, [348]) as well as with the CONSORT 2010 statement (Consolidated Standards Of Reporting Trials, [349]). Furthermore, it also complies with the latest CONSORT 2025 statement [346] where applicable. All required items are part of this protocol as verified with the SPIRIT and CONSORT checklists.

The extensive multi-modality of the investigations described in this protocol has further motivated us to strive for conformance with the following guidelines and recommendations wherever applicable, be it in this protocol or for future reporting of results: COBIDAS recommendations (OHBM Committee on Best Practices in Data Analysis and Sharing, [350]), STORMS checklist (Strengthening The Organization and Reporting of Microbiome Studies, [351]), MIQE (Minimum Information for Publication of Quantitative Real-Time PCR Experiments, [352]), MIAPE (Minimum Information About a Proteomics Experiment, [353]), and Lipidomics Minimal Reporting Checklist [354]. Solid novel standards and future recommendations will additionally be considered accordingly.

All authors of this protocol have agreed to this conformance statement on guidelines and recommendations and reviewed the protocol content for their field of expertise, accordingly.

### Citation diversity statement

There is increasing awareness of a need to reduce citation bias as, for example, identified in regard to gender in neuroscience [355] and authors have been recommended to consider their own bias in their publications [356]. In 2020, Dworkin et al. reported observed citation rates in regard to gender from five top-ranking neuroscience journals from 1995 to 2020 in Nature Neuroscience as 61.7% (MM), 23.6% (WM), 9.0% (MW) and 5.8% (WW) while expected citation rates disregarding gender were calculated to be 58.6% (MM), 25.3% (WM), 9.4% (MW) and 6.7% (WW). The letters describe gender of first and last author of a publication.

Identifying citations for this protocol was blind in regard to gender of authors that may have been derived from their first names after looking into author lists more closely. The goal was to cite the most original and appropriate references available, independent of who wrote them. While doing so, there have been no preferences towards specific colleagues, academic institutions or countries.

The citation diversity statement has its roots in gender imbalances in academic science. Among the many sources for possible discrimination described below, gender is probably the only aspect that can be identified in a list of author names. While gender imbalances remain a problem that needs to be addressed and remedied, we have observed many more imbalances that cause severe injustice in science and that cannot be derived from author names or their affiliations.

We take a strong position against discrimination and invalidation and agree with the importance of equality and diversity on all levels of life and thus, also of research endeavors. We therefore support the equal support and gratification of all contributors independent of their gender, age, income, country of origin, religion, sexual preference, outer appearance, personality accentuation, levels of assertiveness or their neurodiversity, just to name some prominent aspects among many possible sources of inequality.

### Disclosure statement on AI-tool usage

This study protocol has been prepared, written and reviewed without any assistance from AI-tools.

### Author Contributions

CRediT (Contributor Role Taxonomy), 14 machine-readable roles (see https://credit.niso.org)

André Manook: Conceptualization, Data Curation, Formal Analysis, Investigation, Methodology, Project Administration, Resources, Software, Supervision, Validation, Visualization, Writing – Original Draft, Writing – Review & Editing.

Andreas Hiergeist: Data Curation, Formal Analysis, Investigation, Methodology, Resources, Software, Validation, Visualization, Writing – Review & Editing.

Gerhard Liebisch: Formal Analysis, Investigation, Methodology, Resources, Validation, Writing – Review & Editing.

Ulrich Dischinger: Investigation, Methodology, Resources, Validation, Writing – Review & Editing.

Agnese Petrera: Formal Analysis, Investigation, Methodology, Resources, Validation, Writing – Review & Editing.

Jens Schwarzbach: Conceptualization, Methodology, Software, Supervision, Writing – Review & Editing.

André Gessner: Conceptualization, Resources, Writing – Review & Editing.

Oliver Gruber (roles for co-enrollment trial OptiMD SP5 as part of this trial): Conceptualization, Data Curation, Formal Analysis, Funding Acquisition, Methodology, Project Administration, Resources, Validation, Writing – Review & Editing.

Thomas C. Baghai: Conceptualization, Funding Acquisition, Project Administration, Resources, Supervision, Writing – Review & Editing.

## Acknowledgments

First and foremost, we express our deepest gratitude to all patients and healthy volunteers who were willing to participate in this trial.

We thank Michael Koller for inspiring the writing of this protocol and his colleague Florian Zeman for valuable statistical review of our final sample size estimations, as well as Marc Greenlee, Thomas Frodl and Monika Sommer for initial statistical support with sample size estimations; Florian Hartig for inspiring a protocol overview for this manuscript; Marcus Ising for inspiring the implementation of copeptin analysis into to this trial and for reviewing investigational axis II; Clemens von Schacky for performing Omegametrix analyses and for reviewing text on its methodology; Christian Stroszczynski for granting access to his MRI facilities and personnel at Universitätsklinikum Regensburg on weekends during scanner retrofitting on our campus; Ralph Burkhardt for connecting us to investigators and co-authors GL and UD and for hinting us at Olink technology as a potential alternative to ELISA-based multiplex assays. Also, we would like to thank Johannes Weigl for assisting in trial recruitment and psychometric assessments; former and current team members from our biomedical imaging group for assisting with MR operation, including Seth Levine, Philip Seidel, Marlene Tahedl, Viola Wagner and Lisa-Marie Brunner. Furthermore, we thank Wilhelm Malloni for continuous support in MR technology and maintenance; Karin Völlner and Ulrike Stadler for valuable support as study nurses. We very much appreciate expert technical assistance in stool processing and microbiome sequencing by Ursula Holzinger and Christine Irtenkauf and expert technical assistance in Olink Explore analysis by Jennifer Behler and Bianca Eichner.

